# UK Biobank release and systematic evaluation of optimised polygenic risk scores for 53 diseases and quantitative traits

**DOI:** 10.1101/2022.06.16.22276246

**Authors:** Deborah J. Thompson, Daniel Wells, Saskia Selzam, Iliana Peneva, Rachel Moore, Kevin Sharp, William A. Tarran, Edward J. Beard, Fernando Riveros-Mckay, Carla Giner-Delgado, Duncan Palmer, Priyanka Seth, James Harrison, Marta Futema, Genomics England Research Consortium, Gil McVean, Vincent Plagnol, Peter Donnelly, Michael E. Weale

**Author notes:** Genomics England Research Consortium author list available from https://www.genomicsengland.co.uk/about-gecip/publications/. These authors contributed equally to the work (Deborah J. Thompson, Daniel Wells, Saskia Selzam). These authors jointly supervised the work (Peter Donnelly, Michael E. Weale).

## Abstract

We present and assess the UK Biobank (UKB) Polygenic Risk Score (PRS) Release, a set of PRSs for 28 diseases and 25 quantitative traits being made available on the individuals in UKB. We also release a benchmarking software tool to enable like-for-like performance evaluation for different PRSs for the same disease or trait. Extensive benchmarking shows the PRSs in the UKB Release to outperform a broad set of 81 published PRSs. For many of the diseases and traits we also validate the PRS algorithms in other cohorts. The availability of PRSs for 53 traits on the same set of individuals also allows a systematic assessment of their properties, and the increased power of these PRSs increases the evidence for their potential clinical benefit.

## Introduction

Polygenic risk scores (PRSs) provide a personalised measure of genetic liability of disease, combining genetic risk information from across the genome ^1,2^. PRS scores can also be used to measure the genetic contribution to quantitative traits (for simplicity, we also use the term PRS here for such traits). The field is growing rapidly, with advances in methods ^3^, reporting standards ^4^, and cataloguing ^5,6^. There is also mounting evidence for their clinical utility ^7^. For example, a PRS algorithm for coronary artery disease has similar predictive power to LDL cholesterol ^8^, an established clinical risk factor, while PRS algorithms for coronary artery disease ^9^ and for breast cancer ^10^ have been shown to identify groups with equivalent risk to clinically relevant rare variant carriers. There has been extensive recent interest in PRSs, with a large number now available; in some cases more than 100 PRS algorithms have been published for the same disease ^6^.

Prospectively collected biobanks such as UK Biobank ^11^ (UKB) play an important role in enabling PRS development, evaluation, and application ^12^. They can provide data for PRS training, but more importantly they provide large representative population samples for evaluation of PRS scores in multiple contexts, including ancestry. They also provide a broad base of other clinical and biomolecular information, both to train and evaluate multi-factor clinical risk models and for other research applications ^13–17^. To enable PRS research and development, we present the UK Biobank PRS Release, which comprises well-powered PRS scores for 28 diseases and 25 quantitative traits, together with associated data and analysis. We present two PRS sets, a Standard Set with scores calculated for all individuals in UKB, trained on external data only, and an Enhanced Set calculated for a Testing Subgroup of 104,231 individuals in UKB, which has the advantage of being trained on external data plus additional training data from a separate subgroup of UKB.

A level playing field is essential for fair comparisons and evaluations of PRS performance ^18^. Reported performance can be influenced by many factors, including the choice of performance metric, covariate adjustment, demographic and study properties of the evaluation cohort, and decisions on how the phenotype was defined ^12^. These choices can confound inferences about the performance of the underlying PRS algorithm or PRS methodologies. To address these issues, we have built a unified pipeline for PRS evaluation, constructing a standardised Testing Subgroup within UKB and a standardised set of disease and quantitative trait definitions. We have made this pipeline available as an open source tool within the UK Biobank Research Access Platform, along with the associated phenotype definitions, to allow other researchers to check reported metrics and perform evaluations of their own or others’ PRS scores against the UK Biobank Release. We have used this pipeline to benchmark the UK Biobank PRS Release, comparing the Standard and Enhanced PRS sets to each other and to 81 PRS scores from published algorithms, with favourable results.

One recognised limitation of current PRSs is the effect of ancestry (herein, ‘ancestry’ refers to genetically inferred ancestry) on PRS performance ^19,20^. Ultimately, a resolution to this issue requires more representative training data, but in the short term there is a requirement for appropriate cross-ancestry measurement and reporting of PRS performance. Our Comparison Tool allows such standardised measurement and reporting, and we use it to quantify the diminution of performance in non-European ancestry individuals, for the UKB PRS Release and for many other PRSs.

In order to understand generalisability in PRS performance, and as a check against any UKB-specificity, we adapted the evaluation pipeline to apply it to other trait definitions and testing sets in additional UK-based and US-based cohorts. Variation in phenotype definition across cohorts, and countries, can confound performance comparisons in this context ^12^. We found similar performance in the UK-based 100,000 Genomes Project dataset ^21,22^, which shares the same UK national system for hospital records and which therefore provides some measure of homogeneity in disease phenotype definition.

The predictive power of PRSs is one feature that affects their potential impact in healthcare. The availability of a set of PRSs more powerful than most of those previously studied allows a reassessment of aspects of this clinical potential. As earlier studies have observed ^9,10,23–25^, we show that risk profiles of individuals with appropriately high PRS scores are similar to those seen in carriers of known rare pathogenic variants, that these high-PRS individuals account for a much higher fraction of disease, and of early onset disease, and that the PRS score modulates the effect of rare pathogenic variants. The increased power of the PRSs described here increases the quantitative impact of these effects.

Previous PRS studies have largely focussed on performance of a relatively small number of traits. The availability of PRSs for 53 traits within a large, richly phenotyped cohort allows a systematic assessment of their properties and provides an opportunity to explore a range of PRS analyses, and draw cross-trait conclusions regarding PRS properties. In addition to quantifying the effects of ancestry, sample size, and generalisability, we find that correlations between PRS scores for different traits are generally low, and that PRS effect sizes are usually larger in people of younger age. These resources should further enhance the UKB resource, facilitate the ongoing improvement of PRS algorithms and methodologies, particularly across genetic ancestries, and accelerate development and validation of new research or clinical use cases for PRSs.

For clarity, in what follows we distinguish three closely related concepts. Throughout, we use: (1) *PRS score*, or just PRS, for the score assigned to a particular individual; (2) *PRS algorithm* for the function which calculates the PRS score from genetic data on an individual; and (3) *PRS methodology* for the approach used to determine a particular PRS algorithm.

## Results

### Performance of the PRS Release in UK Biobank

We generated a Standard PRS Set, calculated on all UKB individuals, for 28 diseases and 8 quantitative traits, by meta-analysing multiple external GWAS sources (Supplementary Tables 1 and 2 and Supplementary Information). We also generated an Enhanced PRS Set, calculated on individuals in the UKB Testing Subgroup only, for 28 diseases and an expanded list of 25 quantitative traits, in which the underlying PRS algorithm was trained from both external and a subset of UKB data (Supplementary Tables 3 and 4, UKB phenotype definitions in Supplementary Table 5). To ensure uniformity of evaluation, the performance of both PRS sets was evaluated on a standard array of metrics in the same group of UKB testing individuals. The UKB Testing Subgroup (Supplementary Table 6) comprised 82,346, 9,543, 9,478 and 2,864 individuals of predominantly European, South Asian, African, and East Asian ancestry, respectively, and was designed to maximise the representation of non-European ancestries. (Numbers of non-European ancestry individuals were too small to provide well-powered training data.) Individuals in the Testing Subgroup were selected to ensure no overlap with the previously defined White British Unrelated (WBU) subgroup ^11^, which was used to contribute training data for the Enhanced PRS Set (see Supplementary Information).

We quantify PRS performance in multiple ways. Cumulative incidence plots provide a useful visual tool for comparing disease incidence over time among individuals grouped according to their PRS for that disease. Notwithstanding known issues in UKB healthy bias ^26^ and underreporting of some diseases (e.g. type 2 diabetes is reported mainly from primary care records, which are only available for ∼40% of UKB participants), Figure 1 reveals large differences in disease incidence across ages for groups defined by the Enhanced PRS Set, further emphasising the potential of PRSs for powerful individual risk stratification.

**Figure 1.**
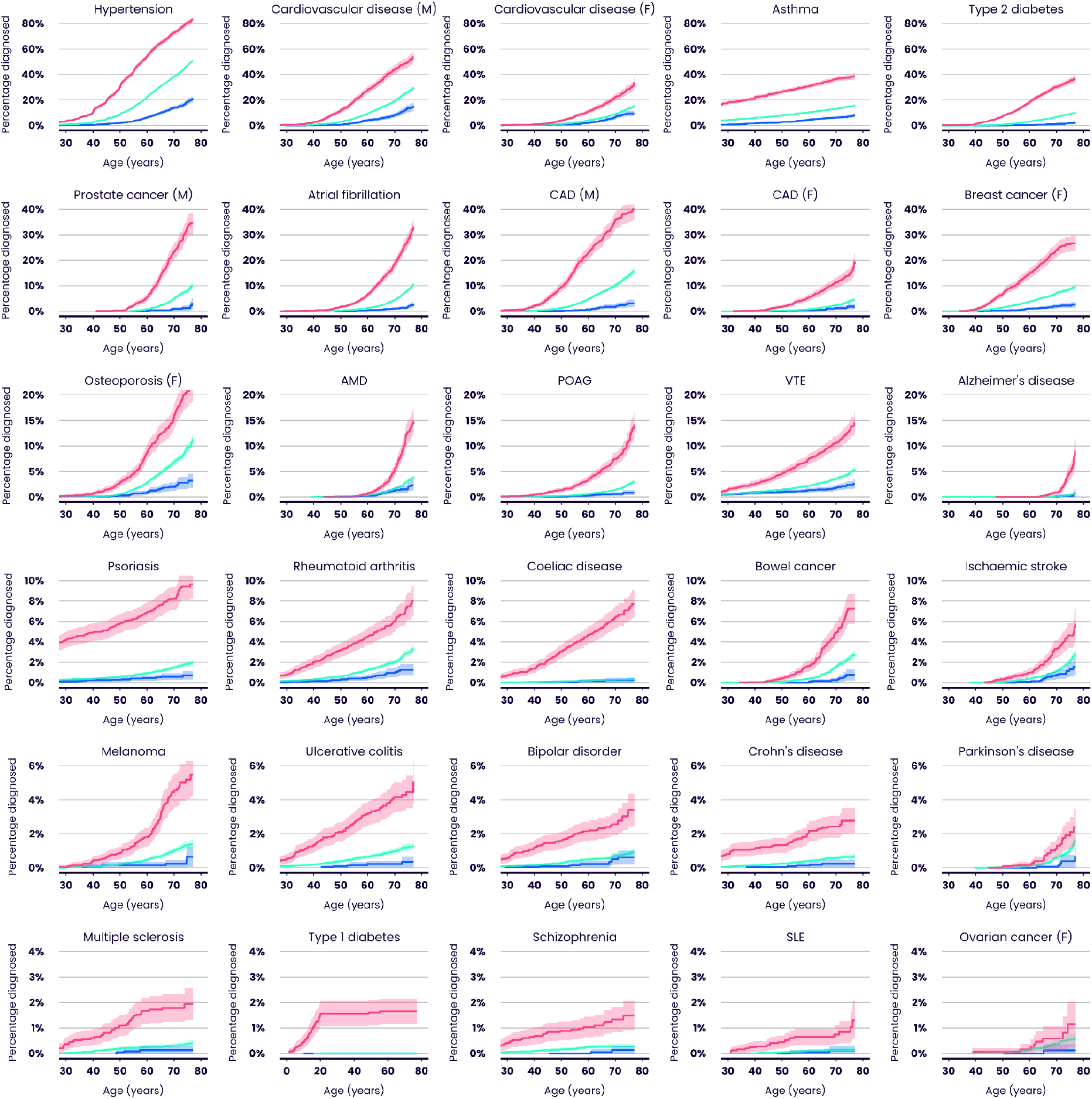
Cumulative incidence plots illustrating the predictive performance of the UK Biobank PRS Release for 28 diseases in European ancestry individuals (Enhanced Set). Each plot shows the estimated percentage of individuals diagnosed with a specific disease by a given age, for three groups within the UKB Testing Subgroup defined only by their PRS scores. Colours indicate the highest 3% (red), median 40-60% (green) and lowest 3% (blue) of the Enhanced PRS distribution. M = male, F = female. Shadings indicate 95% confidence intervals. Type 1 diabetes age range has been widened to reflect early onset. AMD = age-related macular degeneration. CAD = coronary artery disease. POAG = primary open angle glaucoma. SLE = systemic lupus erythematosus. VTE = venous thromboembolic disease. Throughout, ovarian cancer refers specifically to epithelial ovarian cancer.

Figure 2 and Supplementary Tables 7 and 8 quantify performance properties of the Enhanced PRS Set, for disease and quantitative traits, in the Testing Subgroup. Performance is assessed across multiple ancestries, subject to a minimum threshold on case numbers, as reliable performance metrics cannot be evaluated in some ancestries for diseases which are rare in UKB. In individuals of European ancestry, performance in disease traits (measured by odds ratio per SD of PRS, from logistic regression adjusting for age and sex) was variable, ranging from 3.87 (type 1 diabetes) to 1.39 (epithelial ovarian cancer), with median 1.85. Performance in quantitative traits in individuals of European ancestry (measured by effect on standardised trait per SD of PRS, from linear regression adjusting for age and sex) ranged from 0.427 (estimated BMD T-score) to 0.230 (docosahexaenoic acid), with median 0.274. Similar patterns were seen for the Standard PRS Set (Supplementary Figure 1 and Supplementary Tables 7 and 8), and also when performance was evaluated on the disease traits using the area under the receiver operating characteristic curve (AUC) as an alternative performance metric (Supplementary Figure 2 and Supplementary Table 7).

**Figure 2.**
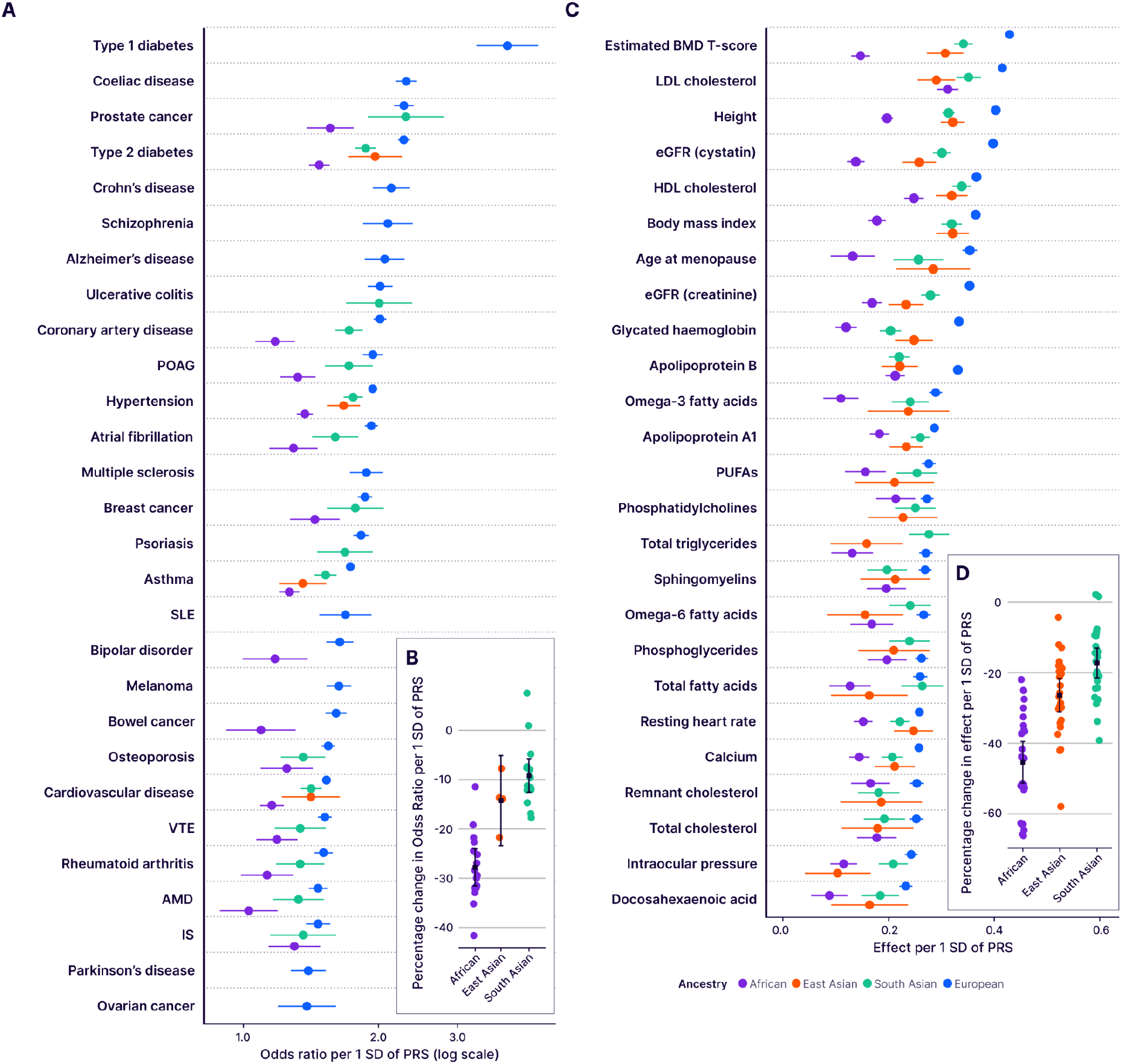
Predictive performance of the UK Biobank PRS Release (Enhanced Set) by ancestry. Performance (odds ratio, or effect on standardised quantitative trait, per SD of PRS, adjusting for age and sex), measured in the independent UKB Testing Subgroup, of the disease traits (**A**) and quantitative traits (**C**), stratified by genetically inferred ancestry. Results for non-European ancestries are shown if at least 100 cases are available for testing. Relative change in performance in non-European ancestries compared to European ancestry for disease traits (**B**) and quantitative traits (**D**). Bars indicate 95% confidence intervals (CI). eGFR = estimated glomerular filtration rate. BMD = bone mineral density. HDL/LDL = high/low density lipoprotein. PUFAs = polyunsaturated fatty acids. Refer to Figure 1 legend for disease abbreviations.

As previously reported for other PRSs ^19,20^, performance in individuals of non-European genetic ancestry was lower than in those of European ancestry (Figure 2B and 2D). Averaging across all diseases, the odds ratio per SD of PRS reduced by 9.1% in individuals of South Asian ancestry (95% CI 5.7-12.5%), and by 14.3% in individuals of East Asian ancestry (95% CI 5.1-23.4%). Consistent with previous observations, the largest reduction was in individuals of African ancestry (27.6%, 95% CI 23.8-31.4%). Reductions in effect per SD were numerically larger for the 25 quantitative traits (Figure 2D), but because OR per SD and effect per SD are on different scales ^27^, changes for the quantitative traits cannot be compared to those for the disease traits. Averaging across all quantitative traits, the effect size of the PRS reduced by 18.1% in individuals of South Asian ancestry (95% CI 13.6-22.6%), 26.6% in individuals of East Asian ancestry (95% CI 21.7-31.6%), and 46.1% in individuals of African ancestry (95% CI 40.7-51.6%).

As expected, the Enhanced PRS Set outperformed the Standard PRS Set (Supplementary Figure 3, Supplementary Table 9). Comparing the two types of PRS for 28 diseases and 8 quantitative traits, there were 27 instances where the Enhanced PRS significantly outperformed (at a nominal 5% level) the Standard PRS in European ancestry individuals, and no instance where the Standard PRS significantly outperformed the Enhanced PRS (median increase in Enhanced relative odds ratio = 1.03, range 0.97-1.16). In a separate comparison, looking across diseases, the relationship between training sample size and predictive performance was noisy (Supplementary Figures 4 and 5), suggesting that other trait-specific factors, such as heritability, genetic architecture, and prevalence, are also important in determining performance ^12,28^.

We benchmarked the UK Biobank PRS Release against PRS scores generated from 81 published algorithms, across a range of disease and quantitative traits (Figure 3 and Supplementary Tables 7, 8, 9 and 10). Among European ancestry individuals, the Enhanced PRS Set in the UKB Release significantly outperformed (at a nominal 5% level) all comparator PRSs for all disease traits apart from systemic lupus erythematosus, cardiovascular disease, Parkinson’s disease, and epithelial ovarian cancer, and for all quantitative traits apart from total cholesterol. For these latter diseases, with one exception (epithelial ovarian cancer), the point estimate for performance of the Enhanced PRS was greater than all comparator PRSs.

**Figure 3.**
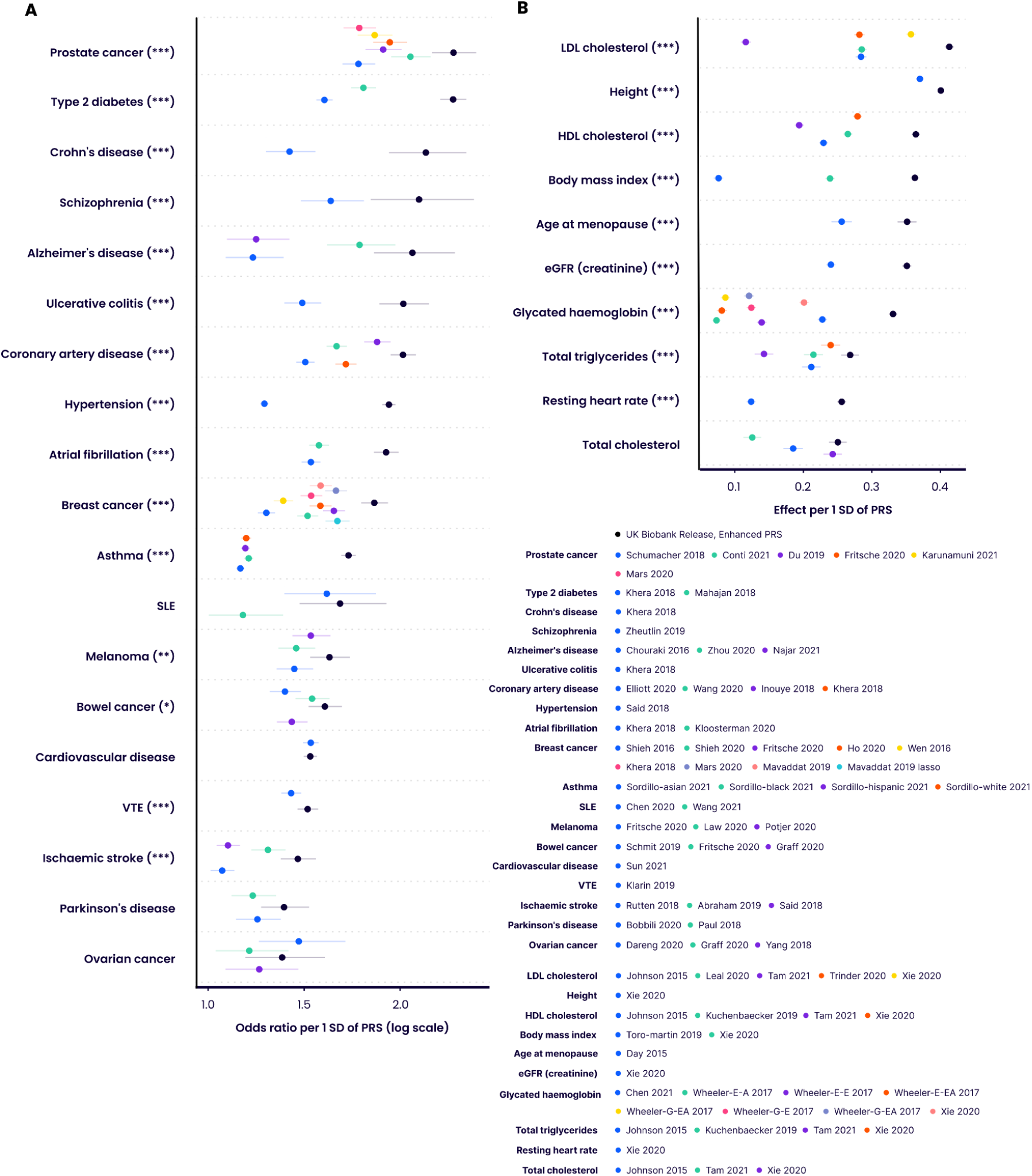
Predictive performance of the UK Biobank PRS Release against published comparator PRSs. Performance (odds ratio, or effect on standardised quantitative trait, per SD of PRS, adjusting for age and sex) in the independent UKB Testing Subgroup (European ancestry) of the Enhanced PRS sets for disease traits (**A**) and quantitative traits (**B**), for those traits for which there are published PRS algorithms (citations provided in Supplementary Table 10). Bars indicate 95% confidence intervals, the log scale makes these larger and asymmetric for lower values in panel **B**. Asterisks indicate significance level for difference in effect size between the Enhanced PRS and the nearest comparator PRS (5000 bootstraps): * p<0.05, ** p<0.01, *** p<0.001. Wheeler-E-A, Wheeler-E-E and Wheeler-E-EA refer respectively to the African, European and East Asian ancestry versions of the Wheeler 2017 PRSs for glycated haemoglobin using erythrocytic variants. Wheeler-G-A, Wheeler-G-E and Wheeler-G-EA refer respectively to the African, European and East Asian ancestry versions of the Wheeler 2017 PRSs for glycated haemoglobin using glycemic variants. Refer to Figure 1 and 2 for disease and quantitative trait abbreviations respectively.

We noted above that absolute performance of the UK Biobank PRS Release was reduced in non-European ancestries. This is also true of the comparator PRSs. When compared within each ancestry group, the UK Biobank PRS Release performed favourably across all traits relative to comparator PRSs (Supplementary Figures 6 and 7). We also note that differences in absolute risk can sometimes compensate for differences in discriminatory performance. For example, the odds ratio per SD of the Enhanced PRS for type 2 diabetes is lower in individuals of South Asian ancestry (OR per SD = 1.87, 95% CI 1.77-1.98), compared to European ancestry (OR per SD = 2.28, 95% CI 2.21-2.35). But because the disease is more prevalent in South Asian ancestry individuals, there is a bigger separation in absolute risk between the top and bottom 3% of the PRS distribution in South Asian compared to European ancestry individuals (Figure 4).

**Figure 4.**
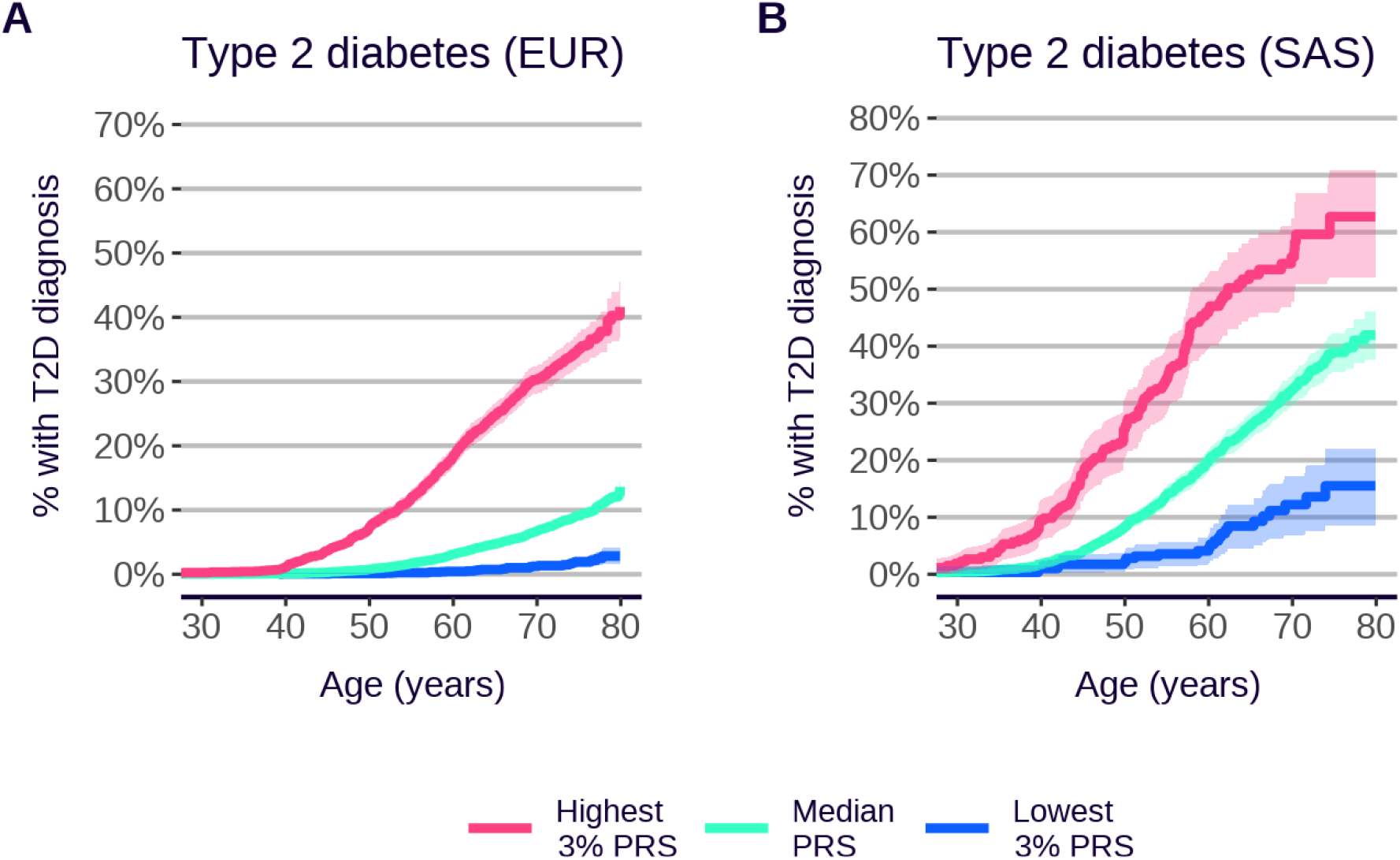
Cumulative incidence of type 2 diabetes, by ancestry, Enhanced PRS Set. Evaluated in the UKB Testing Subgroup. **A**. European ancestry (EUR). **B**. South Asian ancestry (SAS). Shaded areas indicate 95% CI.

### PRS risk profiles compared to high-risk variant carriers

Health systems already use genetics to identify individuals at increased risk of particular diseases, including some common diseases (e.g. breast cancer and heart disease), but to date this has focussed on carriers of high-risk rare mutations ^29,30^. PRS scores provide a way to measure a separate component of genetic risk, via the accumulation of many small-effect common variants, and so it is of interest to compare the risk profiles of these two components. Taking familial hypercholesterolemia (FH) and breast cancer (BC) as examples, and following previous work ^9,10^, we find that individuals possessing high PRS scores have a cumulative incidence risk profile similar to carriers of high-risk variants in known functional genes identified from available whole exome sequencing in the same cohort.

Our analysis of UKB shows that the risk by age 70 of coronary artery disease (CAD) for carriers of a pathogenic or likely-pathogenic mutation in one of the four major FH genes is 13.0% (95% CI 10.0-15.9%), in line with previous studies ^31,32^ (see Supplementary Materials for definition of mutation carriers). A similar risk is seen in individuals who are in the top 19% of the CAD Enhanced PRS distribution (risk by age 70 = 14.2%, 95% CI 13.6-14.9%, Figure 5A). Risks are higher both for mutation carriers and for high PRS individuals who are not using statins for primary prevention ^33^. Restricting the analyses to the subset of 176,564 participants for whom primary care prescribing data are available and who have no recorded statin prescription (other than prescriptions following a CAD diagnosis), the risk to age 70 in carriers is 17.3% (95% CI 10.0-23.9%), which is similar to that seen in statin-free individuals in the top 8% of the Enhanced PRS distribution (risk by age 70 = 18.1%, 95% CI 16.2-20.0%, Figure 5B and Supplementary Table 11). (Note that all analyses here have caveats. For example while FH carriers not on statins will have a higher CAD risk, these individuals might tend to have lower cholesterol levels to have avoided statin prescription, an effect which will tend to reduce their CAD risk compared to other FH carriers.)

**Figure 5.**
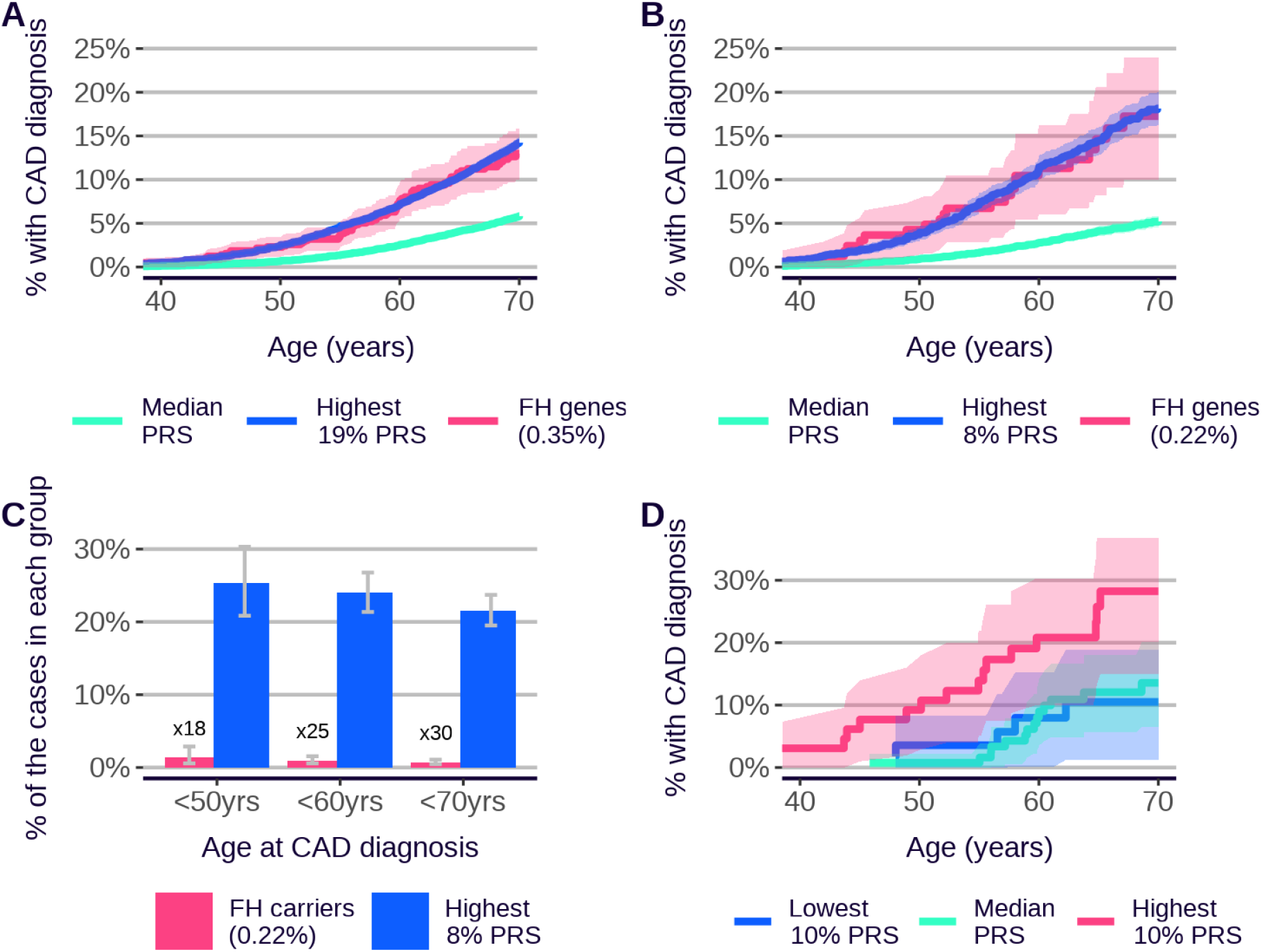
PRS risk profiles compared to functional variant carriers. **A** Cumulative incidence of coronary artery disease (CAD) for 656 carriers (0.35% of evaluation group) of pathogenic or likely-pathogenic mutations in FH genes (*APOB, APOE, LDLR* or *PCSK9*) (red, evaluated in UKB European ancestry individuals for whom whole exome sequencing data were available), compared to individuals in the top 19% percent of the Enhanced CAD PRS distribution (blue) (percentile chosen such that the risk up to age 70 is similar to that for mutation carriers), and the median 40%-60% of the PRS (green). The PRS risks are evaluated in the UKB Testing Subgroup (European ancestry). **B** Cumulative incidence of CAD for 164 carriers (red, 0.22% of restricted evaluation group of individuals with primary care data linkage and no recorded statin prescription prior to CAD event), compared to individuals in the top 8% percent of the Enhanced CAD PRS distribution (blue) and the median 40%-60% of the PRS (green). The PRS risks are evaluated in the UKB Testing Subgroup (European ancestry), additionally restricted to individuals with no recorded statin prescription prior to CAD event. **C** Percentage of CAD cases diagnosed in individuals aged <50, <60, or <70 years that occurred in carriers of pathogenic or likely pathogenic FH gene mutations (red) or in individuals in the top 8% of the Enhanced PRS distribution (blue), restricted to individuals with no recorded statin prescription prior to CAD event. The ratio between the number of high PRS cases and mutation carrier cases in each age group is shown on the plot. **D** Cumulative incidence plots for carriers of pathogenic or likely pathogenic mutations in FH genes (not restricted to individuals with no statins use), with additional stratification by the top 10% (red), median 40%-60% (green), and bottom 10% (blue) of the Standard CAD PRS (the Standard PRS is used here to maximise the number of individuals with both whole exome sequencing data and a PRS value available for analysis). Bars and shadings indicate 95% CI.

For the example of female breast cancer, the top 0.2% of the breast cancer Enhanced PRS distribution is associated with an equivalent level of lifetime risk to deleterious mutations in the *BRCA1/2* genes. If we instead consider breast cancer-associated mutations across a broader range of genes (*ATM, BRCA1, BRCA2, CHEK2* and *PALB2*), the carrier risk is equivalent to the top 3% of the PRS distribution (Supplementary Figure 8, see Supplementary Materials for definition of mutation carriers).

Previous studies ^9,10^ have shown that individuals at equivalently high risk due to PRS typically outnumber high-risk variant carriers, often massively so. Focusing on non-statin users, and on the individuals in the top 8% of the Enhanced UKB CAD PRS distribution with similar lifetime CAD risk to FH carriers (combined carrier frequency 0.22%), the high PRS group accounts for between 18 and 30 times the number of CAD events, depending on age (Figure 5C). Both high PRS and FH mutation carriers convey a higher risk at younger ages (Supplementary Figure 9), but the effect is stronger for FH carriers, explaining the reduced ratio when restricted to age less than 50.

Following previous work ^23–25^, we also find that carriers of high-risk variants can have their disease risk further modulated by their PRS score (Figure 5D), providing one explanation for the phenomena of incomplete penetrance and variable expressivity ^34^. The more-powerful PRSs described here should have a larger effect in modulating the impact of rare variants than has previously been observed.

### PRS risk profiles with age

Cumulative incidence plots across multiple traits and ancestries (Figure 1 and Supplementary Figures 10 and 11) indicate that people with high PRS are at elevated risk of disease throughout the lifecourse, and this observation, coupled with the fact that a person’s PRS score is invariant and can be measured early in life, could be clinically relevant, given that identifying high-risk individuals at younger ages is often challenging via other methods. There is, additionally, evidence for age-dependent PRS performance in some diseases, with a tendency for greater discrimination at younger ages ^35^, and this could add further weight to the use of PRS for risk identification in younger people. We are now able to assess this question systematically for 28 diseases. Figure 6 shows that many of the diseases in the UK Biobank PRS Release display evidence for larger PRS effect size (log hazard ratio per SD of PRS) in younger compared to older individuals in UKB (9 out of 23 diseases with nominal significance, 13 consistent with the null of no age effect and 1 (age-related macular degeneration) with nominal significance in the other direction). This supports earlier observations ^35^, and may point to a general principle that environmental effects on disease risk are more cumulative than genetic effects over the lifecourse.

**Figure 6.**
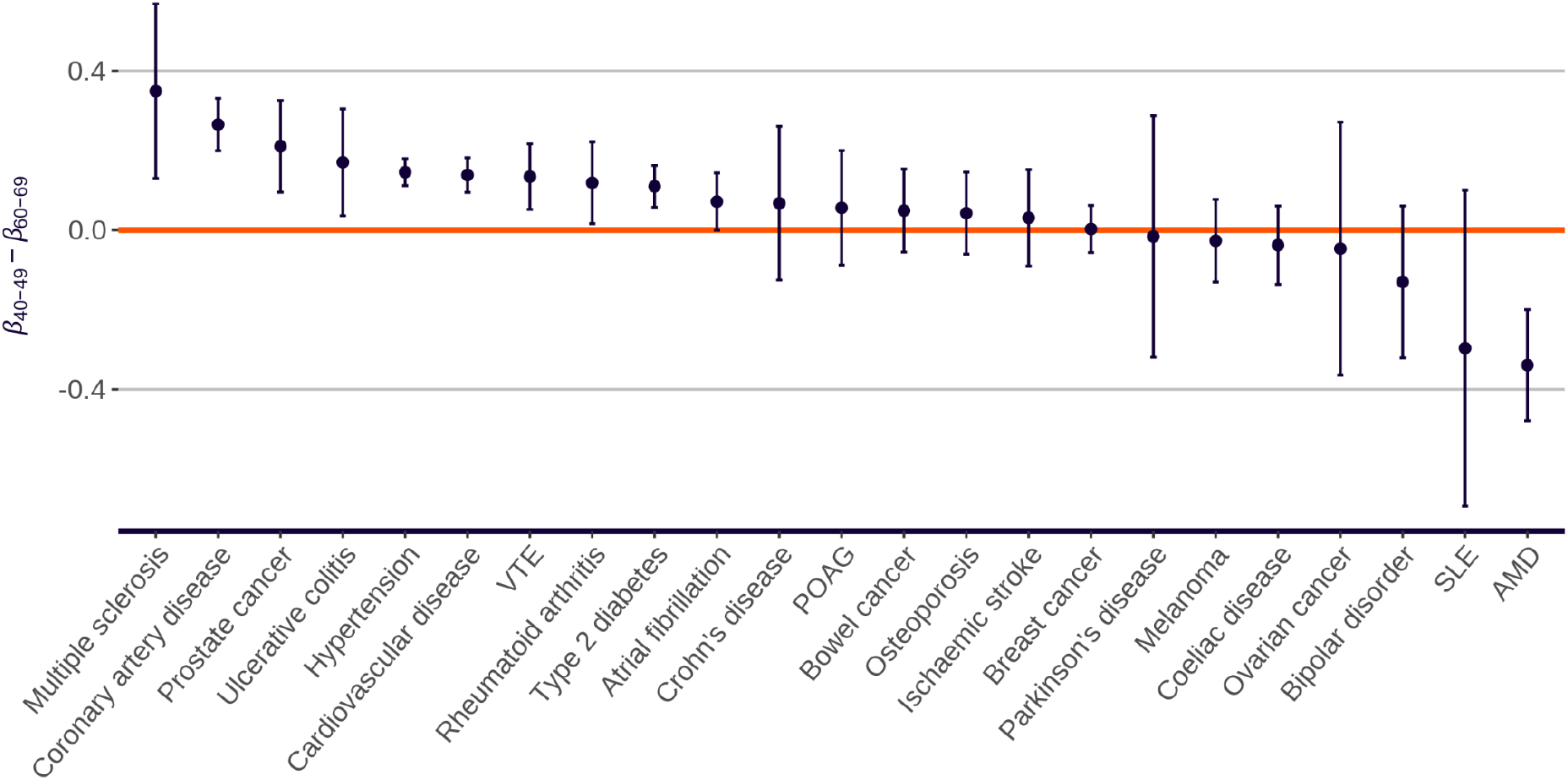
Change in PRS effect size with age. Difference in PRS effect size (log hazard ratio per SD of PRS, based on incident events over the next 10 years) between younger (40-49) and older (60-69) age-at-first-assessment groups. Standard PRSs are presented and evaluated in all UKB to maximise case numbers, for all diseases except Alzheimer’s disease, asthma, psoriasis, schizophrenia and type 1 diabetes, which primarily develop outside the UKB age range. Bars indicate 95% CI. Refer to Figure 1 for disease abbreviations.

### Multivariate PRS properties

We assessed PRS correlations across the 53 diseases and quantitative traits in the UKB PRS Release. Correlations between PRS scores for the disease traits are generally low (Supplementary Figure 12, Supplementary Tables 12 and 13). The only disease-disease correlations greater than 0.5 are between coronary artery disease and cardiovascular disease (r = 0.80, Enhanced Set), hypertension and ischaemic stroke (r = 0.78, Enhanced Set) and Crohn’s disease and ulcerative colitis (r = 0.54, Standard/Enhanced Set). Correlations among PRSs for quantitative traits are stronger, with strong correlations in particular among the traits related to lipid biology, and with the strongest correlation between phosphatidylcholines and phosphoglycerides (r = 0.99, Enhanced Set). The only correlation greater than 0.5 between a disease trait and a quantitative trait is between primary open angle glaucoma and intraocular pressure, a known major risk factor (r = 0.62, Enhanced Set). Comparing the Enhanced PRS for coronary artery disease to Enhanced PRSs for known risk factors, the correlation with the LDL cholesterol PRS is 0.21, while the correlation with the body mass index PRS is 0.16. Despite generally low correlations, clustering of traits by PRS scores generates relationships consistent with known biology (Supplementary Figure 12). For example, type 2 diabetes clusters with body mass index (r = 0.31, Enhanced Set) and glycated haemoglobin (r = 0.31, Enhanced Set).

The relatively low between-PRS correlations suggest that multi-PRS prediction models could be useful for analyses of general mortality. Following previous work ^14,36^, we carried out a stepwise regression of time from first assessment to death from any cause, using all Standard Set disease and quantitative trait PRS scores (use of the Standard Set allowed UKB WBU to be used for training the model; see Supplementary Information). As an encouraging sanity check, we found the expected 2:1 relationship between PRS effect size on participants’ own mortality compared to that of their parents (Supplementary Figure 13). PRS scores for common diseases including coronary artery disease (hazard ratio per SD = 1.05, 95% CI 1.03-1.07) and ischaemic stroke (hazard ratio per SD = 1.06, 95% CI 1.04-1.09) were significant determinants of all-cause mortality, as was the PRS for body mass index (hazard ratio per SD = 1.06, 95% CI 1.04-1.08). The PRS for HDL cholesterol (hazard ratio per SD = 0.98, 95% CI 0.96-1.00) was a notable protective factor. The model including all the PRS risk factors in Supplementary Figure 13, trained in the WBU subgroup and tested in the Testing Subgroup, was a significantly better predictor of mortality than a model including age-at-first-assessment and sex only both for own mortality (change in Harrell’s C = 0.0041, 95% CI 0.0021-0.0059, p = 3.5×10^−5^) and for parental mortality (change in Harrell’s C = 0.0065, 95% CI 0.0059-0.0071, p = 2.1×10^−93^, where the model was adjusted for the participant’s age-at-first-assessment and for the parent’s sex). However, the amount of variation explained by the model was low (Royston’s ^37^ measure of explained variation = 1.8% for the PRS-only model on participants’ own mortality), suggesting the model is useful more for biological insight than direct prediction.

### Validation of the PRS algorithms in other cohorts

The previous sections have validated the PRS scores as powerful predictors of disease and quantitative traits in UK Biobank. To further validate the PRS algorithms (and implicitly the methodology that generated them), and to guard against UKB specificity of results, we examined their performance in other cohorts. We investigated cohort specificity by applying our PRS algorithms for 12 diseases to an external evaluation dataset. The 100,000 Genomes Project ^21,22^ (100KGP) is similar to UK Biobank in being UK-based (specifically, England-based), with linkage to the same UK electronic healthcare record system, but is different in being recruited either via genetic disorder probands or cancer diagnosis, and in being genetically assayed via whole-genome sequencing rather than array-based genotyping. We selected a subgroup of unrelated 100KGP individuals, excluding those with rare genetic or comorbid disorders (see Supplementary Methods). Despite the differences in cohort characteristics, we found predictive performances to be similar (Figure 7; Pearson r (logOR per SD, Enhanced PRS) = 0.948; Pearson r (logOR per SD, Standard PRS) = 0.971). The only disease with a significant difference in logOR per SD (at nominal 5% level) was atrial fibrillation, which had a higher performance in 100KGP than in UKB for both the Enhanced PRS (OR per SD (100KGP) = 2.18 (95% CI 2.01-2.35); OR per SD (UKB) = 1.93 (95% CI 1.86-1.99)) and the Standard PRS (OR per SD (100KGP) = 2.06 (95% CI 1.89-2.22); OR per SD (UKB) of 1.81 (95% CI 1.74-1.87)).

**Figure 7.**
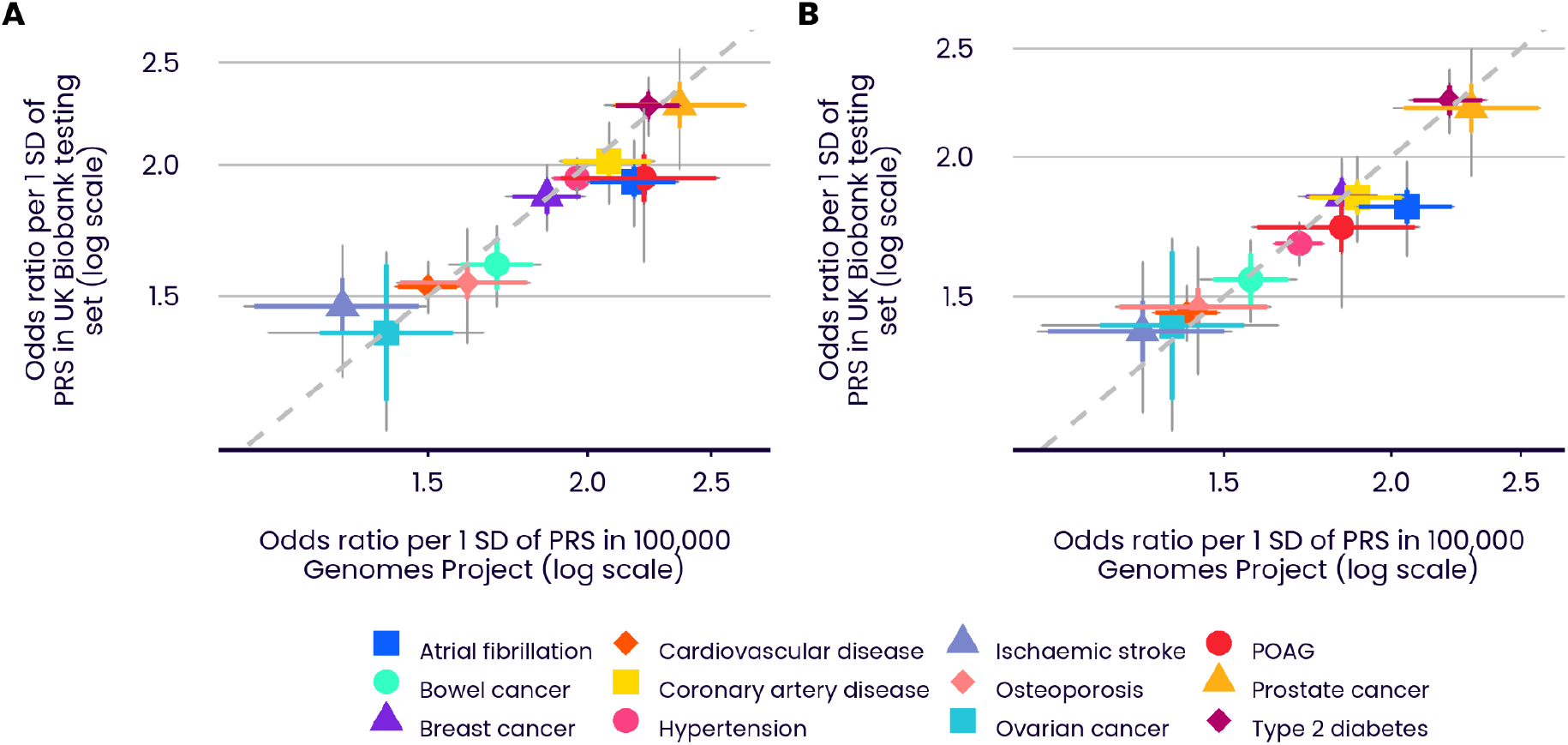
Comparative predictive performance in UK Biobank and 100,000 Genomes Project. Performance (OR per SD) across 12 diseases in the UK Biobank Testing Subgroup and selected individuals of European ancestry from the 100,000 Genomes Project (selected to be free of rare genetic and comorbid disorders). Coloured bars show the 95% CI of the OR per SD. Grey bars show the 95% CI of the change in logOR per SD between the two cohorts. Grey bars that intersect the 1:1 slope (dotted line) indicate non-significance (at 5% level) of the hypothesis that the performance in the two cohorts is different (true for all diseases except atrial fibrillation in both panels). **A** Enhanced PRSs. **B** Standard PRSs. Refer to Figure 1 for disease abbreviations.

We extended our cross-cohort comparisons to additional, primarily US-based, cohorts across multiple genetically inferred ancestries (Supplementary Figure 14 and Supplementary Tables 6, 7 and 8). There is a similar pattern between Enhanced and Standard PRS performance, but some traits (for example, hypertension, venous thromboembolic disease, and age-related macular degeneration) show considerable cross-cohort heterogeneity, underlining the role that cohort-specific factors can play in predictive performance, and reinforcing the need for a standardised comparison framework when evaluating predictive performance. Heterogeneity has also been observed in other cross-cohort comparison studies ^12^.

## Discussion

The availability of population-scale biobanks has generated unparalleled opportunities to create and evaluate genetic tools for predicting disease and quantitative traits in real-world settings ^12^. We have provided a platform for such research, by providing powerful PRS scores for 28 diseases and 25 quantitative traits in UK Biobank.

Validating PRS performance, or comparing the performance of different PRS algorithms, is challenging, because performance is context specific. The primary requirement is for a ‘level playing field’, to correct for cohort-specific and design-specific factors such as phenotype definition and other cohort characteristics ^12^. We have developed and made available a PRS evaluation tool to address this requirement. It enables robust and like-for-like comparisons of the predictive performance of different PRS scores in UKB. It should facilitate the ongoing development of PRS algorithms by the research community.

Our comparisons show the Enhanced PRS scores released in UKB to be more powerful, in European-ancestry and in non-European ancestry individuals, than almost all of those in a large comparator set of 81 previously released PRSs across these traits. The observation of similar performance in a separate UK cohort (the 100,000 Genomes Project) further validates the algorithms underlying the PRSs released in UKB. Performance across a set of non-UK cohorts remained generally powerful, but was more heterogeneous, likely resulting from confounding with cohort-specific differences, including phenotype definition. We anticipate the extension of multiple PRS Releases and evaluation tools to other cohorts, to enable effective cross-cohort analyses. We note that these tools will become especially valuable as PRS algorithms themselves move beyond simple linear combinations of variant weights, and towards other algorithmic forms with pre- and post-processing steps ^3^.

The availability of powerful PRSs for 53 traits on the same large set of extensively characterised individuals allowed a systematic study of PRS properties. PRS performance differs substantially across the diseases studied, presumably due, amongst other things, to differences in GWAS sample size ^38^ and genetic architecture across diseases (for example, ischemic stroke is a collection of multiple subtypes which have differing genetic risk factors ^39^, meaning that an algorithm that generates a single PRS for this compound phenotype will have reduced power). With a few unsurprising exceptions, within-individual pairwise correlations of PRS scores are low. The correlation between the Enhanced coronary artery disease (CAD) PRS and the Enhanced PRSs for known quantitative risk factors for CAD (e.g. LDL cholesterol: r = 0.21; body mass index: r = 0.14) are also not appreciable. Many PRSs show evidence for larger effect sizes for younger, compared to older, individuals in UKB.

One critical aspect is PRS performance in different ancestry groups ^19,20^. We developed our PRS evaluation tool with this in mind, maximising the non-European ancestry representation in the Testing Subgroup of UKB and reporting ancestry-specific results for all analyses with sufficient case data. We confirmed and quantified the widely-observed diminution of performance across ancestries, with average decrease in disease OR per SD of 9.1%, 14.3%, and 27.6%, respectively for South Asian, East Asian, and African ancestry individuals respectively, relative to the performance in European ancestry individuals. Clearly there is an urgent need for additional GWAS data in non-European ancestry individuals, through further studies, and where possible, release of summary statistics from existing studies, to improve PRS training data, and for improved PRS methodologies to further reduce performance differences across ancestries. We note that PRSs nonetheless have predictive power across all ancestries, and that the predictive power of the UKB Enhanced PRS in African-ancestry individuals for some diseases (e.g. type 2 diabetes or prostate cancer, with OR per SD of 1.48 (95% CI 1.40-1.56) and 1.56 (95% CI 1.38-1.76) respectively) are larger than those for European ancestry individuals for other diseases (e.g. cardiovascular disease or age-related macular degeneration, with OR per SD of 1.53 (95% CI 1.50-1.57) and 1.47 (95% CI 1.40-1.54), respectively). Further, differences in baseline risk in different ancestries can mean that, notwithstanding diminished PRS performance, high-PRS individuals in a particular non-European ancestry can be at higher levels of absolute risk than similarly high-PRS individuals of European ancestry (recall Figure 4, which shows much higher risk for type 2 diabetes for similar levels of PRS in South Asian, compared to European ancestry individuals in UKB). Discussion of the application of PRS in different groups should incorporate differences in disease specific performance, but also in baseline disease rates. One limitation of our cross-ancestry analysis is that it reflects the representation in UKB of these non-European groups, which will not necessarily be the same as that in other cohorts. Small numbers of cases for rarer diseases also precluded some comparisons.

Many health systems currently have active programmes to identify carriers of rare, high-penetrance, mutations which increase risk for common diseases, such as familial hypercholesterolemia (FH) for CAD, mutations in *ATM, BRCA1/2, CHEK2*, and *PALB2* for breast cancer, and Lynch syndrome for bowel cancer. As others have noted ^9,10^, it is now possible to identify a different set of individuals, with equivalent levels of risk to that of rare mutation carriers, where the risk is also genetic, but driven by the cumulative impact of large numbers of common variants. Continuing improvement in PRS methodology and training data will further increase the proportion of individuals in the population at levels of risk which would attract attention in health systems if due to rare variants. For example, with the UKB Enhanced CAD PRS we have shown that the top 8% of individuals have the same level of CAD risk as FH mutation carriers (comparing individuals in UKB not on statins).

It seems untenable in the long term to offer interventions or enhanced screening to one group of individuals at high risk because of genetics but not to another, just because the variants contributing to risk are different. This supports the case for an equivalence-of-risk principle, in which risk-based screening guidelines developed for the management of high-risk variant carriers ^29,30^ can be extended to cover individuals with equivalent risk based on their PRS. Further, the high-PRS individuals account for many fold more disease events ^9,10^, an effect which will also increase as the power of PRSs continues to improve. For example, in the FH example, the high PRS group is responsible for up to 30-fold more disease events compared to FH carriers. Increasing detection of FH carriers is, appropriately, a focus of many health systems (e.g. a key metric in the current 10-year plan for the UK NHS is to increase detection from current levels of 7% to at least 25%) ^40^. These results suggest that a parallel approach to detecting high-risk individuals via PRS could have an even greater impact on disease prevention.

The UK Biobank PRS Release provides well-validated PRS scores across multiple traits, and provides opportunities for subsequent research, but we expect that they will evolve in time, and will be improved upon. We have provided a comparative evaluation tool in the expectation that better PRS scores will be developed, as data and methodologies improve. We plan regular updates to the PRS Release, both to improve on performance and to expand the list of traits. In this way, we anticipate the UK Biobank PRS Release will provide an ongoing platform of powerful polygenic risk scores, to enable continuing research and clinical model development.

## Data Availability

The polygenic risk scores computed for each individual in UK Biobank have been made available to all approved UK Biobank researchers through the data showcase mechanism. The new GWAS data have been made available via https://zenodo.org/record/6631952.

https://github.com/Genomicsplc/ukb-pret

## Methods

Please refer to Supplementary Materials.

