## Supplementary information for "UK Biobank release and systematic evaluation of optimised polygenic risk scores for 53 diseases and quantitative traits"

### Data and code availability

The UK Biobank PRS Release is available via application to the UK Biobank's Research Access Platform. The Evaluation Tool is available as a command line tool within the Research Access Platform. Source code for the Evaluation Tool is available at https://github.com/Genomicsplc/ukb-pret. The GWAS summary statistics for 53 traits, performed in the WBU subgroup as additional training data for the Enhanced PRS Set, is available at https://zenodo.org/record/6631952.

### Supplementary methods

#### UK Biobank

A schematic workflow for the generation and standardised evaluation of the UK Biobank PRS Release is presented in Supplementary Figure 15.

The UK Biobank (UKB) is a UK based prospective cohort of ~500,000 individuals aged 40-69 at enrolment. Genotype data were generated using a custom Axiom genotyping array assaying 825,927 genetic variants, followed by genome-wide imputation. Phenotype information was assessed using a combination of a 2-day visit at UKB enrollment, questionnaires, and linkage with primary and secondary care electronic health records, including Hospital Episode Statistics and cancer registry data. See Bycroft et al [1] for further details.

After removal of exclusions and withdrawals, a subset of 337,151 UKB individuals, the White British Unrelated (WBU) subgroup, was defined as the intersection of two sample groups created by Bycroft et al [1]: the 'White British ancestry' group (UKB Data Field 22006) and the 'used in genetic principal components' group (UKB Data Field 22020), the latter being high quality samples that were filtered to avoid closely related individuals. This WBU subgroup (the 'training subgroup') was used to generate genome-wide association study (GWAS) summary statistics, which were then meta-analysed with external GWAS datasets to create the Enhanced PRS Set. 104,231 of the remaining individuals (the Testing Subgroup) were used for evaluation, including 82,346, 9,543, 9,478 and 2,864 individuals of European, South Asian, African, and East Asian ancestries, respectively (see below for details of genetic ancestry inference). There were too few individuals of Native American ancestry (N=390) to be included for evaluation.

All UKB individuals have given informed consent. Our research project (Project Application Number 9659) was approved by the UK Biobank according to their established access procedures [3], and legal and ethical approval is covered by the Research Tissue Bank approval obtained from the UK Biobank's governing Research Ethics Committee (REC 16/NW/0274), as recommended by the National Research Ethics Service.

#### Phenotypes, GWAS and meta-analysis

Phenotype variables for each trait within UK Biobank were created from a combination of Hospital Episode Statistics, Cancer Registry reports (where applicable) and self-report responses. Exclusion criteria resulted in some phenotypes being set to 'missing' for some individuals, for example, low density lipoprotein cholesterol levels were set to missing for individuals on statin medication. For details, see Supplementary Table 5. We carried out various *ad hoc* QC checks for errors and consistencies in UK Biobank data fields; these indicated only a small fraction of remaining individuals are affected.

GWAS summary statistics for each trait were generated by applying PLINK 2.0 to the WBU subgroup, applying a logistic regression model for disease traits, and a linear regression model for quantitative traits. Age at first assessment, genotyping chip, sex (for non-sex specific traits), and principal components (PCs) 1-10 (described by Bycroft et al [1]) were included as covariates in the models. UK Biobank GWAS sample size information is provided in Supplementary Tables 3 and 4.

In addition, a literature search for external GWAS summary statistic data was conducted for each trait (Supplementary Tables 1-4). Studies were excluded if they contained UKB data or if they were included in more recent published releases or meta-analyses.

To provide a common reference, and ensure consistency across studies, all GWAS summary statistics (both published and internally generated) were harmonised to a variant backbone, ensuring alignment to the Alt allele of the forward strand of Genome Reference Consortium Human Build 37 (GRCh37). Summary statistic imputation methodology [4] was deployed to fill in gaps in the variant backbone using 1000 Genomes Project data [5] as a reference panel for LD inference.

GWAS studies were meta-analysed via fixed-effect inverse variance meta-analysis, including a correction for sample overlap where required [5]. Data permitting, two meta-analyses were performed per trait; one including UK Biobank GWAS data to create the Enhanced PRS Set, and one excluding UK Biobank GWAS data to create the Standard PRS Set. For some quantitative traits, no suitable external GWAS data could be identified, and for these traits no Standard PRS was created (see Supplementary Table 2).

#### Genetic ancestry inference

Genome-wide genetic data for UK Biobank participants were used to estimate the proportion of each genome that could be ascribed to each one of five high-level ancestry groups: Sub-Saharan African (AFR), Native/Indigenous American (AMR), East Asian (EAS), European (EUR), and South Asian (SAS). To obtain these estimates, we first applied Principal Component Analysis (PCA) to a subset of common SNP genotypes in the 1000 Genomes reference dataset using standard methods [6]. To select SNPs for PCA, we intersected HapMap3 variants with a collection of well-imputed variants across UK Biobank, GSA, and OMNI arrays. We dropped SNPs with low call-rate (< 0.05) or MAF < 0.02 in 1000 Genomes, and then LD-pruned (using plink --indep 50 5 2) to an approximately independent set of around 185,000 SNPs across the autosomes. For PCA, the genotypes were mean-centred, but not standardised.

Centroid coordinates for ancestry groups in PC space (defined from the first four PC axes) were obtained from those 1000 Genomes Project [7] individuals belonging to population codes as follows (excluding populations that were found empirically to be highly admixed): AFR (ESN, GWD, LWK, MSL, YRI); AMR (MXL, PEL); EAS (CDX, CHB, CHS, JPT, KHV); EUR (CEU, FIN, GBR, IBS, TSI); SAS (BEB, GIH, ITU, PJL, STU). Genotype data from individuals outside the 1000 Genomes cohort were then projected onto the same PC axes to obtain their PC scores. The relationship between each individual's vector of PC scores and each centroid was determined via a softmax transformation (base=exp(3)) applied to the cosine similarity of the two vectors, to obtain the estimated ancestry proportions. The use of base=exp(3) was found empirically to result in a good reclassification of 1000 Genomes individuals to their correct ancestry group (data not shown). Individuals were assigned the superpopulation hard-call for which they presented the highest ancestry proportion, such that no individual was labelled as admixed.

#### Generation of UK Biobank PRS Release

The Standard PRS Set (also referred to as the "UKB-Free" set) of 28 diseases and 8 quantitative traits was generated from external GWAS data described in Supplementary Tables 1 and 2. Published comparator PRSs (see later) were not used in the construction of either the Standard or Enhanced PRS sets. The Enhanced PRS Set (also referred to as the "UKB-WBU" set) of 28 diseases and 25 quantitative traits was generated from external GWAS data plus UKB White British Unrelated (WBU) GWAS data described in Supplementary Tables 3 and 4. An exception was made for Crohn's disease (CD) and ulcerative colitis (UC). The external GWAS data for these two traits derive primarily from Immunochip arrays [8], which have high SNP density around signal genes and low density elsewhere. This, coupled with the relatively low case count in UKB WBU (2064 CD cases and 3914 UC cases), meant that the UKB contribution consisted mostly of between-signal noise, and the PRS with UKB data added had lower performance than without it. Because of these special circumstances, we reverted to the Standard PRS algorithm when adding CD and UC to the Enhanced PRS Set.

Genetic variants used to generate PRS weights were required to have an INFO score > 0.8 in UKB; have an INFO score > 0.8 in the GWAS meta-analysis dataset; have an INFO score > 0.7 in other key reference datasets available to Genomics plc; not display large differences in allele frequency between UKB genetically inferred ancestry groups (see above) and either Gnomad or 1000 Genomes Project (absolute allele frequency difference between Gnomad and UKB of less than 0.2 in any ancestry group, p>1e-12 and p>1e-10 for Gnomad and 1000 Genomes Project respectively in any ancestry group); and not display evidence of large departures from Hardy-Weinberg Equilibrium (p>1e-10) in any ancestry group. The variants also needed to have a definitive one-to-one mapping between Genome Builds 37 and 38. We excluded indels, the pseudoautosomal regions, and any variants with minor allele frequency (MAF) <0.05 in the 1000 Genomes Project dataset (for any ancestries used as LD reference panels in the PRS generation step).

PRS algorithms were built from trait-specific meta-analyses using a Bayesian approach (see Supplementary Tables 1-4), where appropriate combining data across multiple ancestries and related traits. Per-individual PRS values were calculated as the genome-wide sum of the per-variant posterior effect size multiplied by allele dosage.

Following the generation of a raw PRS value for an individual using the PRS weights derived above, a centering and standardisation step was applied in order to produce a corrected PRS value that could be interpreted as coming from a distribution that is of approximately of zero mean and unit variance for people occupying the same position in ‘ancestry space’ as the individual in question. First, the centred PRS was obtained following the method of Khera et al [9], by subtracting out the PRS value predicted from a linear regression of the PRS against the first four principal component scores, fitted in 1000 Genomes Project individuals [7]. Next, the genetic ancestry of each individual was inferred (see above). The centred PRS was then divided by the standard deviation of the PRS in the 1000 Genomes ancestry group with the closest match to the individual in question, to obtain a centred and variance-standardised PRS.

#### Comparator PRS Set

The PGS Catalog database (https://www.pgscatalog.org/) was searched (30th November 2021) to identify comparator PRS algorithms for any of our traits of interest (Supplementary Table 10). From this list we excluded PRS algorithms which had used any UKB GWAS data in the training stage (algorithms based on non-UKB GWAS data which had only used UKB data for the training of hyperparameters were not excluded). We also excluded algorithms which were designed for specific subtypes of the disease or trait, algorithms which were superseded by a more recent algorithm from the same research group, and duplicates. Where the same research group had uploaded multiple algorithms for the same trait to the PGS Catalog, each using different statistical methodologies or different parameters, we selected the one which the authors reported as having the best performance. Where several groups had reported PRS algorithms for a given trait which were all simple lists of GWAS-significant SNPs reported in the literature, we selected the most recent algorithm. We included algorithms which appeared to be similar to other algorithms, but which had been specifically optimised for non-European ethnicities or ancestries. Additional PRS algorithms were identified by regular monitoring of relevant publications, pre-print websites, social media and by conference attendance.

Comparator algorithms were excluded at the QC stage if there was insufficient SNP information to allow the algorithm to be implemented, or if >10% of the SNPs in the algorithm were not present in the imputed UKB dataset. Post-processing steps (PRS centering and standardisation) were applied to comparator PRS in an equivalent way to the UK Biobank PRS Release sets.

#### PRS performance evaluation

All three PRS sets - the Standard Set, the Enhanced Set, and the Comparator PRS Set - were evaluated in the same multi-ancestry Testing Subgroup in UKB (see above), as well as in other evaluation cohorts (Supplementary Table 6).

Additional QC was performed in UKB prior to PRS evaluation on each trait. Individuals without a reported age at first assessment were excluded. Individuals with conflicting case status vs date-of-diagnosis or incident status were excluded. Unless censored by death, a formal withdrawal from the project, or by a hard cutoff date of 1 March 2020, linkage to national HES data was assumed complete for all individuals up to 10 years from first assessment (12 years for age related macular degeneration), but then taken as censored after that time. All case events that occurred more than 10 years after first assessment (12 years for age related macular degeneration), or after the hard cutoff date of 1 March 2020, were treated as controls.

Performance evaluation in a given ancestry group required a minimum of 100 cases in that ancestry group for disease traits, and 100 samples in that ancestry group for quantitative traits. The AMR ancestry group was excluded for all traits due to low sample size. For evaluation in the 100,000 Genomes Project, a minimum of 75 cases was required. For an overview of sample sizes in the evaluation cohorts, see Supplementary Table 6.

PRS performance was evaluated based on several metrics. For disease traits, odds ratios (from logistic regression) are reported per standard deviation (SD) of PRS, and also by comparing the top 3%, 5% and 10% of the PRS distribution against the rest. Age at recruitment and sex were added as covariates in the logistic regression where possible, noting that age was not available for all cohorts, and sex could not be added as a covariate for analyses within a single sex. For disease traits in prospective cohorts, the hazard ratios per SD (from Cox regression, with age and sex as covariates where possible) are also reported. For quantitative traits, the regression coefficients (from linear regression, with age and sex as covariates where possible) on the raw trait scale and standardised trait scale are also reported. For disease traits, the area under the receiver operating characteristic curve (AUC) is also reported. Standard errors for AUC were found using an implementation of deLong's algorithm [10]. For quantitative traits, the variance explained ($r^2$) of the trait by the PRS is also reported. Standard errors for $r^2$ were found by Fisher's z-transformation. Standard errors for relative changes in odds ratios, AUC, and $r^2$, correcting for correlations induced by calculating PRSs in the same individuals, were found using a stratified (within-case/within-control) bootstrapping procedure.

#### Analyses of PRS properties

Age-specific hazard ratios were calculated by splitting the age-at-first-assessment into 10-year age bins, and then using Cox regression (adjusted for sex, where approriate) to estimate an incident hazard ratio within each age bin, allowing a maximum of 10 years of follow-up.

Whole exome sequencing data were available for 189,954 European-ancestry UKB participants. Carriers of deleterious mutations in either a breast cancer risk gene (loss-of-function mutations in *ATM*, *BRCA1*, *BRCA2* or *PALB2,* with classification of a mutation as loss-of-function based on a "high-confidence" scoring using the LOFTEE software [11]*,* or the *CHEK2* 1100delC mutation) or a familial hypercholesterolemia (FH) risk gene (pathogenic or likely-pathogenic mutations in *APOB*, *APOE*, *LDLR* or *PCSK9*, as classified by using the American College of Medical Genetics and Genomics (ACMG) guidelines [12], and extending the list provided by Fahed *et al* [13] by moving from previous 50k UKB exome dataset to the newer 200k UKB exome dataset) were identified. For each gene, the relevant disease (female breast cancer or coronary artery disease (CAD)) hazard ratio for carriers compared to non-carriers was estimated in the UKB Testing subset, and the upper percentile of the Enhanced PRS distribution for which the hazard ratio (HR) (compared to the 40-60$^{th}$ percentiles) matched the carrier HR was found. Individuals with a

PRS above this value can be thought of as having a broadly equivalent overall level of risk to that of mutation carriers. The CAD analyses were repeated in the subset of UKB participants for whom Primary Care prescribing data were available and who did not have a reported prescription for statin medication (other than prescriptions which began after the CAD diagnosis). Statin prescriptions were extracted from the UKB Primary Care data linkage dataset, using read_v2 codes bxd*, bxe*, bxg*, bxi*, bxj*, bxk* and bxl*, BNF codes 02.12.02.00.00 and 02.12.04.00.00, or via text searches for "atorvastatin", "cerivastatin", "ezetimibe", "fluvastatin", "pravastatin", "rosuvastatin", "simvastatin" and associated brand names (the LDL-lowering drug ezetimibe was included in the list, although in practice ezetimibe prescriptions were almost always seen in individuals also receiving a statin prescription). After excluding statins users, 164 of the 656 FH mutation carriers and 30,271 of 82,335 UKB Testing subset were included in the evaluation analyses (all European ancestry). Sample numbers are provided in Supplementary Table 11.

The pattern of age-specific risks associated with a high PRS was compared to that for mutation carriers using cumulative incidence plots. Analyses were repeated combining mutations across all of the genes relevant for that disease, to find the percentile of the PRS distribution for which the HR was equivalent to having a mutation in any of those genes. The HRs for mutation carriers and for PRS above this percentile were also calculated separately for cases diagnosed below ages 50, 60 or 70 years, to explore whether either risk factor has a stronger association with earlier-onset disease. The proportions of cases diagnosed below ages 50, 60 or 70 years who carried a rare mutation or who had a PRS above the risk-equivalent percentile were calculated. Confidence intervals for case proportions were calculated using the "exact" method in the binconf() function of the R Hmisc package. Finally, we used cumulative incidence plots to compare the disease risks between rare mutation carriers in the top and bottom 10% of the PRS distribution. Here we used the Standard PRS, in order to maximise the number of UKB samples with exome sequencing data available for the analyses.

Forwards-backwards stepwise regression was used to find the best-fitting linear combination of PRS scores for predicting all-cause mortality. The WBU subset of UKB was used to train a Cox proportional-hazards model, with years from first assessment until participant's death or censoring as the time variable, searching over all 36 Standard PRS scores (28 diseases plus 8 quantitative traits). Model selection was based on the Akaike information criterion, with significance threshold 0.05. A second forwards-backwards stepwise regression was then applied to parental mortality data. Data for the mother and father of each participant (excluding those who reported that they had been adopted as a child) were taken as two separate observations, using their offspring's PRS scores as predictors. For each, their follow-up was from their birth until either their age at their offspring's UKB assessment or their age at death, as reported by the participant at their first UKB assessment. Ideally, parental follow-up would begin at the parent's age when the participant was born, to avoid immortal time bias [14], but this age is not available for parents who died before UKB assessment. We therefore assumed a minimal effect of immortal time bias on parental outcomes, and this assumption is borne out by the observed 2:1 ratio between PRS effect sizes in the "own mortality" and "parents' mortality" analyses (Supplementary Figure 13). Traits which were selected by both the "own mortality" and "parents' mortality" stepwise regression analyses were then entered into a final training step, in which the PRS scores for these traits were entered as covariates, together with participants' age at first recruitment and sex (of the participant, mother or father, as applicable) as additional covariates, into separate "own mortality" and "parents' mortality" Cox proportional-hazards models. Coefficients from these final models were then fixed and evaluated in the remaining UKB Testing Subgroup. Differences in Harrell's C statistic, in models with and without PRS covariates, were tested via a z-test on the difference, using an estimated variance that accounts for covariance in C statistics and derived from an infinitesimal jackknife estimator [15], and 95% confidence intervals for the difference in C-statistics were estimated using 1,000 bootstrap samples.

### Additional cohorts

**100,000 Genomes Project (100KGP)**. The 100,00 Genomes Project, run by Genomics England, consists of more than 100,000 whole-genomes sequences, with electronic health record data, from ~85,000 NHS patients in England affected by a rare disease or cancer, as well as the parents of some of the rare disease patients [16]. Recruitment of participants to the 100,000 Genomes Project was completed in 2018. All participants gave informed consent.

For our PRS performance evaluations, rare disease affected participants were excluded and from related pairs (up to 3rd degree), one randomly selected individual was included (KING[2] threshold = 0.0442), resulting in a sample of 40,001 for analysis. Germline Whole Genome Sequence (WGS) data were filtered to the GWAS analysis variant list. To fill in gaps in the variant list, genotypes were phased (Eagle v2.4.1 [17]) and imputed (Minimac3 [18]) using the 1000 Genomes Project reference panel. This set of common variant genotypes has been made available as a shared resource in the Genomics England research environment.

Hospital Episode Statistics (HES) and cancer registry data were used to identify disease cases and controls using ICD-10 codes (see Supplementary Table 5). For cancer traits, individuals that received a differing cancer diagnosis prior to, or contemporaneously to, the main diagnosis were removed to ensure the cancer diagnosis was not a result of comorbid metastasis. Controls were selected exclusively from the participant pool of the unaffected rare disease relatives. For non-cancer traits, individuals that received a cancer diagnosis prior to the main diagnosis were removed to avoid disease development due to cancer treatment effects. Controls were selected from both rare disease arm (unaffected relatives) and cancer arm. 12 disease traits were evaluated (for sample size information, see Supplementary Table 6).

To enable fair PRS performance comparisons of 100KGP with UKB and other evaluation cohorts, software was imported into the 100KGP research environment to apply the same approach to ancestry estimation, PRS calculation and ancestry centering and variance normalisation.

**The Atherosclerosis Risk in Communities (ARIC) Study**. ARIC comprises over 15,000 adults from predominantly 2 study-defined racial/ethnic groups ("Black" and "White"), from defined populations in 4 sites in the USA, aged 45–64 years when recruited between 1987 and 1989 [19]. Participants were extensively examined at baseline, and have had continuing follow-up via annual phone calls. Genotype and phenotype data for ARIC were obtained from dbGaP accession id phs000090.v3.p1. All participants gave informed consent. Our use of these data was approved by the Western Institutional Review Board (Study Number 1264897, IRB Tracking Number 20192201).

**Discovery, Biology, and Risk of Inherited Variants in Breast Cancer (DRIVE)**. Discovery, Biology, and Risk of Inherited Variants in Breast Cancer (DRIVE) was a project funded in 2010 as part of the National Cancer Institute's Genetic Associations and Mechanisms in Oncology (GAME-ON) initiative (https://epi.grants.cancer.gov/gameon/). The DRIVE genotype and phenotype data were obtained from dbGAP accession id phs001265.v1.p1, which includes genotype data from 60,015 breast cancer cases and controls drawn from 17 studies and genotyped using the OnocArray chip [20]. All participants gave informed consent. Our use of these data was approved by the Western Institutional Review Board (Study Number 1264897, IRB Tracking Number 20192201). The majority of the samples are from European-ancestry women, and are a subset of those represented in the BCAC summary statistics [21] which were used in the training of our PRS. We therefore restricted our analysis to the 1,000 women with inferred African ancestry who had not been included in the BCAC European GWAS.

**Electronic Medical Records and Genomics (eMERGE)**. The Electronic Medical Records and Genomics (eMERGE) Network [22] is an NHGRI-funded consortium of ten participating sites (Cincinnati Children's Hospital Medical Center/Boston Children's Hospital, Children's Hospital of Philadelphia, Essentia Institute of Rural Health, Marshfield Clinic Research Foundation and Pennsylvania State University, Geisinger Clinic, Group Health Cooperative/University of Washington, Mayo Clinic, Icahn School of Medicine at Mount Sinai, Northwestern University, Vanderbilt University Medical Center). The goal of eMERGE is to conduct genome-wide association studies in approximately 55,000 individuals using EMR-derived phenotypes and DNA from linked Biorepositories (https://emerge-network.org/emerge-sites/). All participants gave informed consent. Our use of these data, under dbGaP accession phs000888.v1.p1, was approved by the Western Institutional Review Board (Study Number 1264897, IRB Tracking Number 20192201). We have split eMERGE into 6 cohorts (labelled eMERGE-1, eMERGE-2, etc) that reflect the participants' dbGaP research use permission codes (HMB-PUB-GSO, HMB-GSO, GRU, GRU-IRB-PUB, HMB and DS-DEM respectively).

**The Charles Bronfman Institute for Personalized Medicine (IPM) BioMe BioBank**. IPM is a consented, EMR-linked medical care setting biorepository of the Mount Sinai Medical Center drawing from a population of over 70,000 inpatients and 800,000 outpatient visits annually [23]. Genotype and phenotype data were obtained from dbGaP accession id phs000388.v1.p1. All participants gave informed consent. Our use of these data was approved by the Western Institutional Review Board (Study Number 1264897, IRB Tracking Number 20192201).

**Jackson Heart Study (JHS).** The Jackson Heart Study (JHS) is a community-based observational study comprising 5,301 African-American participants residing in Jackson,

Mississippi metropolitan statistical area. Approximately 30% of the participants are also enrolled in the ARIC study. A subset of 2,702 participants were genotyped for the Candidate gene Association Resource (CARe) using the Affymetrix 6.0 Array [24]. Genotype and phenotype data were obtained from dbGaP accession id phs000499.v3.p1. All participants gave informed consent. Our use of these data was approved by the Western Institutional Review Board (Study Number 1264897, IRB Tracking Number 20192201).

**Multi-Ethnic Cohort (MEC)**. The Multi-Ethnic Cohort is a population-based prospective cohort study of over 215,000 men and women living in Hawaii and California (45-75 yrs at baseline, collected from 1993-1996). The cohort includes large representations of older adults for five US racial/ethnic groups (Japanese Americans, African Americans, European Americans, Latinos and Native Hawaiians) [25]. A subset of ~28,000, with associated data on cardiometabolic traits, were selected for genotyping on the Multi-Ethnic Genotyping Array (MEGA) as part of the Population Architecture using Genomics and Epidemiology (PAGE) study [26] (dbGaP accession phs000220.v2.p2). A separate set of breast cancer cases and age- and ethnicity-matched controls were selected for genotyping on Human660W-Quad and Human-1M arrays as part of a separate study (dbGaP accession phs000517.v3.p1). All participants gave informed consent. Our use of these data was approved by the Western Institutional Review Board (Study Number 1264897, IRB Tracking Number 20192201).

**The Multi-Ethnic Study of Atherosclerosis (MESA)**. MESA comprises over 6,000 adults from 4 study-defined racial/ethnic groups ("African American", "Chinese American", "Hispanic", and "White/Caucasian"), recruited primarily via phone call invitation to 6 sites in the USA, aged 45–84 years and free of cardiovascular disease when recruited between 2000 and 2002 [27,28]. Participants were extensively examined at baseline, and have had continuing follow-up via annual phone calls. Genotype and phenotype data for MESA were obtained from dbGaP accession ids phs000420.v6.p3 and phs000209.v13.p3 respectively. All participants gave informed consent. Our use of these data was approved by the Western Institutional Review Board (Study Number 1264897, IRB Tracking Number 20192201).

**The Omics in Latinos (OLA) component of the Hispanic Community Health Study / Study of Latinos Project**. The Hispanic Community Health Study / Study of Latinos Project is a multi-center epidemiologic study in Hispanic/Latino populations to assess the role of acculturation in the prevalence and development of disease. Genotype and phenotype data for OLA were obtained from dbGaP accession id phs000880.v1.p1. All participants gave informed consent. Our use of these data was approved by the Western Institutional Review Board (Study Number 1264897, IRB Tracking Number 20192201).

**GWAS of Breast Cancer in the African Diaspora (ROOT study)**. The ROOT consortium study is a case-control GWAS of breast cancer in women of African ancestry, including Africans living in Nigeria, African Americans and African Barbadians [29]. The dbGaP dataset (phs000383.v1.p1) includes genotypes for 3,766 women. After excluding close relatives, those for whom we were not able to calculate a PRS, and 23 women of inferred European or admixed ancestry, the analyses included 1,643 breast cancer cases and 2,049 controls, all of inferred African ancestry. All participants gave informed consent. Our use of these data was approved by the Western Institutional Review Board (Study Number 1264897, IRB

Tracking Number 20192201). The African American component of the ROOT dataset includes up to 220 cases and 430 controls from the Southern Community Cohort Study (SCCS). The BCAC African-American/Afro-Caribbean iCOGS study [30] for which summary statistics were included in the our PRS training also includes 679 cases and 680 cases from the SCCS. It was not possible to remove either the SCCS data from the summary statistics, or the overlapping SCCS women from the ROOT data, but it is unlikely that this will have introduced bias, since the SCCS cases only represent 0.4% of the training summary statistics.

### Acknowledgements

UK Biobank (UKB)
This research has been conducted using the UK Biobank Resource under Application Number 9659. We thank Alan Young, Lucy Bukitt-Gray, Caroline Clark and Oliver Gray for their help in making the UK Biobank PRS Release available.

100,000 Genomes Project
This research was made possible through access to the data and findings generated by the 100,000 Genomes Project. The 100,000 Genomes Project is managed by Genomics England Limited (a wholly owned company of the Department of Health and Social Care). The 100,000 Genomes Project is funded by the National Institute for Health Research and NHS England. The Wellcome Trust, Cancer Research UK and the Medical Research Council have also funded research infrastructure. The 100,000 Genomes Project uses data provided by patients and collected by the National Health Service as part of their care and support. Our use of the data was approved by Genomics England's Access Review Committee under application reference AR88.

Atherosclerosis Risk in Communities (ARIC)
The Atherosclerosis Risk in Communities Study is carried out as a collaborative study supported by National Heart, Lung, and Blood Institute contracts (HHSN268201100005C, HHSN268201100006C, HHSN268201100007C, HHSN268201100008C, HHSN268201100009C, HHSN268201100010C, HHSN268201100011C, HHSN268201100012C, HHSN268201700001I, HHSN268201700002I, HHSN268201700003I, HHSN268201700004I, and HHSN268201700005I). The authors thank the staff and participants of the ARIC study for their important contributions. Funding for GENEVA was provided by National Human Genome Research Institute grant U01HG004402 (E. Boerwinkle).
The datasets used for the analyses described in this manuscript were obtained from dbGaP at http://www.ncbi.nlm.nih.gov/projects/gap/cgi-bin/study.cgi?study_id=phs000090.v7.p1

Discovery, Biology, and Risk of Inherited Variants in Breast Cancer (DRIVE)
OncoArray genotyping and phenotype data harmonization for the Discovery, Biology, and Risk of Inherited Variants in Breast Cancer (DRIVE) breast-cancer case control samples was supported by X01 HG007491 and U19 CA148065 and by Cancer Research UK (C1287/A16563). Genotyping was conducted by the Center for Inherited Disease Research (CIDR), Centre for Cancer Genetic Epidemiology, University of Cambridge, and the National Cancer Institute. The following studies contributed germline DNA from breast cancer cases and controls: the Two Sister Study (2SISTER), Breast Oncology Galicia Network (BREOGAN), Copenhagen General Population Study (CGPS), Cancer Prevention Study 2 (CPSII), The European Prospective Investigation into Cancer and Nutrition (EPIC), Melbourne Collaborative Cohort Study (MCCS), Multiethnic Cohort (MEC), Nashville Breast Health Study (NBHS), Nurses Health Study (NHS), Nurses Health Study 2 (NHS2), Polish Breast Cancer Study (PBCS), Prostate Lung Colorectal and Ovarian Cancer Screening Trial

(PLCO), Studies of Epidemiology and Risk Factors in Cancer Heredity (SEARCH), The Sister Study (SISTER), Swedish Mammographic Cohort (SMC), Women of African Ancestry Breast Cancer Study (WAABCS), Women's Health Initiative (WHI).
The datasets used for the analyses described in this manuscript were obtained from dbGaP at http://www.ncbi.nlm.nih.gov/projects/gap/cgi-bin/study.cgi?study_id=phs001265.v1.p1.

Electronic Medical Records and Genomics (eMERGE) Network
Cincinnati Children's Hospital Medical Center (CCHMC) – Acknowledgement Text: CCHMC is a participating pediatric institution for Phase III of the eMERGE network, a national consortium selected to expand best practices and knowledge in effective implementation of genomic medicine to pursue a broad-based program sufficiently large to define health outcomes associated with rare variants in ~100 clinically relevant genes. CCHMC Principal Investigators (PIs) have contributed sequencing data generated from the following cohorts: (1) Better Outcomes for Children (BOfC). Since January, 2011, the Cincinnati Biobank has managed the sample repository developed through the BOfC protocol (PI: John Harley), an institution-wide broad based consent project to utilize remnant clinical samples for biomedical research from participants consented at registration. This project is made possible by institutional resources. To date, over 261,000 participants have consented to BOfC and DNA samples are stored from more than 84,000 unique patients. Through an IRB approved protocol led by Dr. Bahram Namjou (2015-7778), 2,800 samples were selected for sequencing on the eMERGE sequencing panel representing >15 primary phenotypes including Arrhythmia, Asthma, Cardiomyopathy, Chronic kidney disease, Ehlers-Danlos Syndrome, Hyperlipidemia, Autistic behavior, and Tuberous Sclerosis 1. This project is made possible by the support of U01HG008666 (PI: John Harley). (2) Return of eMERGE III Genomic Results. Through an IRB approved protocol led by Dr. Melanie Myers (2016-3361), 200 adolescent patients and their parents were consented to examine (1) their choices about results to be returned on the eMERGE sequencing panel, (2) their responses to learning negative genetic test results, and (3) the parents' responses after learning their children's positive results. All 200 participants provided blood samples. Extracted DNA samples were sequenced on the eMERGE sequencing panel. Results are to be returned to participants. This project is made possible by the support of U01HG008666 (PI: John Harley). Patients of interest were identified using anthropometric measurements, clinical data and ICD codes extracted from the EPIC electronic medical record (EMR). The extraction of data from the EMR into the de-identified data warehouse, i2b2, was made possible by institutional resources and UL1RR026314/UL1TR001425, the Cincinnati Center for Clinical and Translational Sciences and Training Grant (PI: James Heubi). Children's Hospital of Philadelphia (CHOP) Center for Applied Genomics, The Children's Hospital of Philadelphia Samples and associated genomic and phenotype data used in this study were provided by the Center for Applied Genomics at the Children's Hospital of Philadelphia (CHOP). Support for genotyping was provided by an Institutional Development Award from CHOP. Support for sequencing was provided by the National Institutes of Health through an award from the National Human Genome Research Institute's Electronic Medical Records and Genomics (eMERGE) program (U01HG008684). Columbia University Samples and data used in this study were provided by the Center for Glomerular Diseases at Columbia University, the Columbia Transplant Programs, the DataBase Shared Resource at the Herbert Irving Comprehensive Cancer Center, and the Institute for Genomic Medicine at Columbia

University. Funding support for the Columbia eMERGE III research study was provided by a U01 grant from the National Human Genome Research Institute (U01HG008680; PIs – Chunhua Weng, PhD; George Hripcsak, MD; Ali Gharavi, MD). Geisinger Funding for the MyCode® sample and data collection was provided by grants from Commonwealth of Pennsylvania, the Clinic Research Fund of Geisinger Clinic, and the Regeneron Genetics Center. Partners Health Care (Harvard University) Samples and data used in this study were provided by the Partners Health Care Biobank (https://biobank.partners.org/). Funding support for the Partners Biobank was provided by Partners Health Care and Partners Personalized Medicine. Assistance with phenotype harmonization was provided by the eMERGE Coordinating Center (Grant number U01HG04603). Additional support was provided by the NIH, NHGRI eMERGE Network (U01HG 5U01HG008685-03). Funding support for genotyping, which was performed at the Translational Genomics Core, Partners Personalized Medicine and funded by Partners Personalized Medicine. Assistance with phenotype harmonization and genotype data cleaning was provided by the eMERGE Administrative Coordinating Center (U01HG004603) and the National Center for Biotechnology Information (NCBI). The datasets used for the analyses described in this manuscript were obtained from dbGaP at http://www.ncbi.nlm.nih.gov/gap through dbGaP accession number; phs000944.v1.p1. Kaiser Washington/University of Washington Funding support for Alzheimer's Disease Patient Registry (ADPR) and Adult Changes in Thought (ACT) study was provided by a U01 from the National Institute on Aging (Eric B. Larson, PI, U01AG006781). A gift from the 3M Corporation was used to expand the ACT cohort. DNA aliquots sufficient for GWAS from ADPR Probable AD cases, who had been enrolled in Genetic Differences in Alzheimer's Cases and Controls (Walter Kukull, PI, R01 AG007584) and obtained under that grant, were made available to eMERGE without charge. Funding support for genotyping, which was performed at Johns Hopkins University, was provided by the NIH (U01HG004438). Genome-wide association analyses were supported through a Cooperative Agreement from the National Human Genome Research Institute, U01HG004610 (Eric B. Larson, PI). Assistance with phenotype harmonization and genotype data cleaning was provided by the eMERGE Administrative Coordinating Center (U01HG004603) and the National Center for Biotechnology Information (NCBI). The datasets used for the analyses described in this manuscript were obtained from dbGaP at http://www.ncbi.nlm.nih.gov/gap through dbGaP accession number phs000234.v1.p1. Mayo Clinic Samples and associated genotype and phenotype data used in this study were provided by the Mayo Clinic. Funding support for the Mayo Clinic was provided through a cooperative agreement with the National Human Genome Research Institute (NHGRI), Grant #: UOIHG004599, UOIHG006379; and the Mayo Center for Individualized Medicine. Funding support for sequencing, which was performed at The Baylor Human Genomics Sequencing Center, was provided by the NIH. Assistance with phenotype harmonization and genotype data cleaning was provided by the eMERGE Administrative Coordinating Center and the National Center for Biotechnology Information (NCBI). Northwestern University Samples and data used in this study were obtained from patients of Northwestern Medicine, Chicago, IL, who were recruited for the eMERGE II Pharmacogenomics Study and the eMERGE III Your Genes and Your Health Study. The Pharmacogenomics Study, a supplement to the Northwestern eMERGE II Project (U01HG006388) and the Your Genes and Your Health Study (U01HG008673) were funded through the NIH, NHGRI eMERGE Network. Vanderbilt University Funding support for the Vanderbilt Genome-Electronic

Records (VGER) project was provided through a cooperative agreement (U01HG008672) with the National Human Genome Research Institute (NHGRI) with additional funding from the National Institute of General Medical Sciences (NIGMS). The dataset(s) used for the analyses described were obtained from Vanderbilt University Medical Center. Assistance with phenotype harmonization and genotype data cleaning was provided by the eMERGE Administrative Coordinating Center (U01HG004603) and the National Center for Biotechnology Information (NCBI). The datasets used for the analyses described in this manuscript were obtained from dbGaP at http://www.ncbi.nlm.nih.gov/gap through dbGaP accession number phs000188.v1.p1.
The datasets used for the analyses in this manuscript were obtained from dbGaP through dbGaP accession study number phs001584.v2.p2.

Charles R. Bronfman Institute for Personalized Medicine (IPM)
The datasets used for the analyses in this manuscript were obtained from dbGaP through dbGaP accession study number phs000388.v1.p1.

Jackson Heart Study (JHS)
The Jackson Heart Study is supported and conducted in collaboration with Jackson State University (HHSN268201300049C and HHSN268201300050C), Tougaloo College (HHSN268201300048C), and the University of Mississippi Medical Center (HHSN268201300046C and HHSN268201300047C) contracts from the National Heart, Lung, and Blood Institute (NHLBI) and the National Institute for Minority Health and Health Disparities (NIMHD). This manuscript was not prepared in collaboration with JHS investigators and does not necessarily reflect the opinions or views of JHS, or the NHLBI. Funding for CARe genotyping was provided by NHLBI Contract N01-HC-65226.
The datasets used for the analyses in this manuscript were obtained from dbGaP through dbGaP accession study number phs000499.v4.p2.

Multiethnic Cohort (MEC)
The Multiethnic Cohort and the genotyping in this study were funded by grants from the National Institute of Health (CA63464, CA54281, CA098758, CA132839 and HG005922) and the Department of Defense Breast Cancer Research Program (W81XWH-08-1-0383). The datasets used for the analyses described in this manuscript were obtained from dbGaP at http://www.ncbi.nlm.nih.gov/projects/gap/cgi-bin/study.cgi?study_id=phs000517.v3.p1. Funding support for the PAGE Multiethnic Cohort study was provided through the National Cancer Institute (R37CA54281, R01CA6364, P01CA33619, U01CA136792, and U01CA98758) and the National Human Genome Research Institute (U01HG004802). Assistance with phenotype harmonization, SNP selection, data cleaning, meta-analyses, data management and dissemination, and general study coordination, was provided by the PAGE Coordinating Center (U01HG004801-01).
The datasets used for the analyses described in this manuscript were obtained from dbGaP at http://www.ncbi.nlm.nih.gov/projects/gap/cgi-bin/study.cgi?study_id=phs000220.v1.p1

Multi-Ethnic Study of Atherosclerosis (MESA)
MESA and the MESA SHARe project are conducted and supported by the National Heart, Lung, and Blood Institute (NHLBI) in collaboration with MESA investigators. Support for

MESA is provided by contracts N01-HC95159, N01-HC-95160, N01-HC-95161, N01-HC-95162, N01-HC-95163, N01-HC-95164, N01-HC-95165, N01-HC95166, N01-HC-95167, N01-HC-95168, N01-HC-95169, UL1-RR-025005, and UL1-TR-000040. Funding for SHARe genotyping was provided by NHLBI Contract N02-HL-64278. Genotyping was performed at Affymetrix (Santa Clara, California, USA) and the Broad Institute of Harvard and MIT (Boston, Massachusetts, USA) using the Affymetrix Genome-Wide Human SNP Array 6.0. A full list of participating MESA investigators and institutions can be found at http://www.mesa-nhlbi.org.
The datasets used for the analyses in this manuscript were obtained from dbGaP through dbGaP accession study number phs000420.v6.p3.

The Omics in Latinos (OLA) component of the Hispanic Community Health Study /Study of Latinos (HCHS/SOL)
The Hispanic Community Health Study/Study of Latinos is a collaborative study supported by contracts from the National Heart, Lung, and Blood Institute (NHLBI) to the University of North Carolina (HHSN268201300001I / N01-HC-65233), University of Miami (HHSN268201300004I / N01-HC-65234), Albert Einstein College of Medicine (HHSN268201300002I / N01-HC-65235), University of Illinois at Chicago – HHSN268201300003I / N01-HC-65236 Northwestern Univ), and San Diego State University (HHSN268201300005I / N01-HC-65237). The following Institutes/ Centers/ Offices have contributed to the HCHS/SOL through a transfer of funds to the NHLBI: National Institute on Minority Health and Health Disparities, National Institute on Deafness and Other Communication Disorders, National Institute of Dental and Craniofacial Research, National Institute of Diabetes and Digestive and Kidney Diseases, National Institute of Neurological Disorders and Stroke, NIH Institution-Office of Dietary Supplements. The Genetic Analysis Center at the University of Washington was supported by NHLBI and NIDCR contracts (HHSN268201300005C, AM03 and MOD03). A complete list of staff and investigators is provided by Sorlie et al in Ann Epidemiol. 2010 Aug;20: 642-649 and is also available on the study website http://www.cscc.unc.edu/hchs/.
The datasets used for the analyses described in this manuscript were obtained from dbGaP at http://www.ncbi.nlm.nih.gov/projects/gap/cgi-bin/study.cgi?study_id=phs000810.v1.p1 and http://www.ncbi.nlm.nih.gov/projects/gap/cgi-bin/study.cgi?study_id=phs000880.v1.p1

ROOT
The GWAS of Breast Cancer in the African Diaspora is conducted by the University of Chicago and supported by the National Cancer Institute (R01 CA142996-02). This manuscript was not prepared in collaboration with investigators of the GWAS of Breast Cancer in the African Diaspora and does not necessarily reflect the opinions or views of University of Chicago, or NCI.
The datasets used for the analyses described in this manuscript were obtained from dbGaP at http://www.ncbi.nlm.nih.gov/projects/gap/cgi-bin/study.cgi?study_id=phs000383.v1.p1

Genotype and phenotype data for the Whole Genome Association Study of Systemic Lupus Erythematosus (dbGaP accession number phs000122.v1.p1) were provided by Timothy W. Behrens, Peter K. Gregersen, Lindsey Criswell, and Susan Manzi. Funding support for the original study was provided by Genentech, the National Institutes of Health, and other sources as detailed in Hom G, et al. Association of systemic lupus erythematosus with C8orf13-BLK and ITGAM-ITGAX. N Engl J Med. 2008. 358(9):900-9. The datasets used for the analyses in this manuscript were obtained from dbGaP through dbGaP accession study number phs000122.v1.p1.

Funding for the Collaborative Association Study of Psoriasis was provided by the National Institutes of Health, the Foundation for the National Institutes of Health, and the National Psoriasis Foundation. Support for genotyping of samples was provided through the Genetic Association Information Network (GAIN). The dataset(s) used for the analyses described in this manuscript were obtained from the database of Genotypes and Phenotypes (dbGaP) found at http://www.ncbi.nlm.nih.gov/gap through dbGaP accession number phs000019. Samples and associated phenotype data for the Collaborative Association Study of Psoriasis were provided by Drs. James T Elder (University of Michigan, Ann Arbor, MI), Gerald G Krueger (University of Utah, Salt Lake City, UT), Anne Bowcock (Washington University, St. Louis, MO) and Gonçalo R Abecasis (University of Michigan, Ann Arbor, MI). For a description of the dataset, phenotypes, genotype data and quality control procedures see Nair et al (2009) Nature Genetics 41:200-204. The datasets used for the analyses in this manuscript were obtained from dbGaP through dbGaP accession study number phs000019.v1.p1.

We are grateful to the International Multiple Sclerosis Genetics Consortium; Wellcome Trust Case Control Consortium 2, Sawcer S, Hellenthal G, Pirinen M, Spencer CC, Patsopoulos NA, Moutsianas L, Dilthey A, Su Z, Freeman C, Hunt SE, Edkins S, Gray E, Booth DR, Potter SC, Goris A, Band G, Oturai AB, Strange A, Saarela J, Bellenguez C, Fontaine B, Gillman M, Hemmer B, Gwilliam R, Zipp F, Jayakumar A, Martin R, Leslie S, Hawkins S, Giannoulatou E, D'alfonso S, Blackburn H, Martinelli Boneschi F, Liddle J, Harbo HF, Perez ML, Spurkland A, Waller MJ, Mycko MP, Ricketts M, Comabella M, Hammond N, Kockum I, McCann OT, Ban M, Whittaker P, Kemppinen A, Weston P, Hawkins C, Widaa S, Zajicek J, Dronov S, Robertson N, Bumpstead SJ, Barcellos LF, Ravindrarajah R, Abraham R, Alfredsson L, Ardlie K, Aubin C, Baker A, Baker K, Baranzini SE, Bergamaschi L, Bergamaschi R, Bernstein A, Berthele A, Boggild M, Bradfield JP, Brassat D, Broadley SA, Buck D, Butzkueven H, Capra R, Carroll WM, Cavalla P, Celius EG, Cepok S, Chiavacci R, Clerget-Darpoux F, Clysters K, Comi G, Cossburn M, Cournu-Rebeix I, Cox MB, Cozen W, Cree BA, Cross AH, Cusi D, Daly MJ, Davis E, de Bakker PI, Debouverie M, D'hooghe MB, Dixon K, Dobosi R, Dubois B, Ellinghaus D, Elovaara I, Esposito F, Fontenille C, Foote S, Franke A, Galimberti D, Ghezzi A, Glessner J, Gomez R, Gout O, Graham C, Grant SF, Guerini FR, Hakonarson H, Hall P, Hamsten A, Hartung HP, Heard RN, Heath S, Hobart J, Hoshi M, Infante-Duarte C, Ingram G, Ingram W, Islam T, Jagodic M, Kabesch M, Kermode AG, Kilpatrick TJ, Kim C, Klopp N, Koivisto K, Larsson M, Lathrop M, Lechner-Scott JS, Leone MA, Leppä V, Liljedahl U, Bomfim IL, Lincoln RR, Link J, Liu J, Lorentzen AR, Lupoli S, Macciardi F, Mack T, Marriott M, Martinelli V, Mason D, McCauley JL, Mentch F, Mero IL, Mihalova T, Montalban X, Mottershead J, Myhr KM, Naldi P, Ollier W, Page A, Palotie A, Pelletier J, Piccio L, Pickersgill T, Piehl F, Pobywajlo S, Quach HL, Ramsay PP, Reunanen M, Reynolds R, Rioux JD, Rodegher M, Roesner S, Rubio JP, Rückert IM, Salvetti M, Salvi E, Santaniello A, Schaefer CA, Schreiber S, Schulze C, Scott RJ, Sellebjerg F, Selmaj KW, Sexton D, Shen L, Simms-Acuna B, Skidmore S, Sleiman PM, Smestad C, Sørensen PS,

Søndergaard HB, Stankovich J, Strange RC, Sulonen AM, Sundqvist E, Syvänen AC, Taddeo F, Taylor B, Blackwell JM, Tienari P, Bramon E, Tourbah A, Brown MA, Tronczynska E, Casas JP, Tubridy N, Corvin A, Vickery J, Jankowski J, Villoslada P, Markus HS, Wang K, Mathew CG, Wason J, Palmer CN, Wichmann HE, Plomin R, Willoughby E, Rautanen A, Winkelmann J, Wittig M, Trembath RC, Yaouanq J, Viswanathan AC, Zhang H, Wood NW, Zuvich R, Deloukas P, Langford C, Duncanson A, Oksenberg JR, Pericak-Vance MA, Haines JL, Olsson T, Hillert J, Ivinson AJ, De Jager PL, Peltonen L, Stewart GJ, Hafler DA, Hauser SL, McVean G, Donnelly P, and Compston A for providing data from their paper "Genetic risk and a primary role for cell-mediated immune mechanisms in multiple sclerosis".

We acknowledge the investigators of the original PDGene studies [31,32] for sharing the genetic data used for this study.

Funding for BCAC and iCOGS came from: Cancer Research UK [grant numbers C1287/A16563, C1287/A10118, C1287/A10710, C12292/A11174, C1281/A12014, C5047/A8384, C5047/A15007, C5047/A10692, C8197/A16565], the European Union's Horizon 2020 Research and Innovation Programme (grant numbers 634935 and 633784 for BRIDGES and B-CAST respectively), the European Community's Seventh Framework Programme under grant agreement n° 223175 [HEALTHF2-2009-223175] (COGS), the National Institutes of Health [CA128978] and Post-Cancer GWAS initiative [1U19 CA148537, 1U19 CA148065-01 (DRIVE) and 1U19 CA148112 - the GAME-ON initiative], the Department of Defence [W81XWH-10-1-0341], and the Canadian Institutes of Health Research CIHR) for the CIHR Team in Familial Risks of Breast Cancer [grant PSR-SIIRI-701]. All studies and funders as listed in Michailidou K et al (2013 and 2015) and in Guo Q et al (2015) are acknowledged for their contributions.

The authors acknowledge the essential role of the Cohorts for Heart and Aging Research in Genomic Epidemiology (CHARGE) Consortium in the development and support of this research. (See http://web.chargeconsortium.com for more details) The authors thank the investigators, the staff, and the participants of each contributing cohort in the CHARGE consortium publication from which these results were obtained. The citation for the CHARGE Consortium is "Cohorts for Heart and Aging Research in Genomic Epidemiology (CHARGE) Consortium: Design of prospective meta-analyses of genome-wide association studies from five cohorts. Circ Cardiovasc Genet 2:73-80, 2009. Support for the CHARGE consortium infrastructure was provided by the NIH grant R01 HL105756 (B Psaty). Support for establishing and curation of the dbGaP CHARGE Summary site (phs000930) was provided by the University of Virginia. The datasets used for the analyses in this manuscript were obtained from dbGaP through dbGaP accession study number phs000930.v9.p1.

The authors wish to acknowledge the CORECT study which was supported by the National Cancer Institute, National Institutes of Health under RFA # CA-09-002, NIH/NCI U19 CA148107 and NIH/NCI P30 CA014089. The datasets used for the analyses in this manuscript were obtained from dbGaP through dbGaP accession study number phs001499.v1.p1.

Data on coronary artery disease / myocardial infarction have been contributed by CARDIoGRAMplusC4D investigators and have been downloaded from www.CARDIOGRAMPLUSC4D.ORG.

The CIMBA data management and data analysis were supported by Cancer Research UK grants C12292/A20861, C12292/A11174. iCOGS: the European Community's Seventh Framework Programme under grant agreement n° 223175 (HEALTH-F2-2009-223175) (COGS), Cancer Research UK (C1287/A10118, C1287/A 10710, C12292/A11174, C1281/A12014, C5047/A8384, C5047/A15007, C5047/A10692, C8197/A16565), the National Institutes of Health (CA128978) and Post-Cancer GWAS initiative (1U19 CA148537, 1U19 CA148065 and 1U19 CA148112 – the GAME-ON initiative), the Department of Defence (W81XWH-10-1-0341), the Canadian Institutes of Health Research (CIHR) for the CIHR Team in Familial Risks of Breast Cancer (CRN-87521), and the Ministry of Economic Development, Innovation and Export Trade (PSR-SIIRI-701), Komen Foundation for the Cure, the Breast Cancer Research Foundation, and the Ovarian Cancer Research Fund. The PERSPECTIVE project was supported by the Government of Canada through Genome Canada and the Canadian Institutes of Health Research, the Ministry of Economy, Science and Innovation through Genome Québec, and The Quebec Breast Cancer Foundation. All studies and funders are listed in Milne et al (Nat Genet, 2017) and Phelan et al (Nat Genet, 2017).

The authors thank Million Veteran Program (MVP) staff, researchers, and volunteers, who have contributed to MVP, and especially participants who previously served their country in the military and now generously agreed to enroll in the study. (See https://www.research.va.gov/mvp/ for more details). The citation for MVP is Gaziano, J.M. et al. Million Veteran Program: A mega-biobank to study genetic influences on health and disease. J Clin Epidemiol 70, 214-23 (2016). This research is based on data from the Million Veteran Program, Office of Research and Development, Veterans Health Administration, and was supported by the Veterans Administration (VA) Cooperative Studies Program (CSP) award #G002. The datasets used for the analyses in this manuscript were obtained from dbGaP through dbGaP accession study number phs001672.v6.p1.

The breast cancer genome-wide association analyses for BCAC and CIMBA were supported by Cancer Research UK (C1287/A10118, C1287/A16563, C1287/A10710, C12292/A20861, C12292/A11174, C1281/A12014, C5047/A8384, C5047/A15007, C5047/A10692, C8197/A16565), The National Institutes of Health (CA128978, X01HG007492- the DRIVE consortium), the PERSPECTIVE project supported by the Government of Canada through Genome Canada and the Canadian Institutes of Health Research (grant GPH-129344) and the Ministère de l'Économie, Science et Innovation du Québec through Genome Québec and the PSRSIIRI-701 grant, the Quebec Breast Cancer Foundation, the European Community's Seventh Framework Programme under grant agreement n° 223175 (HEALTH-F2-2009-223175) (COGS), the European Union's Horizon 2020 Research and Innovation Programme (634935 and 633784), the Post-Cancer GWAS initiative (U19 CA148537, CA148065 and CA148112 – the GAME-ON initiative), the Department of Defence (W81XWH-10-1-0341), the Canadian Institutes of Health Research (CIHR) for the CIHR Team in Familial Risks of Breast Cancer (CRN-87521), the Komen Foundation for the

Cure, the Breast Cancer Research Foundation and the Ovarian Cancer Research Fund. All studies and funders are listed in Zhang H et al (Nat Genet, 2020).

ELLIPSE Prostate Cancer Meta-Analysis and Genotyping. Meta-Analysis: Funding for the meta-analysis provided by NIH grant U19CA148537. de novo Genotyping: We would like to acknowledge the NCRN nurses and Consultants for their work in the UKGPCS study. We thank all the patients who took part in this study. This work was supported by Cancer Research UK (grant numbers C5047/A7357, C1287/A10118, C1287/A5260, C5047/A3354, C5047/A10692, C16913/A6135 and C16913/A6835). We would also like to thank the following for funding support: Prostate Research Campaign UK (now Prostate Cancer UK), The Institute of Cancer Research and The Everyman Campaign, The National Cancer Research Network UK, The National Cancer Research Institute (NCRI) UK. We are grateful for support of NIHR funding to the NIHR Biomedical Research Centre at The Institute of Cancer Research and The Royal Marsden NHS Foundation Trust. The MEC was supported by NIH grants CA63464, CA54281 and CA098758. The datasets used for the analyses in this manuscript were obtained from dbGaP through dbGaP accession study number phs001120.v2.p2.

We want to acknowledge the participants and investigators of FinnGen study.

We thank Douglas P. Wightman, Florence Demenais, Mark M. Iles, Jia Nee and John Perry for additional assistance with external summary statistics.

Funding support for the GWAS of Venous Thrombosis study was provided through the NIH Genes, Environment and Health Initiative [GEI] (U01HG004735). The GWAS of Venous Thrombosis study is one of the genome-wide association studies funded as part of the Gene Environment Association Studies (GENEVA) under GEI. Assistance with phenotype harmonization and genotype cleaning, as well as with general study coordination, was provided by the GENEVA Coordinating Center (U01 HG004446). Assistance with data cleaning was provided by the National Center for Biotechnology Information. Funding support for genotyping, which was performed at the Johns Hopkins University Center for Inherited Disease Research, was provided by the NIH GEI (U01HG004438) and the NIH contract “High throughput genotyping for studying the genetic contributions to human disease” (HHSN268200782096C). The datasets used for the analyses in this manuscript were obtained from dbGaP through dbGaP accession study number phs000289.v2.p1.

Funding support for Genetics & Epidemiology of Colorectal Cancer Consortium (GECCO) was provided by the National Cancer Institute, National Institutes of Health, U.S. Department of Health and Human Services (X01 HG008596; U01 CA164930). Funding support for studies participating in the Genetics & Epidemiology of Colorectal Cancer Consortium and included in this study are as follows:
BWHS: National Institutes of Health (R01 CA058420, UM1 CA164974, and R01 CA098663).
CLUE-II: American Institute for Cancer Research, the Maryland Cigarette Restitution Fund at Johns Hopkins, and National Institutes of Health (P30 CA006973 to W.G. Nelson).
CORSA: Austrian Research Funding Agency (FFG) grant 829675.
CPS-II: Grants from the American Cancer Society (to P.T. Campbell and S.M. Gapstur).

Czech CCS: Ministry of Health of the Czech Republic (grants AZV 15-26535A and AZV 15-27580A), Ministry of Education and Youth of the Czech Republic (COST LD 14050) and EU COST Action BM 1206 (Eucolongene).
EDRN: NCI, EDRN Grant (U01 CA 84968-06).
EPICOLON: Fondo de Investigación Sanitaria/FEDER (14/00173, 14/00230), Ministerio de Economia y Competitividad (SAF2014-54453R), Fundación Científica de la Asociación Española contra el Cáncer (GCB13131592CAST), Agència de Gestió d'Ajuts Universitaris i de Recerca (AGAUR, Generalitat de Catalunya, 2014SGR135, 2014SGR255), Catalan Tumour Bank Network (Pla Director d'Oncologia, Generalitat de Catalunya) and COST Action BM1206. CIBERehd is funded by the Instituto de Salud Carlos III.
Hawaii Adenoma Study: National Institutes of Health (R01 CA72520).
HNPCC: National Institutes of Health (CA67941, and CA16058).
HPFS: National Institutes of Health (P01 CA055075, UM1 CA167552, R01 CA137178, R01 CA151993, R35 CA197735, K07 CA190673, and P50 CA127003).
LBS: National Institutes of Health (RFA CA-95-011), and through cooperative agreements with members of the colon family registry and principal investigators from Australian Colorectal Cancer Family Registry (UO1 CA097735), USC Familial Colorectal Neoplasia Collaborative Group (UO1 CA074799), Mayo Clinic Cooperative Family Registry for Colon Cancer Studies (UO1 CA074800), Ontario Registry for Studies of Familial Colorectal Cancer (UO1 CA074783), Seattle Colorectal Cancer Family Registry (UO1 CA074794), University of Hawaii Colorectal Cancer Family Registry (UO1 CA074806), and University of California, Irvine Informatics Center (UO1 CA078296).
NCCCS I/II: National Institutes of Health (R01 CA 066635, and P30 Dk 034987).
NHS: National Institutes of Health (R01 CA137178, P01 CA087969, UM1 CA186107, R01 CA151993, R35 CA197735, K07 CA190673, and P50 CA127003).
NSHDS (Umeå): Swedish Cancer Society; Cancer Research Foundation in Northern Sweden; Swedish Research Council; J C Kempe Memorial Fund; Faculty of Medicine, Umeå University, Umeå, Sweden; and Cutting-Edge Research Grant from the County Council of Västerbotten, Sweden.
OCCPI: National Institutes of Health (P30 CA16058).
PLCO: National Institutes of Health, to 10 PLCO screening centers, a coordinating center, a data management center and sample processing laboratories: University of Colorado Denver (N01-CN-25514); Georgetown University (N01-CN-25522); Pacific Health Research Institute (N01-CN-25515); Henry Ford Health System (N01-CN-25512); University of Minnesota (N01-CN-25513); Washington University (N01-CN-25516); University of Pittsburgh (N01-CN-25511); University of Utah (N01-CN-25524); Marshfield Clinic Research Foundation (N01-CN-25518); University of Alabama at Birmingham (N01-CN-75022); Westat CC (N01-CN-25476); IMS, Inc. (HHSN261201300008I); and Leidos, Inc. (HHSN261200800001E).
SELECT: National Institutes of Health (U10CA37429 and 5UM1CA182883).
SMS-REACH: National Institutes of Health (P01 CA074184 to J.D.P. and P.A.N.; R01 CA097325, R03 CA153323, and K05 CA152715 to P.A.N.; and KL2 TR000421 to A.N.B.-H.).
WHI: National Institutes of Health (contracts HHSN268201600018C, HHSN268201600001C, HHSN268201600002C, HHSN268201600003C, and HHSN268201600004C). Please refer to the Women's Health Initiative Clinical Trial and Observational Study page on dbGaP (phs000200).

The MEGASTROKE project received funding from sources specified at http://www.megastroke.org/acknowledgments.html. MEGASTROKE consortium members: Rainer Malik, Ganesh Chauhan, Matthew Traylor, Muralidharan Sargurupremraj, Yukinori Okada, Aniket Mishra, Loes Rutten-Jacobs, Anne-Katrin Giese, Sander W. van der Laan, Solveig Gretarsdottir, Christopher D. Anderson, Michael Chong, Hieab H. H. Adams, Tetsuro Ago, Peter Almgren, Philippe Amouyel, Hakan Ay, Traci M. Bartz, Oscar R. Benavente, Steve Bevan, Giorgio B. Boncoraglio, Robert D. Brown Jr, Adam S. Butterworth, Caty Carrera, Cara L. Carty, Daniel I. Chasman, Wei-Min Chen, John W. Cole, Adolfo Correa, Ioana Cotlarciuc, Carlos Cruchaga, John Danesh, Paul I. W. de Bakker, Anita L. DeStefano, Marcel den Hoed, Qing Duan, Stefan T. Engelter, Guido J. Falcone, Rebecca F. Gottesman, Raji P. Grewal, Vilmundur Gudnason, Stefan Gustafsson, Jeffrey Haessler, Tamara B. Harris, Ahamad Hassan, Aki S. Havulinna, Susan R. Heckbert, Elizabeth G. Holliday, George Howard, Fang-Chi Hsu, Hyacinth I. Hyacinth, M. Arfan Ikram, Erik ingelsson, Marguerite R. Irvin, Xueqiu Jian, Jordi Jimenez-Conde, Julie A. Johnson, J. Wouter Jukema, Masahiro Kanai, Keith L. Keene, Brett M. Kissela, Dawn O. Kleindorfer, Charles Kooperberg, Michiaki Kubo, Leslie A. Lange, Carl D. Langefeld, Claudia Langenberg, Lenore J. Launer, Jin-Moo Lee, Robin Lemmens, Didier Leys, Cathryn M. Lewis, Wei-Yu Lin, Arne G. Lindgren, Erik Lorentzen, Patrik K. Magnusson, Jane Maguire, Ani Manichaikul, Patrick F. McArdle, James F. Meschia, Braxton D. Mitchell, Thomas H. Mosley, Michael A. Nalls, Toshiharu Ninomiya, Martin J. O'Donnell, Bruce M. Psaty, Sara L. Pulit, Kristiina Rannikmäe, Alexander P. Reiner, Kathryn M. Rexrode, Kenneth Rice, Stephen S. Rich, Paul M. Ridker, Natalia S. Rost, Peter M. Rothwell, Jerome I. Rotter, Tatjana Rundek, Ralph L. Sacco, Saori Sakaue, Michele M. Sale, Veikko Salomaa, Bishwa R. Sapkota, Reinhold Schmidt, Carsten O. Schmidt, Ulf Schminke, Pankaj Sharma, Agnieszka Slowik, Cathie L. M. Sudlow, Christian Tanislav, Turgut Tatlisumak, Kent D. Taylor, Vincent N. S. Thijs, Gudmar Thorleifsson, Unnur Thorsteinsdottir, Steffen Tiedt, Stella Trompet, Christophe Tzourio, Cornelia M. van Duijn, Matthew Walters, Nicholas J. Wareham, Sylvia Wassertheil-Smoller, James G. Wilson, Kerri L. Wiggins, Qiong Yang, Salim Yusuf, Najaf Amin, Hugo S. Aparicio, Donna K. Arnett, John Attia, Alexa S. Beiser, Claudine Berr, Julie E. Buring, Mariana Bustamante, Valeria Caso, Yu-Ching Cheng, Seung Hoan Choi, Ayesha Chowhan, Natalia Cullell, Jean-Franęois Dartigues, Hossein Delavaran, Pilar Delgado, Marcus Dörr, Gunnar Engström, Ian Ford, Wander S. Gurpreet, Anders Hamsten, Laura Heitsch, Atsushi Hozawa, Laura Ibanez, Andreea Ilinca, Martin Ingelsson, Motoki Iwasaki, Rebecca D. Jackson, Katarina Jood, Pekka Jousilahti, Sara Kaffashian, Lalit Kalra, Masahiro Kamouchi, Takanari Kitazono, Olafur Kjartansson, Manja Kloss, Peter J. Koudstaal, Jerzy Krupinski, Daniel L. Labovitz, Cathy C. Laurie, Christopher R. Levi, Linxin Li, Lars Lind, Cecilia M. Lindgren, Vasileios Lioutas, Yong Mei Liu, Oscar L. Lopez, Hirata Makoto, Nicolas Martinez-Majander, Koichi Matsuda, Naoko Minegishi, Joan Montaner, Andrew P. Morris, Elena Muiño, Martina Müller-Nurasyid, Bo Norrving, Soichi Ogishima, Eugenio A. Parati, Leema Reddy Peddareddygari, Nancy L. Pedersen, Joanna Pera, Markus Perola, Alessandro Pezzini, Silvana Pileggi, Raquel Rabionet, Iolanda Riba-Llena, Marta Ribases, Jose R. Romero, Jaume Roquer, Anthony G. Rudd, Antti-Pekka Sarin, Ralhan Sarju, Chloe Sarnowski, Makoto Sasaki, Claudia L. Satizabal, Mamoru Satoh, Naveed Sattar, Norie Sawada, Gerli Sibolt, Asgeir Sigurdsson, Albert Smith, Kenji Sobue, Carolina Soriano-Tarraga, Tara Stanne, O. Colin Stine, David J. Stott, Konstantin Strauch, Takako Takai, Hideo Tanaka,

Kozo Tanno, Alexander Teumer, Liisa Tomppo, Nuria P. Torres-Aguila, Emmanuel Touze, Shoichiro Tsugane, Andre G. Uitterlinden, Einar M. Valdimarsson, Sven J. van der Lee, Henry Völzke, Kenji Wakai, David Weir, Stephen R. Williams, Charles D. A. Wolfe, Quenna Wong, Huichun Xu, Taiki Yamaji, Dharambir K. Sanghera, Olle Melander, Christina Jern, Daniel Strbian, Israel Fernandez-Cadenas, W. T. Longstreth Jr, Arndt Rolfs, Jun Hata, Daniel Woo, Jonathan Rosand, Guillaume Pare, Jemma C. Hopewell, Danish Saleheen, Kari Stefansson, Bradford B. Worrall, Steven J. Kittner, Sudha Seshadri, Myriam Fornage, Hugh S. Markus, Joanna M. M. Howson, Yoichiro Kamatani, Stephanie Debette and Martin Dichgans.

### Supplementary figures

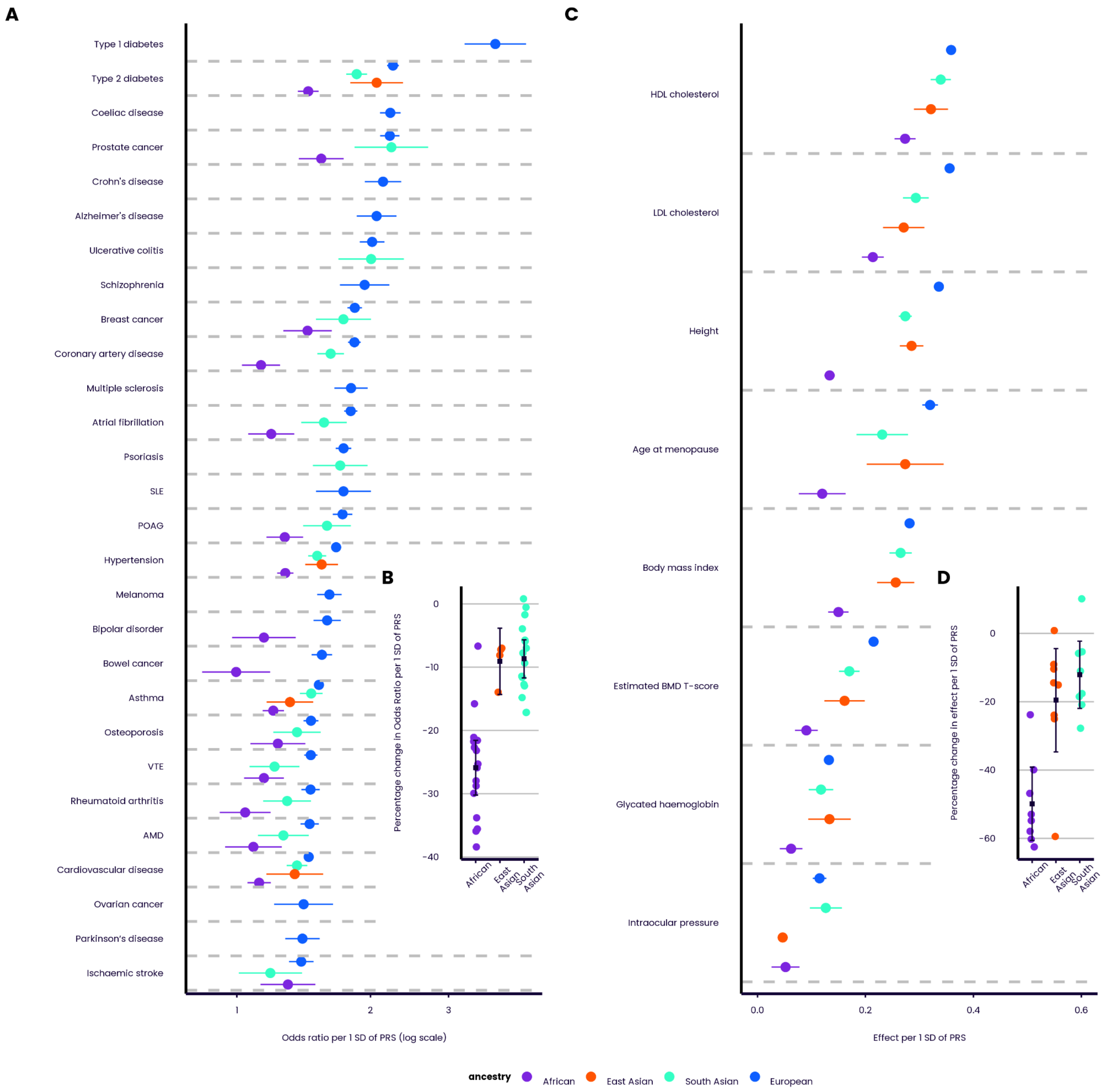


**Supplementary Figure 1. Predictive performance of the UK Biobank PRS Release (Standard Set) by ancestry** . Performance (odds ratio, or effect on standardised quantitative trait, per SD of PRS, adjusting for age and sex), measured in the independent UKB Testing Subgroup, of the disease traits (**A**) and quantitative traits (**C**), stratified by genetically inferred ancestry. Results for non-European ancestries are shown if at least 100 cases are available for testing. Relative change in performance in non-European ancestries compared to European ancestry for disease traits (**B**) and quantitative traits (**D**). Bars indicate 95% confidence intervals (CI). Refer to Figure 1 and 2 for disease and quantitative trait abbreviations respectively. Throughout, ovarian cancer refers specifically to epithelial ovarian cancer.

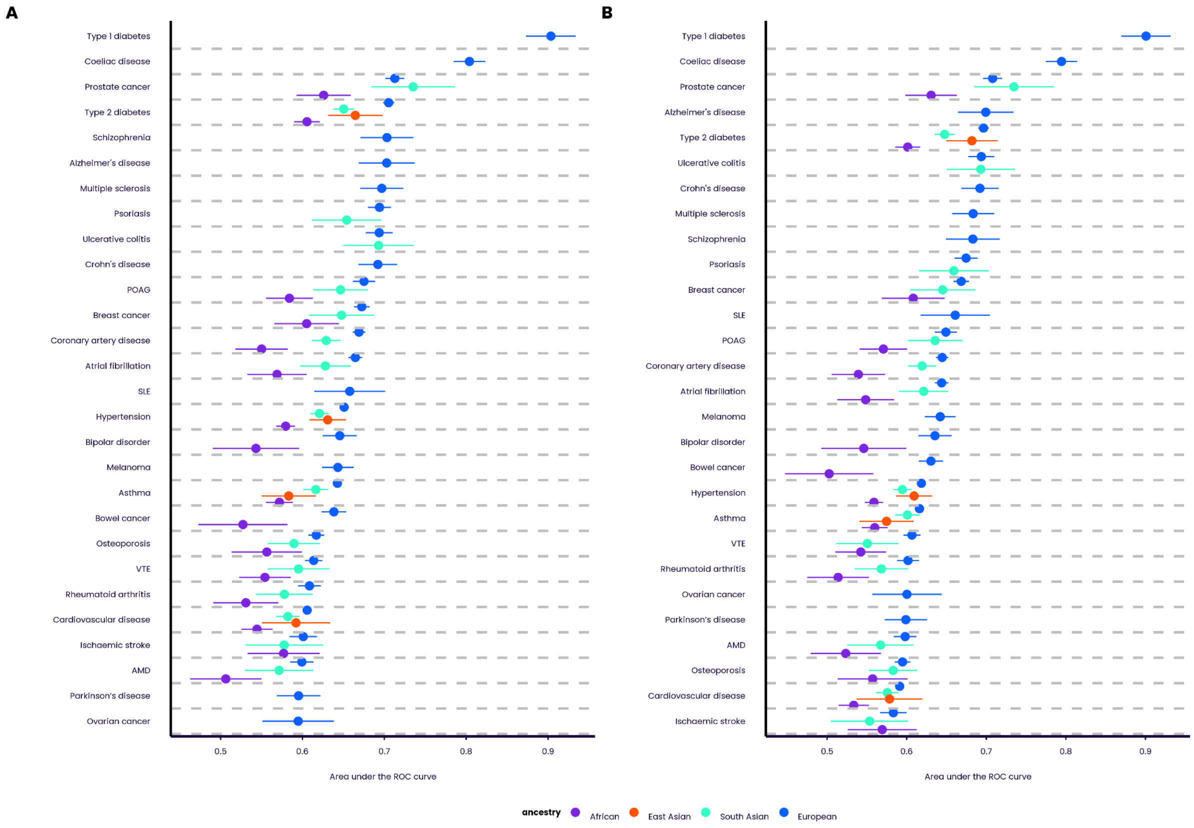


**Supplementary Figure 2. Predictive performance (AUC) of the UK Biobank PRS Release disease traits by ancestry**. Performance (area under the receiver operating characteristic curve, AUC), measured in the independent UKB Testing Subgroup, of the disease traits in the Standard (**A**) and Enhanced (**B**) PRS sets, stratified by genetically inferred ancestry. Results for non-European ancestries are shown if at least 100 cases are available for testing. Bars indicate 95% confidence intervals (CI). Refer to Figure 1 for disease abbreviations.

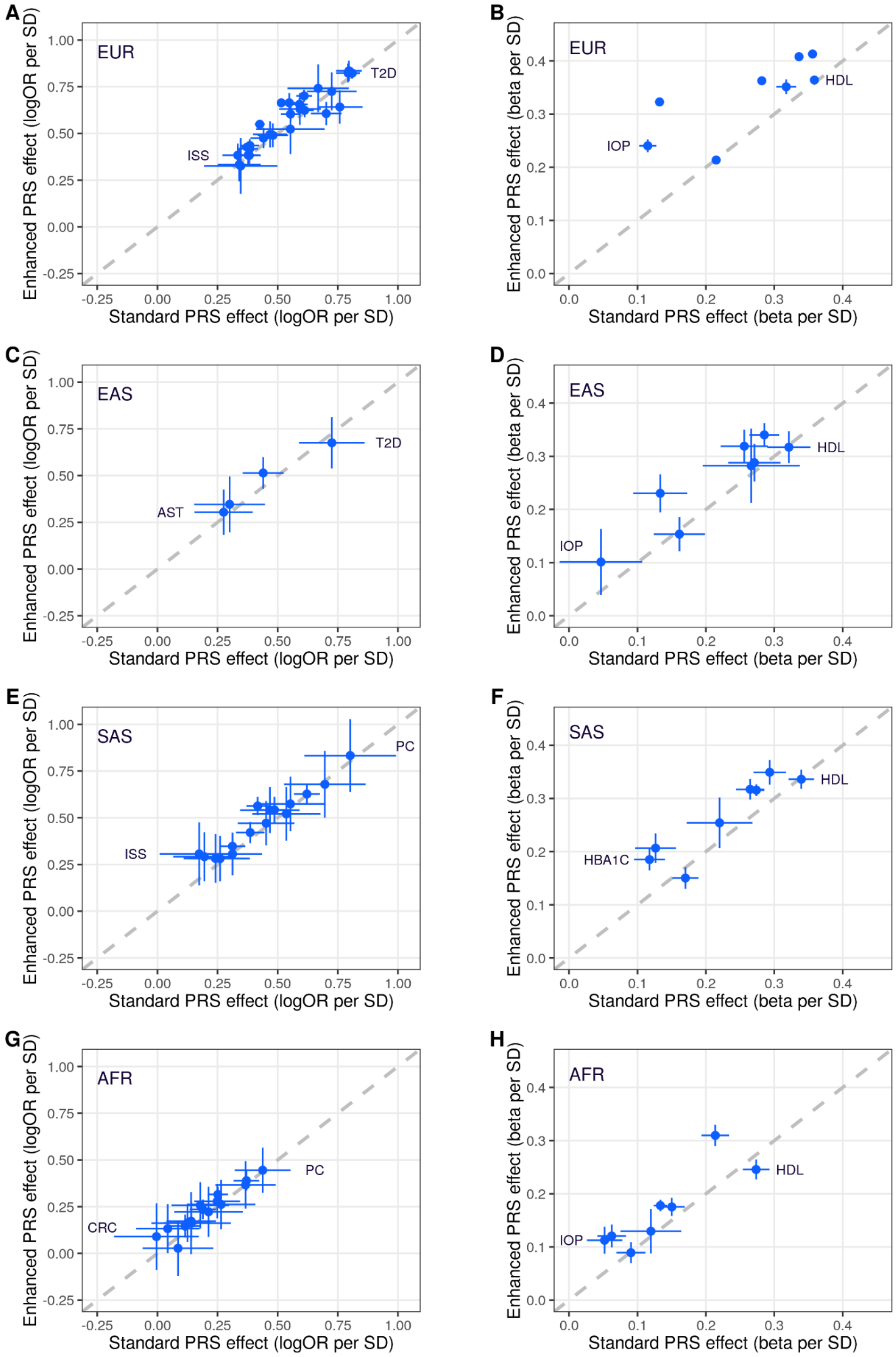


**Supplementary Figure 3. Comparison of the predictive performance of the Standard and Enhanced PRS sets**. Performance (odds ratio, or effect on standardised quantitative trait, per SD of PRS, adjusting for age and sex), measured in the independent UKB Testing Subgroup, of the disease traits (**A, C, E, G**) and quantitative traits (**B, D, F, H**) in the Standard and Enhanced PRS sets in different ancestries. EUR = European ancestry (**A, B**). EAS = East Asian ancestry (**C, D**). SAS = South Asian ancestry (**E, F**). AFR = Sub-Saharan African ancestry (**G, H**). Bars indicate 95% confidence intervals (CI). Traits with highest and lowest performance are labelled. For trait codes see Supplementary Table 5.

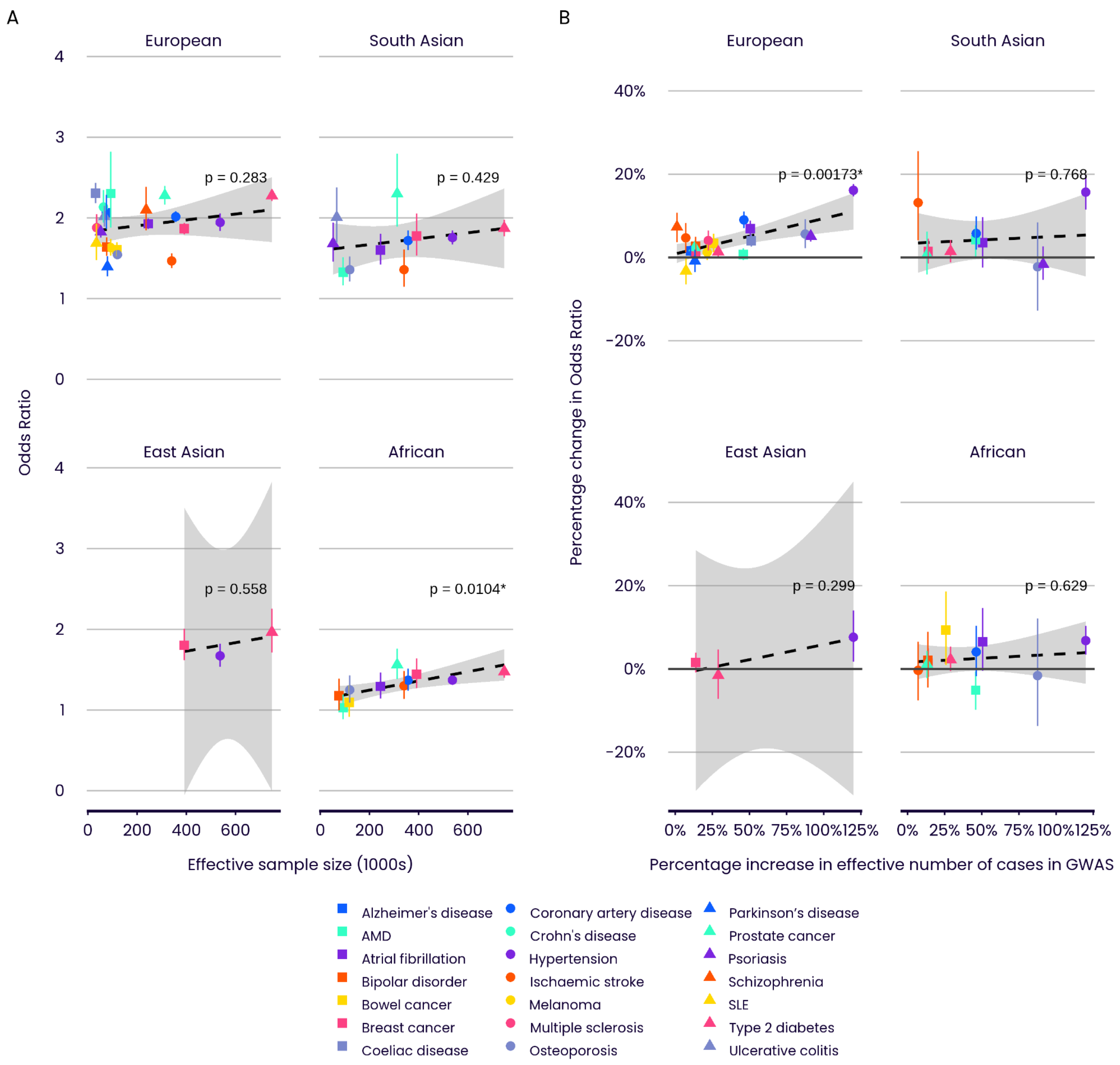


**Supplementary Figure 4. Relationship between disease trait predictive performance and GWAS effective sample size across genetically inferred ancestry groups**. **A** Relationship between odds ratio and effective sample size for the Enhanced PRS Set. **B** Relationship between relative change in odds ratio and relative change in effective sample size, comparing the Enhanced to the Standard PRS Set. Effective sample size is defined as $4\sum_j n_j c_j(1 - c_j)$ , where $n_j$ and $c_j$ are respectively the total sample size and the proportion of cases for the *j*th constituent GWAS for a given trait. Only those diseases with non-overlapping samples in the constituent GWASs are displayed. Bars indicate 95% confidence intervals. Dashed lines indicate linear regression slopes, with p-values indicating the significance of the slope. Refer to Figure 1 for disease abbreviations.

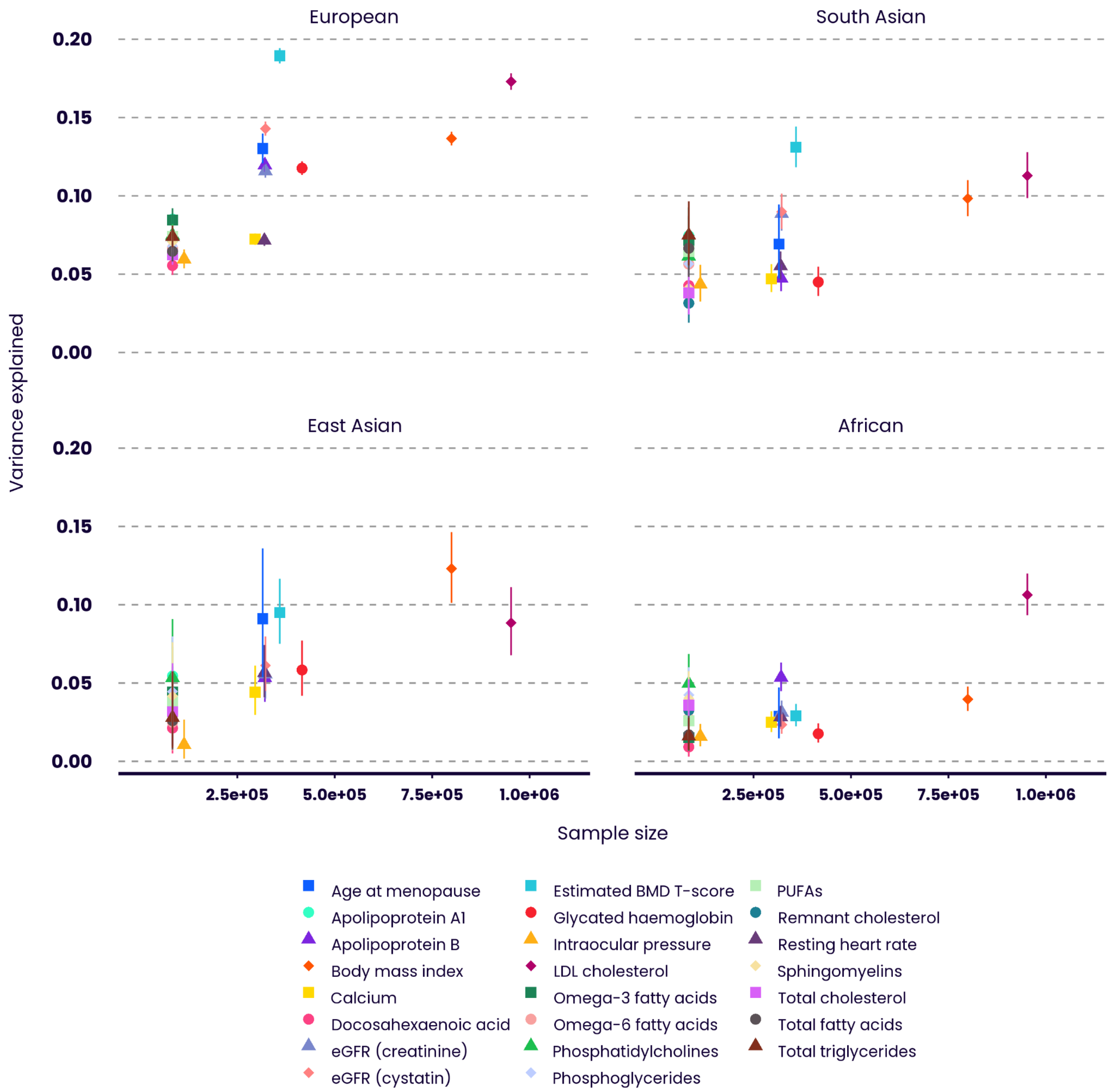


**Supplementary Figure 5**. **Relationship between quantitative trait predictive performance and GWAS sample size across genetically inferred ancestry groups**. Relationship between variance explained and sample size for the Enhanced PRS Set. Only those diseases with non-overlapping samples in the constituent GWASs are displayed. Bars indicate 95% confidence intervals. Refer to Figure 2 for quantitative trait abbreviations.

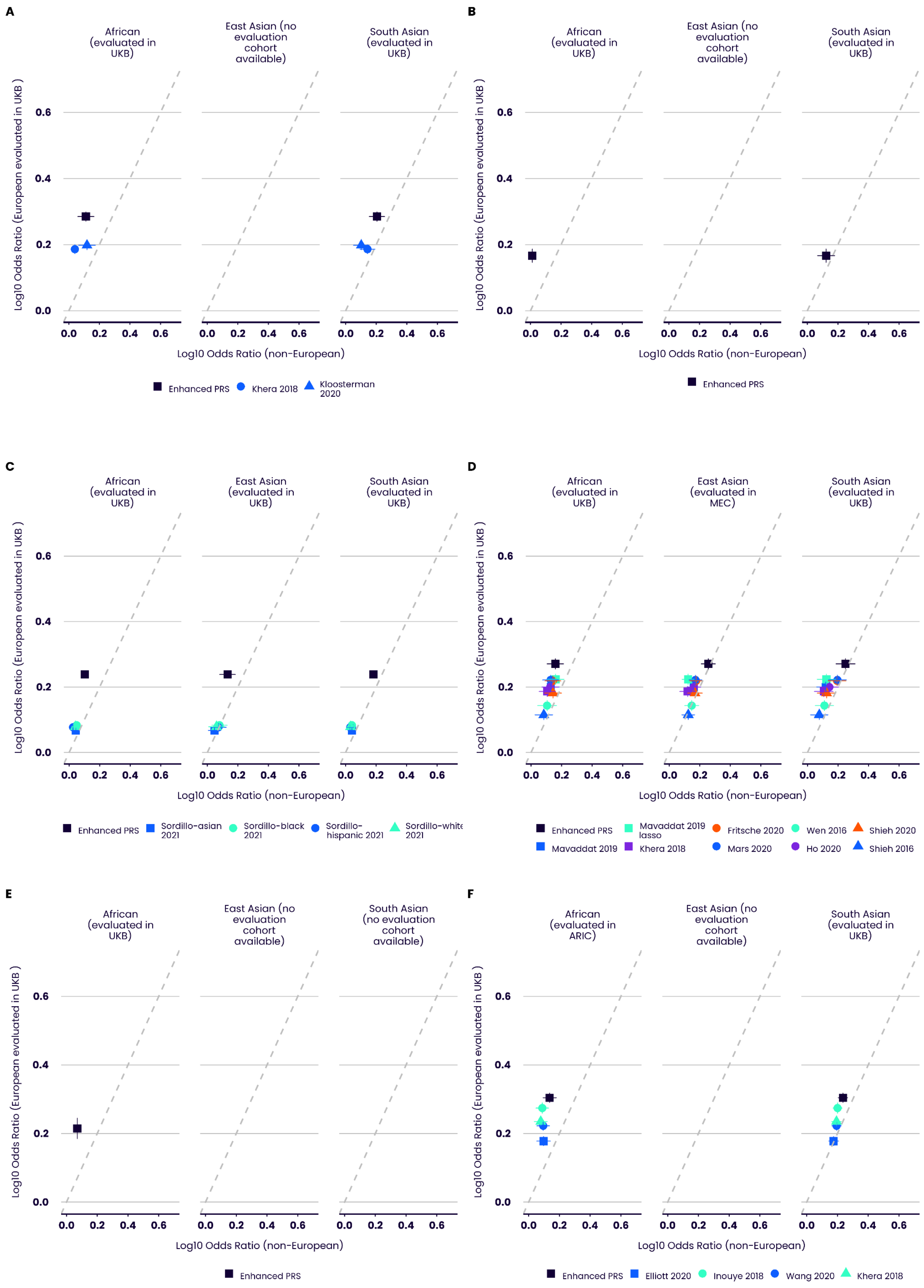
A
African (evaluated in UKB)
East Asian (no evaluation cohort available)
South Asian (evaluated in UKB)
Log10 Odds Ratio (European evaluated in UKB )
Log10 Odds Ratio (non-European)
Enhanced PRS
Khera 2018
Kloosterman 2020
B
African (evaluated in UKB)
East Asian (no evaluation cohort available)
South Asian (evaluated in UKB)
Enhanced PRS
C
African (evaluated in UKB)
East Asian (evaluated in UKB)
South Asian (evaluated in UKB)
Enhanced PRS
Sordillo-asian 2021
Sordillo-black 2021
Sordillo-hispanic 2021
Sordillo-white 2021
D
African (evaluated in UKB)
East Asian (evaluated in MEC)
South Asian (evaluated in UKB)
Enhanced PRS
Mavaddat 2019 lasso
Fritsche 2020
Wen 2016
Shieh 2020
Mavaddat 2019
Khera 2018
Mars 2020
Ho 2020
Shieh 2016
E
African (evaluated in UKB)
East Asian (no evaluation cohort available)
South Asian (no evaluation cohort available)
Enhanced PRS
F
African (evaluated in ARIC)
East Asian (no evaluation cohort available)
South Asian (evaluated in UKB)
Enhanced PRS
Elliott 2020
Inouye 2018
Wang 2020
Khera 2018

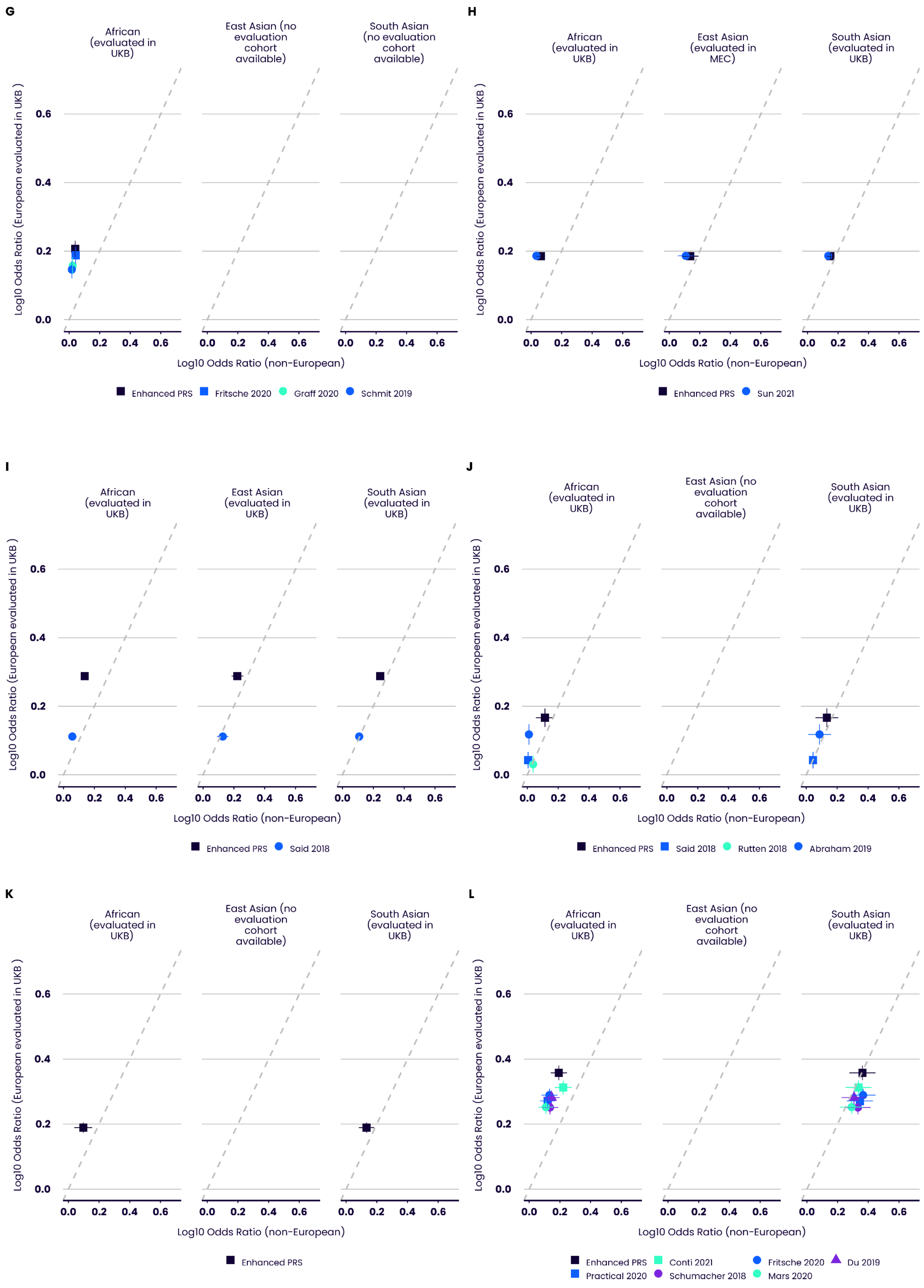
G
African (evaluated in UKB)
East Asian (no evaluation cohort available)
South Asian (no evaluation cohort available)
Log10 Odds Ratio (European evaluated in UKB )
Log10 Odds Ratio (non-European)
Enhanced PRS
Fritsche 2020
Graff 2020
Schmit 2019
H
African (evaluated in UKB)
East Asian (evaluated in MEC)
South Asian (evaluated in UKB)
Enhanced PRS
Sun 2021
I
African (evaluated in UKB)
East Asian (evaluated in UKB)
South Asian (evaluated in UKB)
Enhanced PRS
Said 2018
J
African (evaluated in UKB)
East Asian (no evaluation cohort available)
South Asian (evaluated in UKB)
Enhanced PRS
Said 2018
Rutten 2018
Abraham 2019
K
African (evaluated in UKB)
East Asian (no evaluation cohort available)
South Asian (evaluated in UKB)
Enhanced PRS
L
African (evaluated in UKB)
East Asian (no evaluation cohort available)
South Asian (evaluated in UKB)
Enhanced PRS
Conti 2021
Fritsche 2020
Du 2019
Practical 2020
Schumacher 2018
Mars 2020

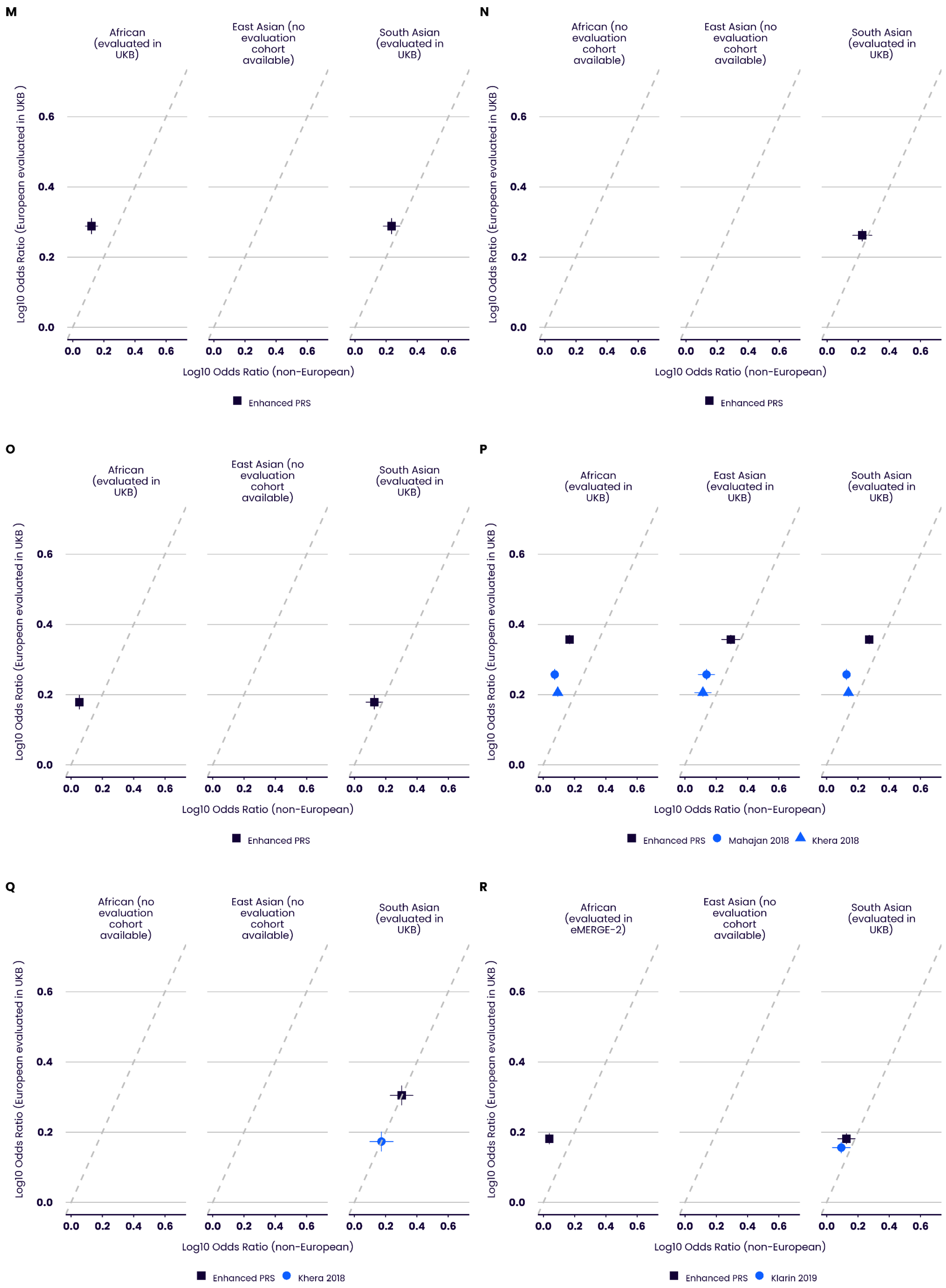


**Supplementary Figure 6. Performance relative to comparator PRSs across ancestries, disease traits.** Performance (odds ratio per SD of PRS) of the disease trait Enhanced PRS Set, compared to other published polygenic scores (citations provided in Supplementary Table 10) and comparing performance in European ancestry (y-axis) to other ancestries (x-axis). Non-UK Biobank cohorts are used if they provide a larger sample size for a given ancestry and trait than UK Biobank. **A** Atrial fibrillation. **B** Age-related macular degeneration. **C** Asthma. **D** Breast cancer. **E** Bipolar disorder. **F** Coronary artery disease. **G** Bowel cancer. **H** Cardiovascular disease. **I** Hypertension. **J** Ischaemic stroke. **K** Osteoporosis. **L** Prostate cancer. **M** Primary open angle glaucoma.**N** Psoriasis. **O** Rheumatoid arthritis. **P** Type 2 diabetes **Q** Ulcerative colitis. **R** Venous thromboembolic disease. Bars indicate 95% confidence intervals.

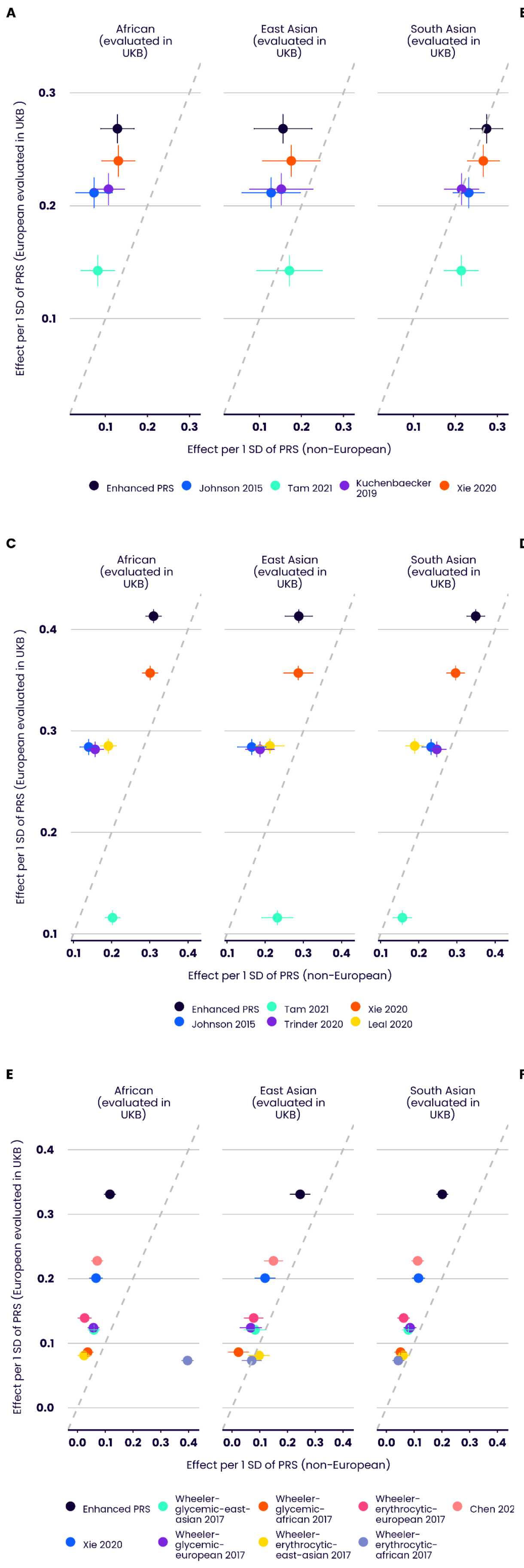
A
B
C
D
E
F
African (evaluated in UKB)
East Asian (evaluated in UKB)
South Asian (evaluated in UKB)
Effect per 1 SD of PRS (European evaluated in UKB )
Effect per 1 SD of PRS (non-European)
Enhanced PRS
Johnson 2015
Tam 2021
Kuchenbaecker 2019
Xie 2020
Trinder 2020
Leal 2020
Wheeler-glycemic-east-asian 2017
Wheeler-glycemic-african 2017
Wheeler-erythrocytic-european 2017
Chen 202
Wheeler-glycemic-european 2017
Wheeler-erythrocytic-east-asian 2017
Wheeler-erythrocytic-african 2017

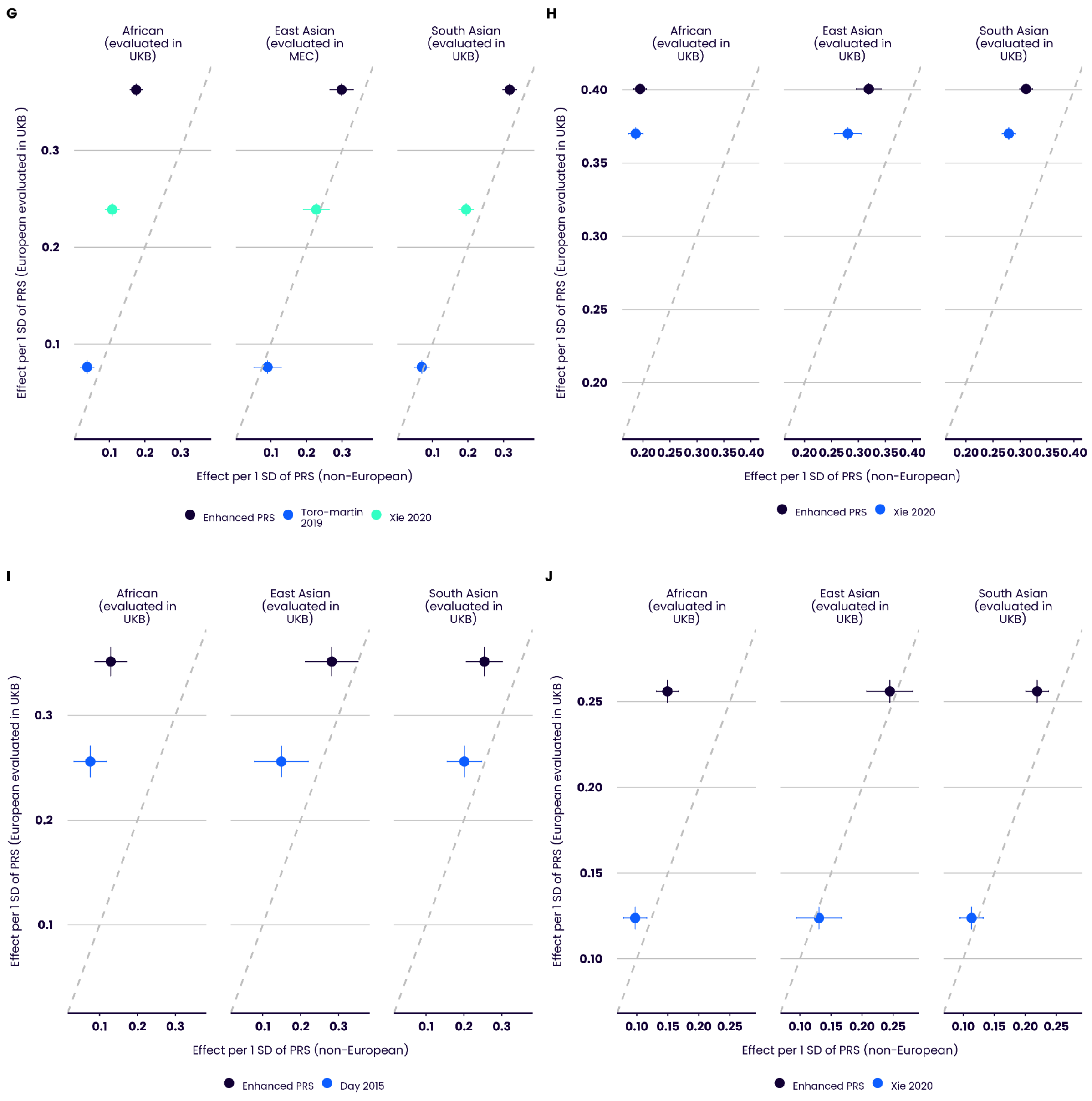


**Supplementary Figure 7. Performance relative to comparator PRS across ancestries, quantitative traits.** Performance (effect on standardised quantitative trait per SD of PRS) of the quantitative trait Enhanced PRS Set, compared to other published polygenic scores (citations provided in Supplementary Table 10) and comparing performance in European ancestry (y-axis) to other ancestries (x-axis). Non-UK Biobank cohorts are used if they provide a larger sample size for a given ancestry and trait than UK Biobank. **A** Total triglycerides. **B** Total cholesterol. **C** LDL cholesterol. **D** HDL cholesterol. **E** Glycated haemoglobin. **F** eGFR (creatinine). **G** Body mass index. **H** Height. **I** Age at menopause. **J** Resting heart rate. Bars indicate 95% confidence intervals.

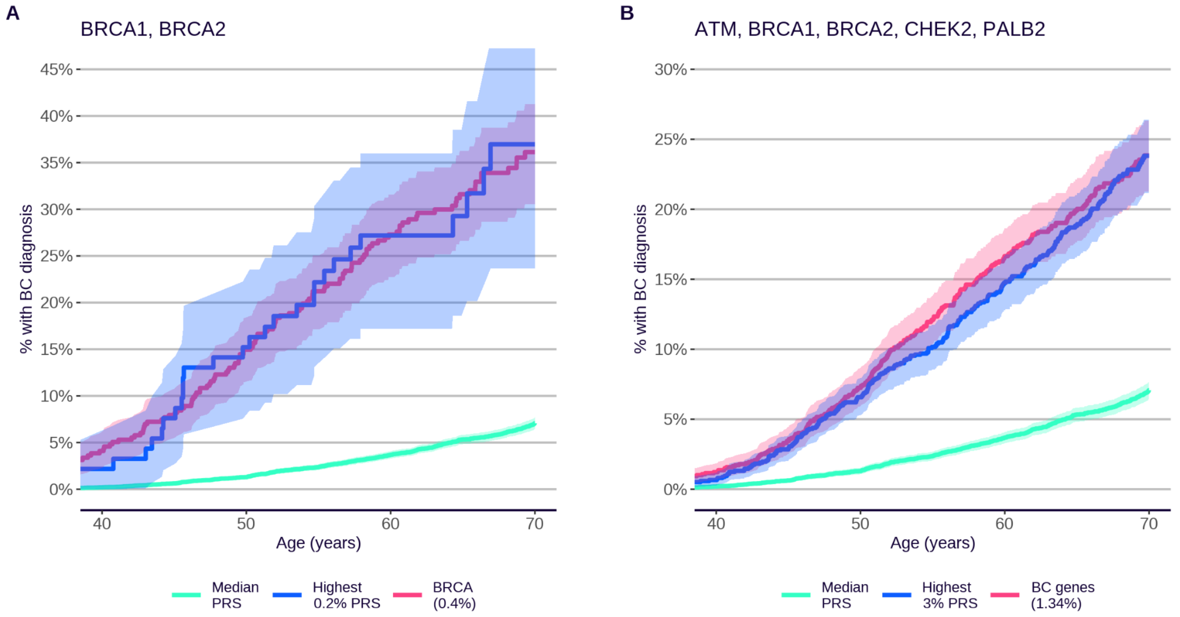


**Supplementary Figure 8. Comparative cumulative incidence plots in high-risk breast cancer (BC) gene mutation carriers and high-PRS individuals of equivalent risk**. High-risk mutation carriers (red) compared to individuals in the top percentile of the PRS distribution (blue) corresponding to the equivalent high-risk variant risk, evaluated in the subset of UKB (EUR) for whom exome sequencing data are available (for mutation carriers) or in the subset of UKB Testing Subgroup (EUR) (median and high PRS individuals). **A**, Incidence of female breast cancer in *BRCA1* + *BRCA2* loss-of-function variant carriers (415 carriers) vs breast cancer Enhanced PRS. **B**, Incidence of breast cancer in combined *BRCA1*+*BRCA2*+*ATM*+*CHEK2*+*PALB2* loss-of-function variant carriers (1,395 carriers) vs breast cancer Enhanced PRS. Shaded areas indicate 95% CI.

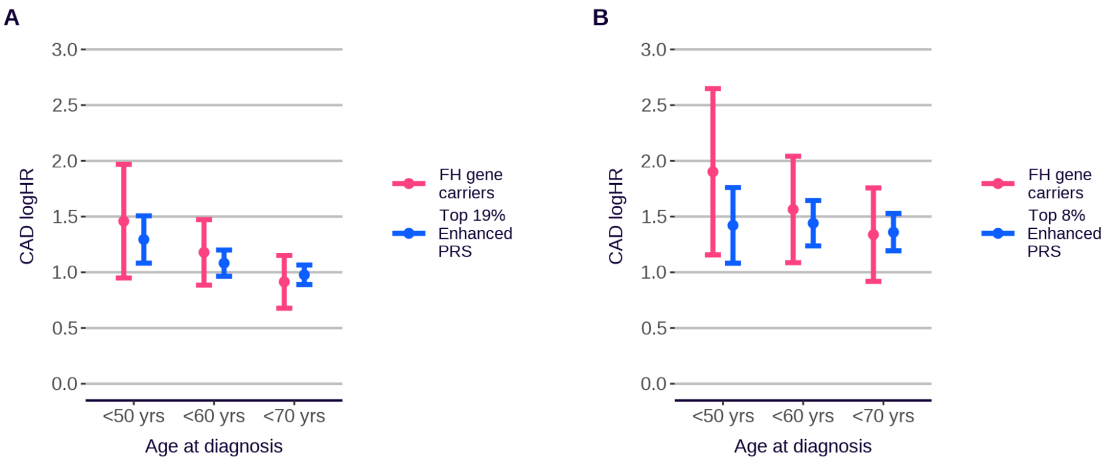


**Supplementary Figure 9. Effects of familial hypercholesterolemia (FH) gene mutations and PRS on risk of CAD, by age of diagnosis**. The log(HR) for CAD associated with pathogenic or likely pathogenic mutations in any of four FH genes (*ABOB*, *ABOE*, *LDLR*, *PCSK9*), relative to non-carriers (pink, evaluated in the subset of UKB (EUR) for whom exome sequencing data are available), or associated with a PRS (Enhanced CAD PRS) in the top 19% (**A**) or 8% (**B**) of the distribution, relative to the median PRS (40-60th percentiles) (blue). **A** shows analyses including all eligible European ancestry UKB participants in the Testing subgroup; **B** is further restricted to participants with Primary Care data linkage and with no reported statin prescription, other than prescriptions which began after a first CAD diagnosis. FH carrier effect estimated in the subset of UKB for which whole exome sequencing data are available (n=189,954 in **A** and n=75,351 in **B**). Bars represent 95% CI.

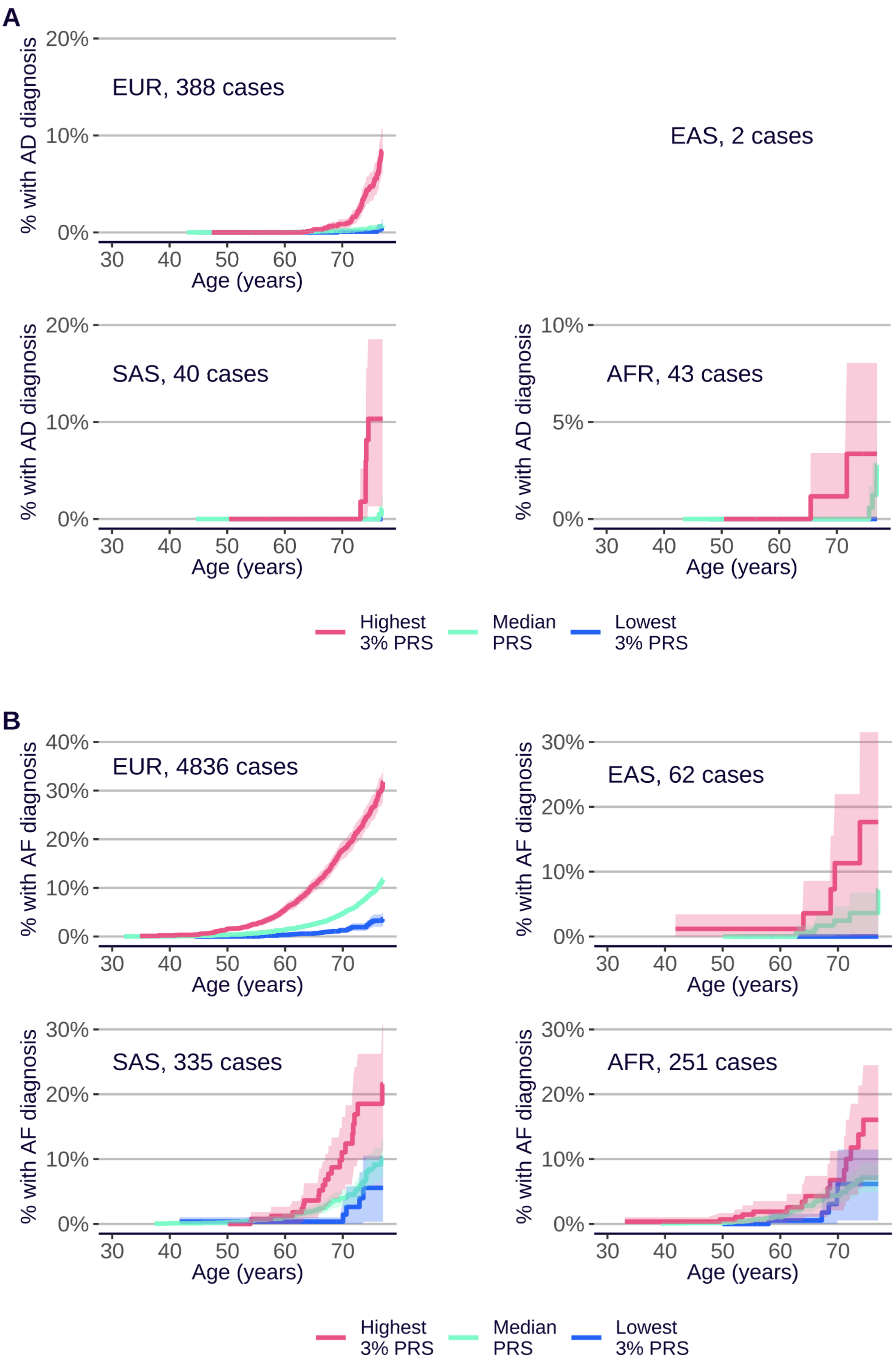
A
EUR, 388 cases
EAS, 2 cases
SAS, 40 cases
AFR, 43 cases
% with AD diagnosis
Age (years)
20%
10%
5%
0%
30
40
50
60
70
Highest 3% PRS
Median PRS
Lowest 3% PRS
B
EUR, 4836 cases
EAS, 62 cases
SAS, 335 cases
AFR, 251 cases
% with AF diagnosis
40%
30%
20%
10%
0%

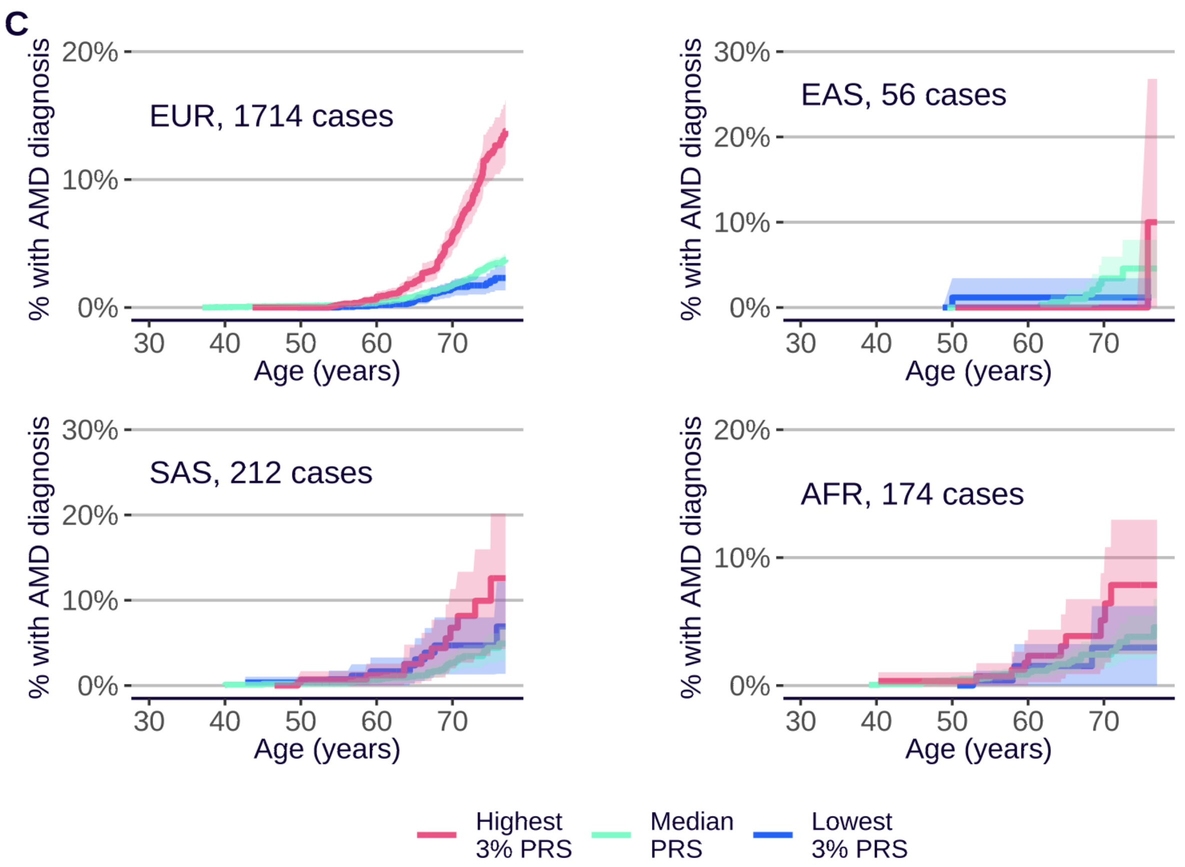
C
% with AMD diagnosis
EUR, 1714 cases
EAS, 56 cases
SAS, 212 cases
AFR, 174 cases
0%
10%
20%
30%
30 40 50 60 70
Age (years)
Highest 3% PRS
Median PRS
Lowest 3% PRS

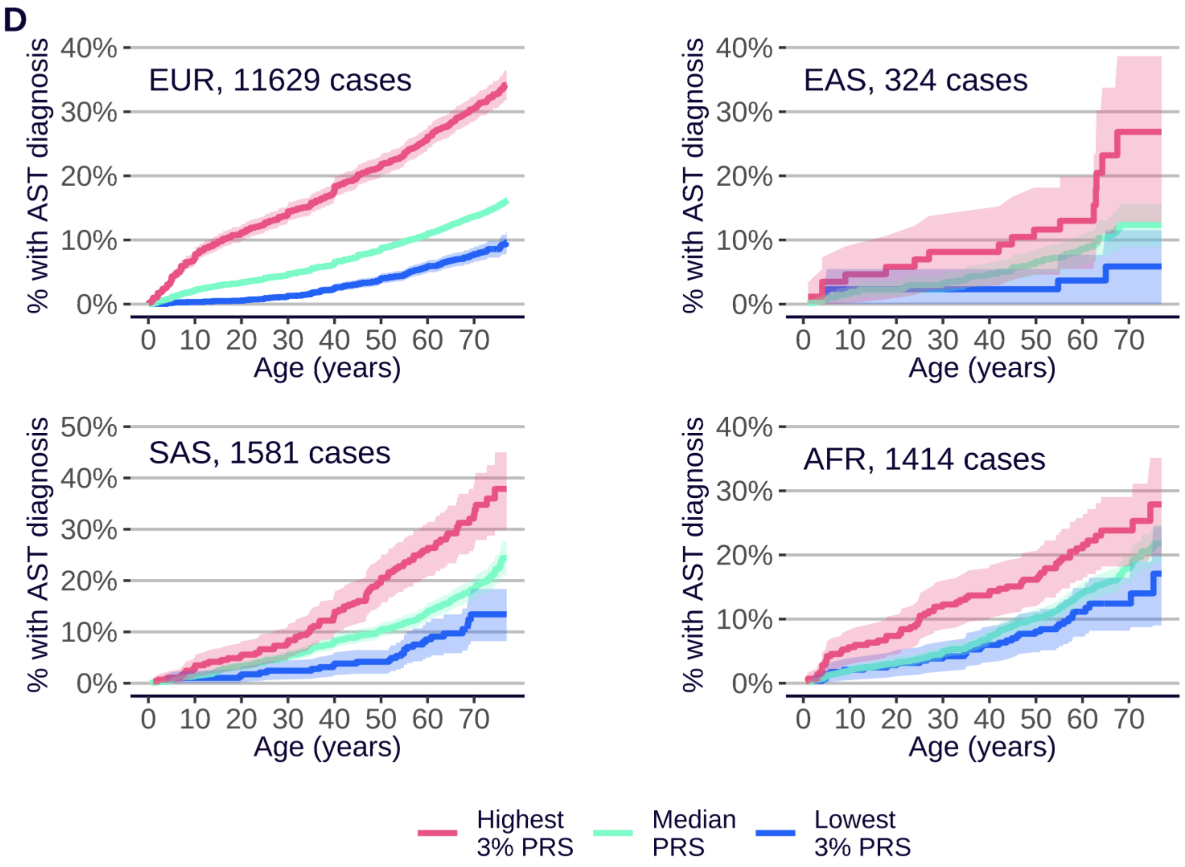
D
% with AST diagnosis
EUR, 11629 cases
EAS, 324 cases
SAS, 1581 cases
AFR, 1414 cases
0%
10%
20%
30%
40%
50%
0 10 20 30 40 50 60 70
Age (years)
Highest 3% PRS
Median PRS
Lowest 3% PRS

**E**

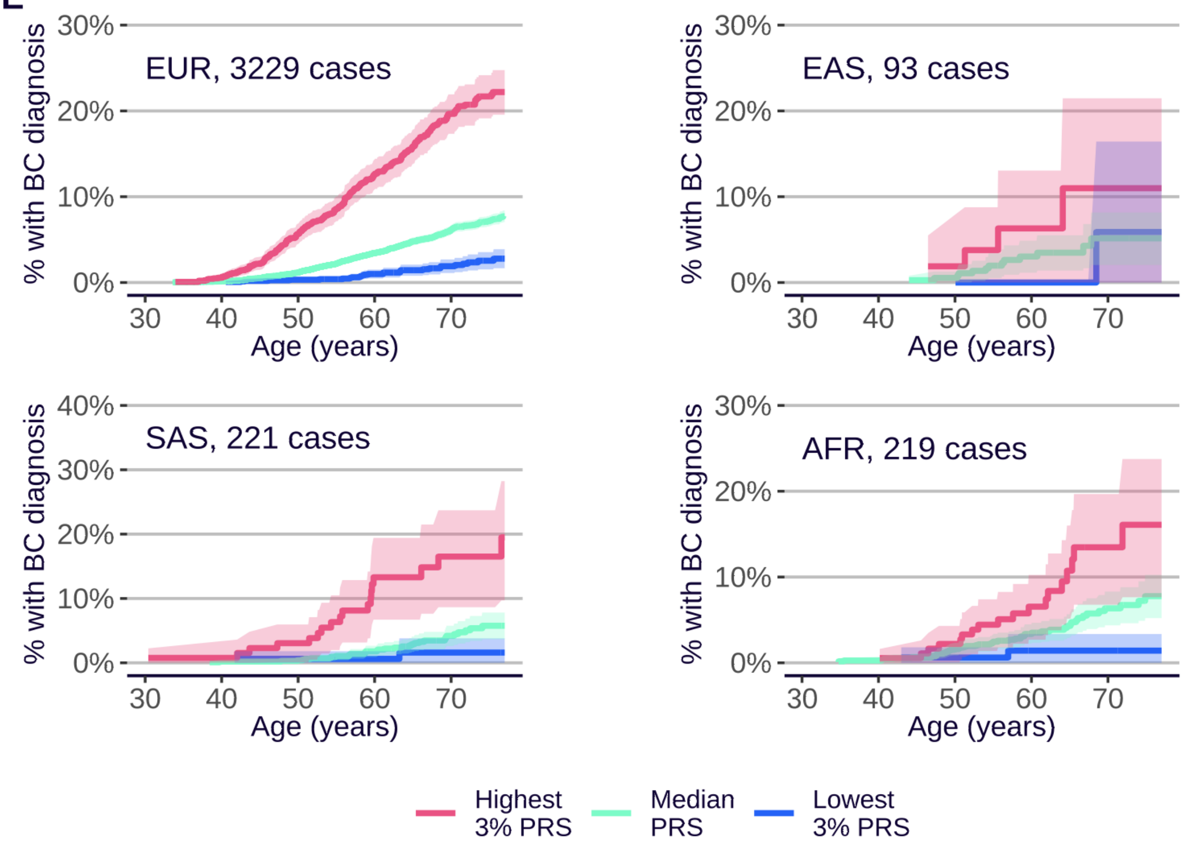
EUR, 3229 cases
EAS, 93 cases
SAS, 221 cases
AFR, 219 cases
% with BC diagnosis
Age (years)
0%
10%
20%
30%
40%
30
40
50
60
70
Highest 3% PRS
Median PRS
Lowest 3% PRS

**F**

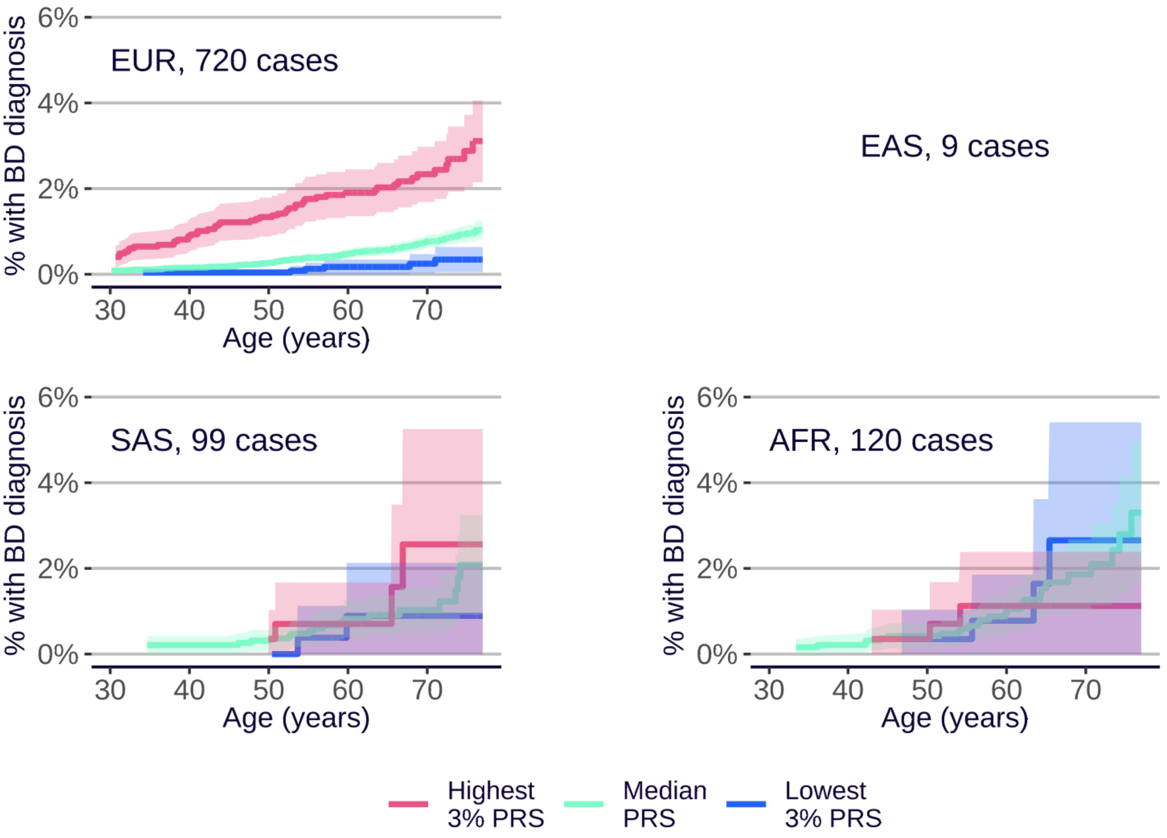
EUR, 720 cases
EAS, 9 cases
SAS, 99 cases
AFR, 120 cases
% with BD diagnosis
Age (years)
0%
2%
4%
6%
30
40
50
60
70
Highest 3% PRS
Median PRS
Lowest 3% PRS

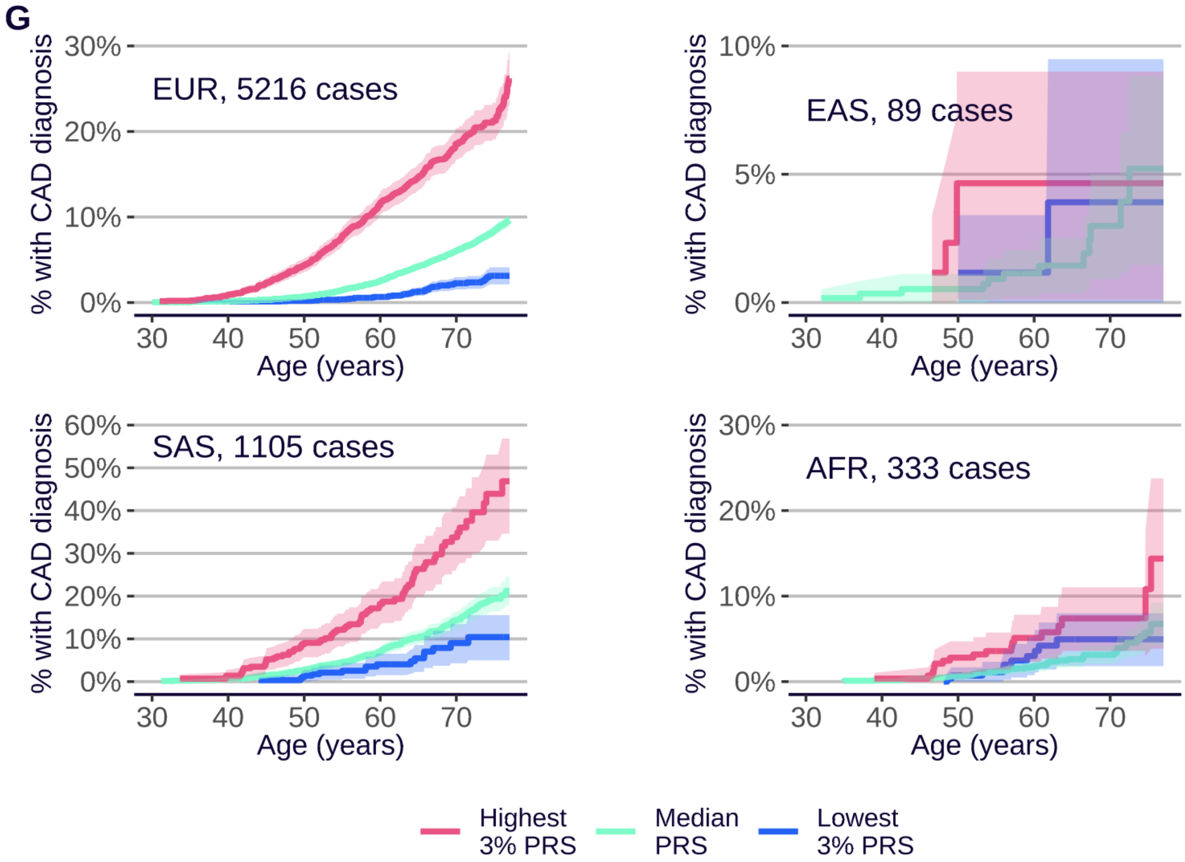
G
EUR, 5216 cases
EAS, 89 cases
SAS, 1105 cases
AFR, 333 cases
% with CAD diagnosis
Age (years)
Highest 3% PRS
Median PRS
Lowest 3% PRS

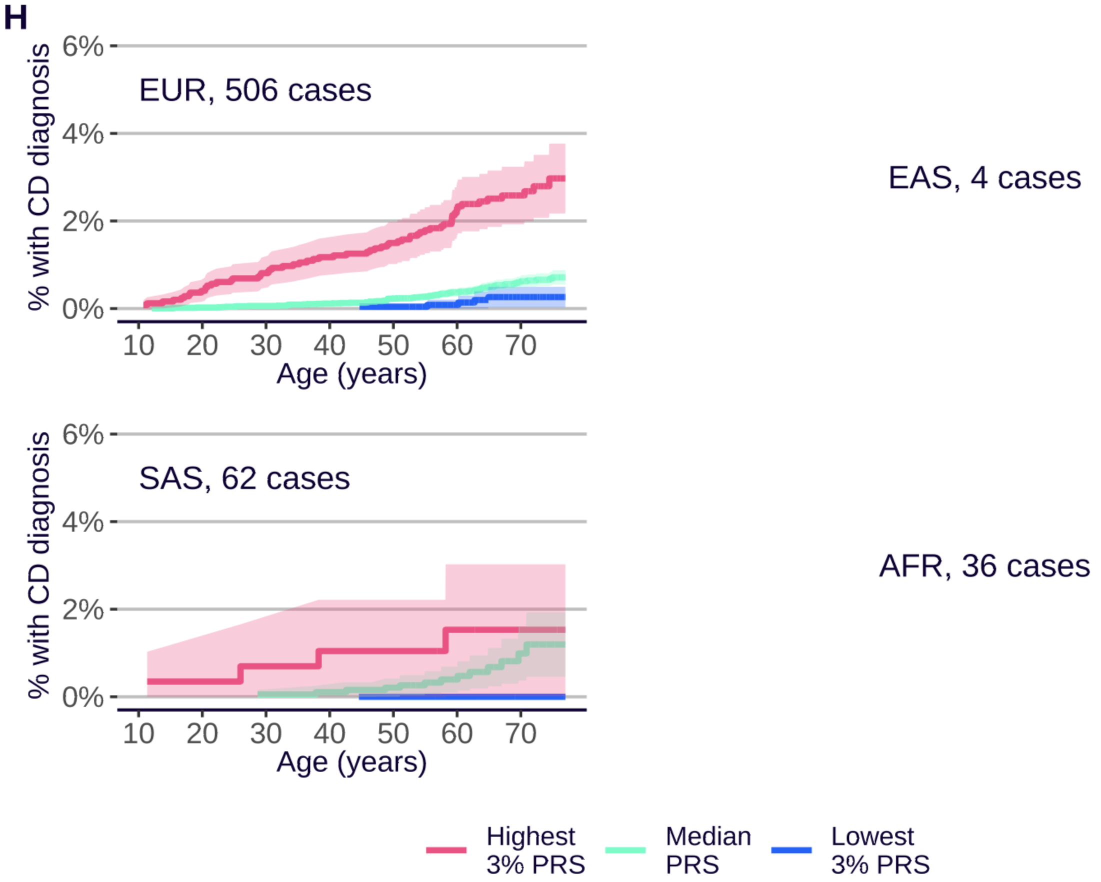
H
EUR, 506 cases
EAS, 4 cases
SAS, 62 cases
AFR, 36 cases
% with CD diagnosis
Age (years)
Highest 3% PRS
Median PRS
Lowest 3% PRS

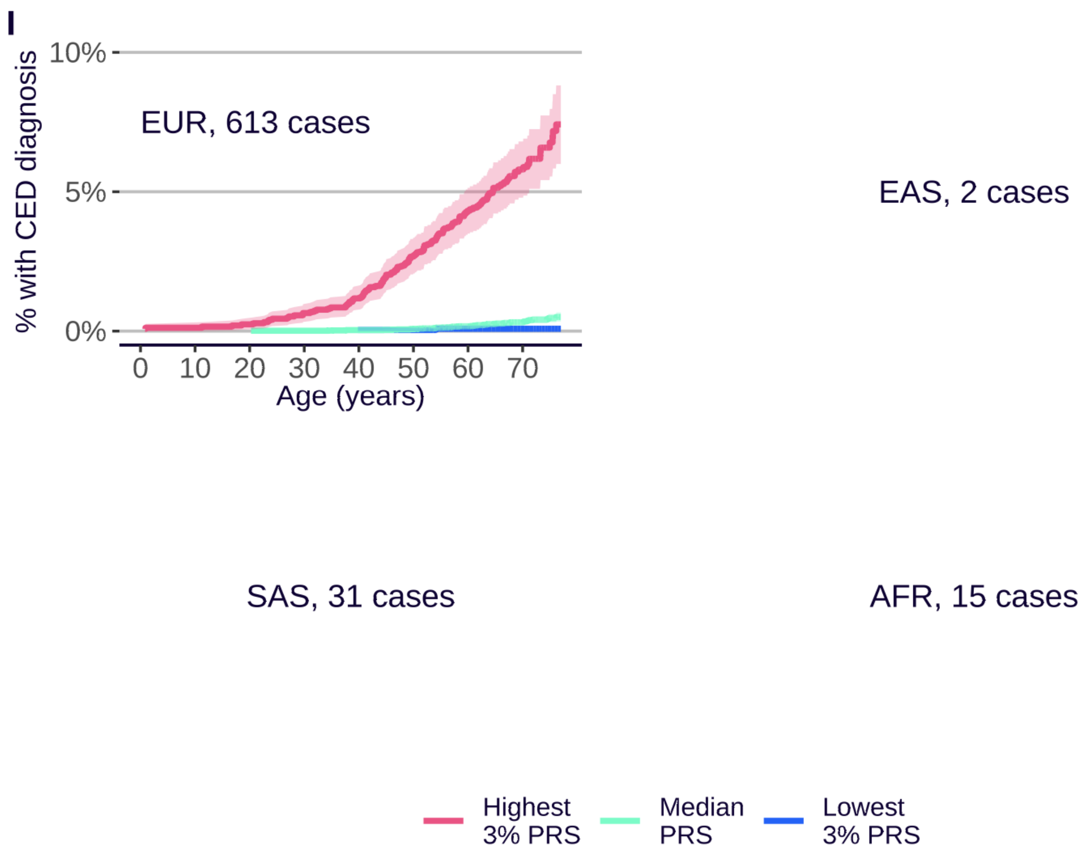
I
% with CED diagnosis
10%
5%
0%
EUR, 613 cases
0 10 20 30 40 50 60 70
Age (years)
EAS, 2 cases
SAS, 31 cases
AFR, 15 cases
Highest 3% PRS
Median PRS
Lowest 3% PRS

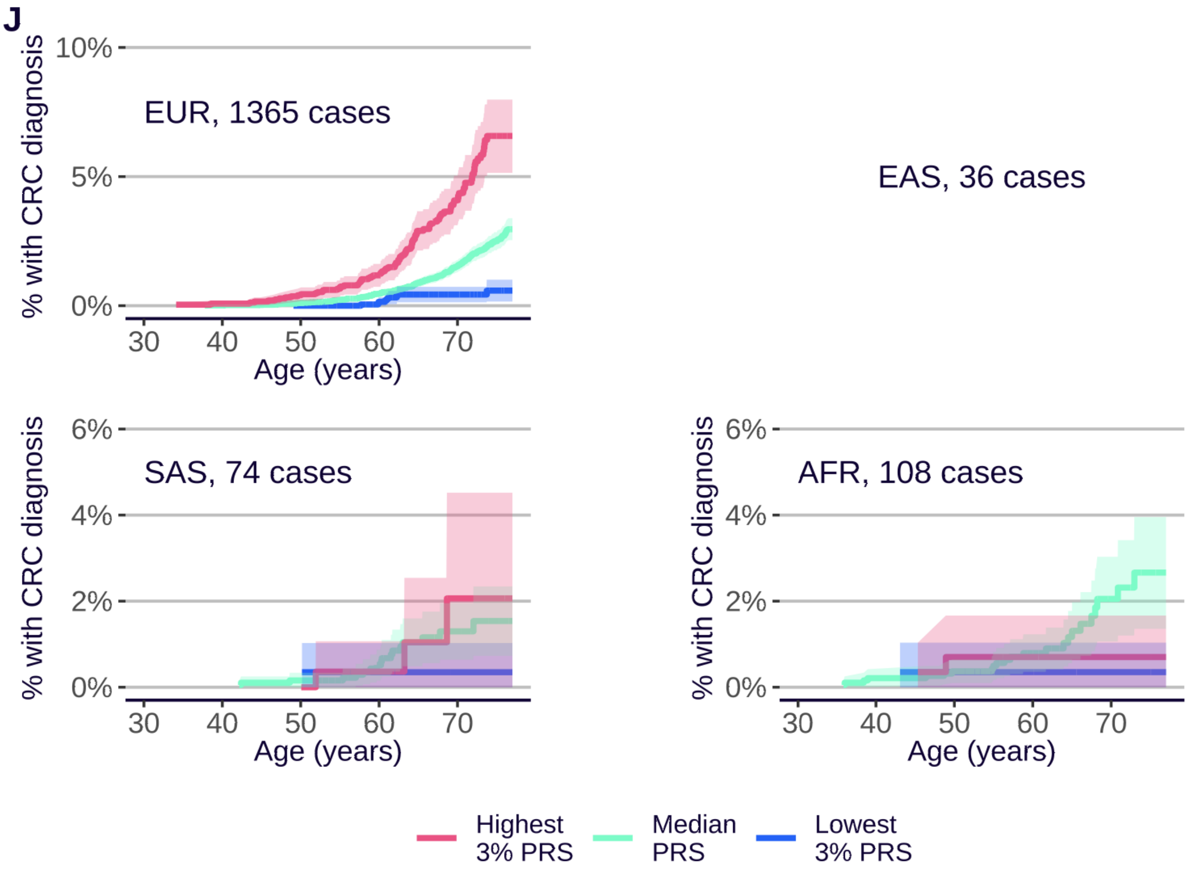
J
% with CRC diagnosis
10%
5%
0%
EUR, 1365 cases
30 40 50 60 70
Age (years)
EAS, 36 cases
6%
4%
2%
0%
SAS, 74 cases
AFR, 108 cases
Highest 3% PRS
Median PRS
Lowest 3% PRS

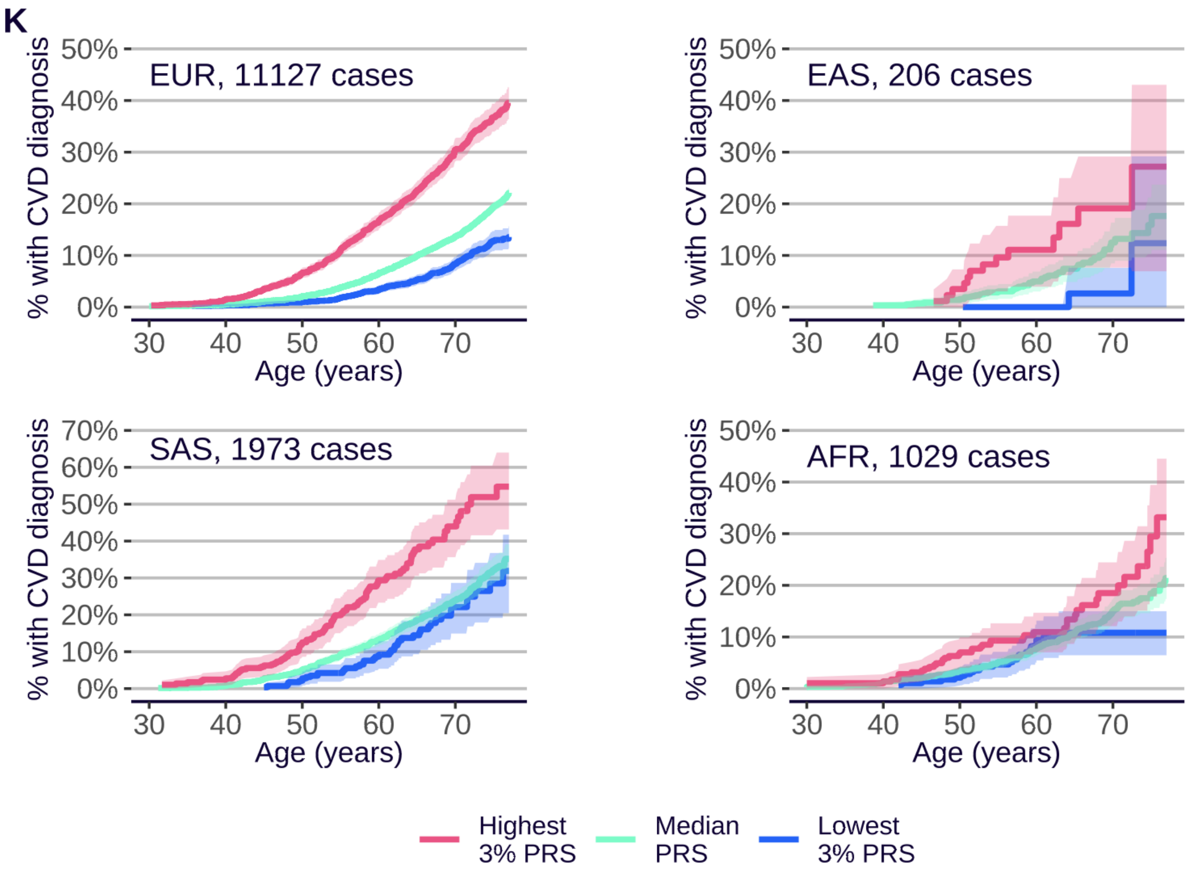
K
EUR, 11127 cases
EAS, 206 cases
SAS, 1973 cases
AFR, 1029 cases
% with CVD diagnosis
0%
10%
20%
30%
40%
50%
60%
70%
30
40
50
60
70
Age (years)
Highest
3% PRS
Median
PRS
Lowest
3% PRS

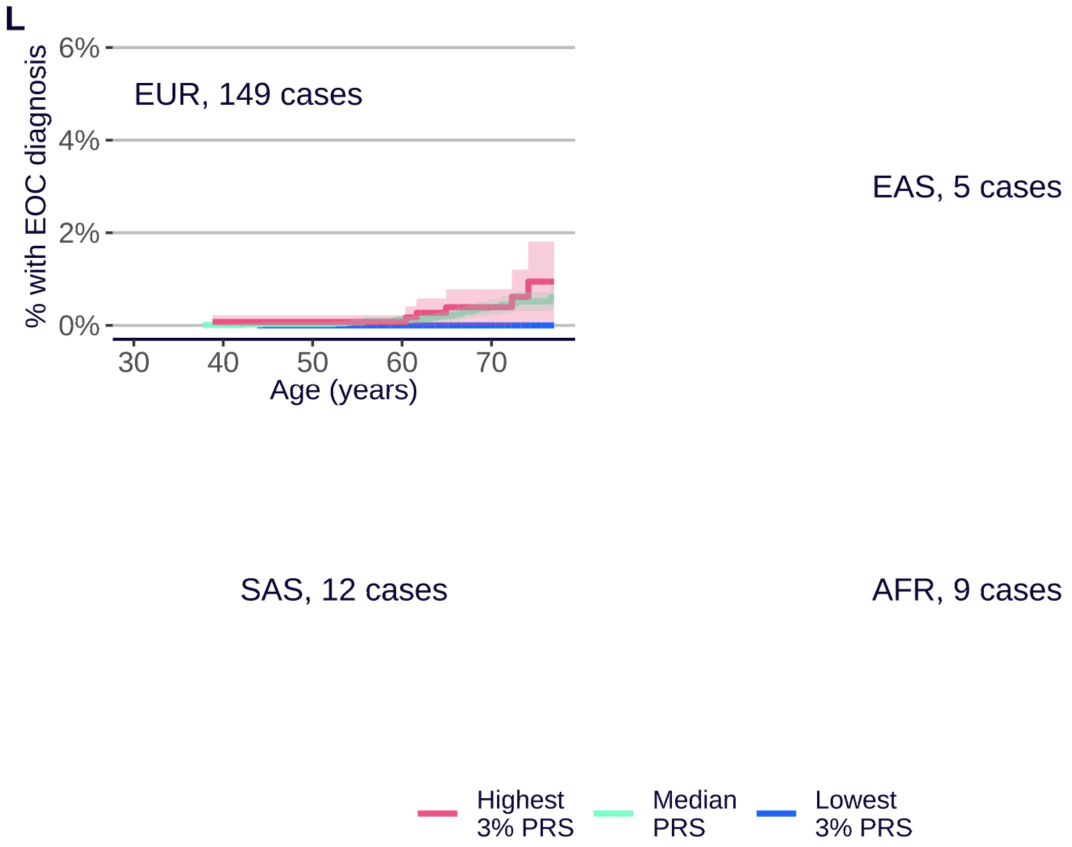
L
EUR, 149 cases
EAS, 5 cases
SAS, 12 cases
AFR, 9 cases
% with EOC diagnosis
0%
2%
4%
6%
30
40
50
60
70
Age (years)
Highest
3% PRS
Median
PRS
Lowest
3% PRS

M

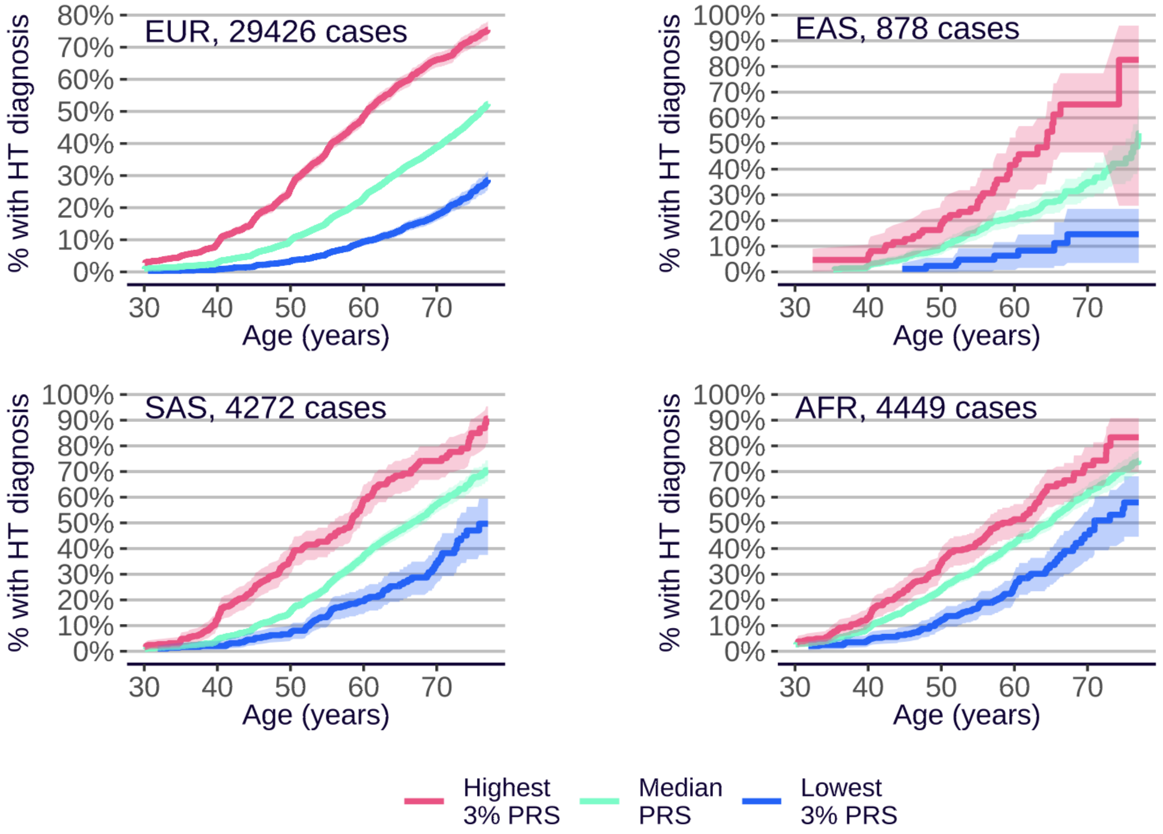
EUR, 29426 cases
EAS, 878 cases
SAS, 4272 cases
AFR, 4449 cases
% with HT diagnosis
Age (years)
Highest 3% PRS
Median PRS
Lowest 3% PRS

N

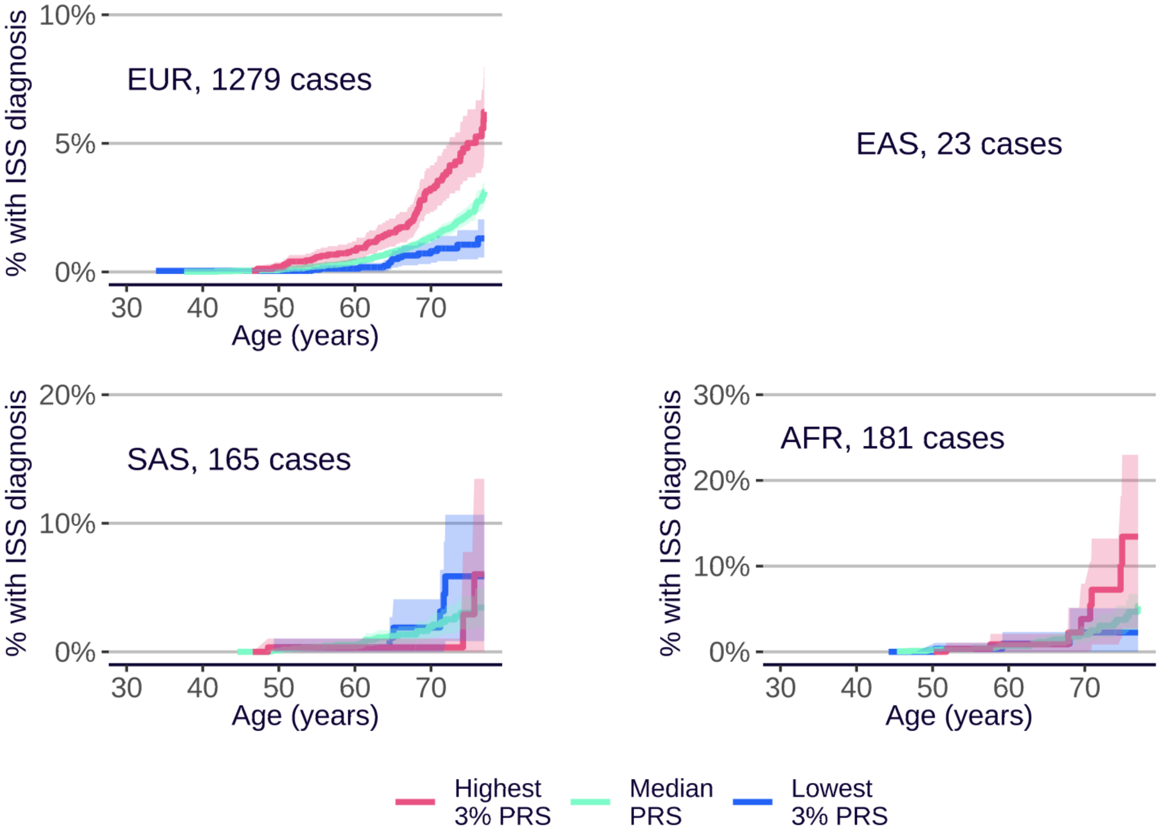
EUR, 1279 cases
EAS, 23 cases
SAS, 165 cases
AFR, 181 cases
% with ISS diagnosis
Age (years)
Highest 3% PRS
Median PRS
Lowest 3% PRS

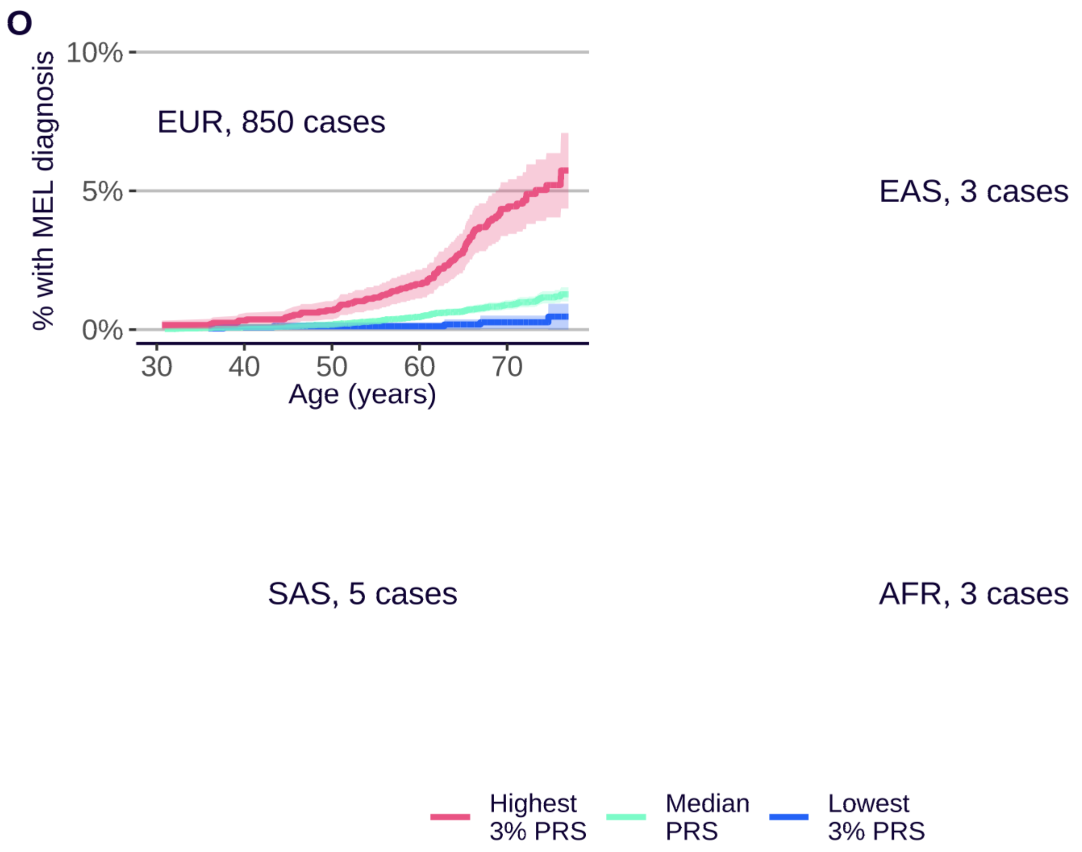
O
% with MEL diagnosis
10%
5%
0%
EUR, 850 cases
30
40
50
60
70
Age (years)
EAS, 3 cases
SAS, 5 cases
AFR, 3 cases
Highest 3% PRS
Median PRS
Lowest 3% PRS

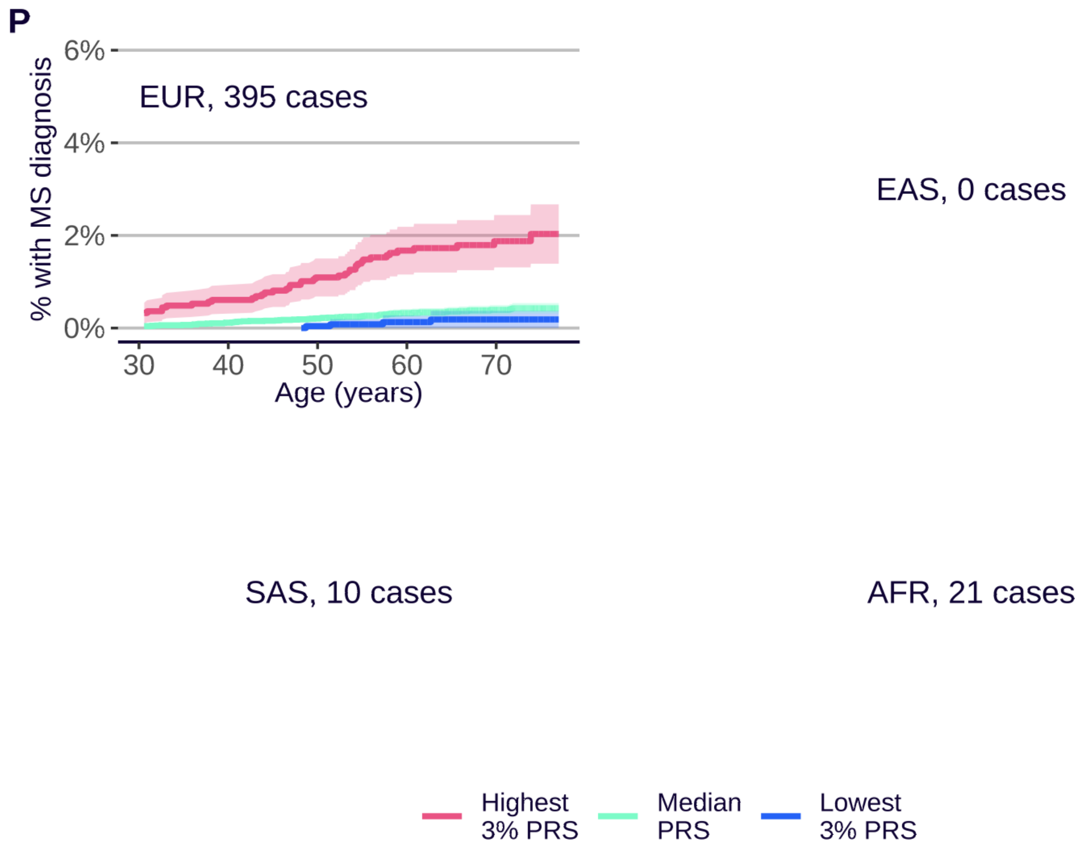
P
% with MS diagnosis
6%
4%
2%
0%
EUR, 395 cases
30
40
50
60
70
Age (years)
EAS, 0 cases
SAS, 10 cases
AFR, 21 cases
Highest 3% PRS
Median PRS
Lowest 3% PRS

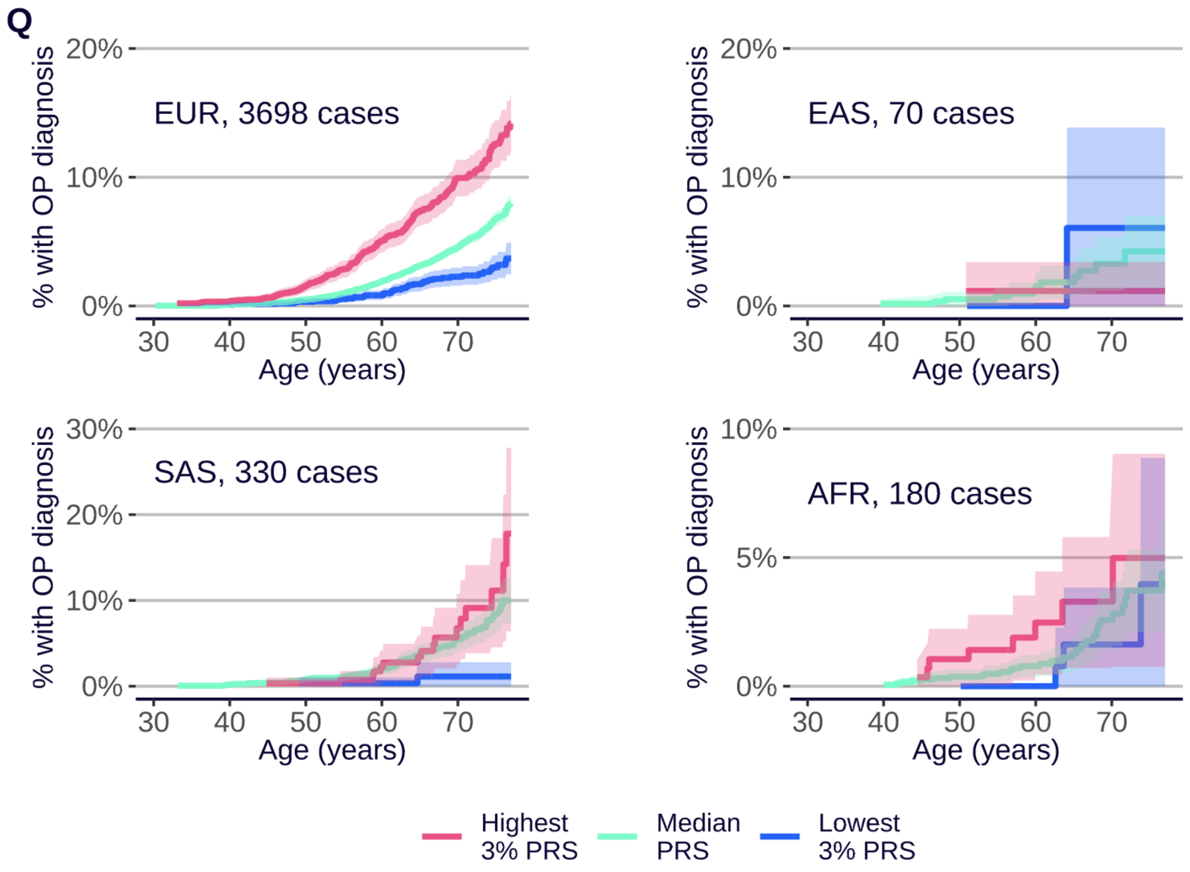
Q
EUR, 3698 cases
EAS, 70 cases
SAS, 330 cases
AFR, 180 cases
% with OP diagnosis
Age (years)
0%
5%
10%
20%
30%
30
40
50
60
70
Highest
3% PRS
Median
PRS
Lowest
3% PRS

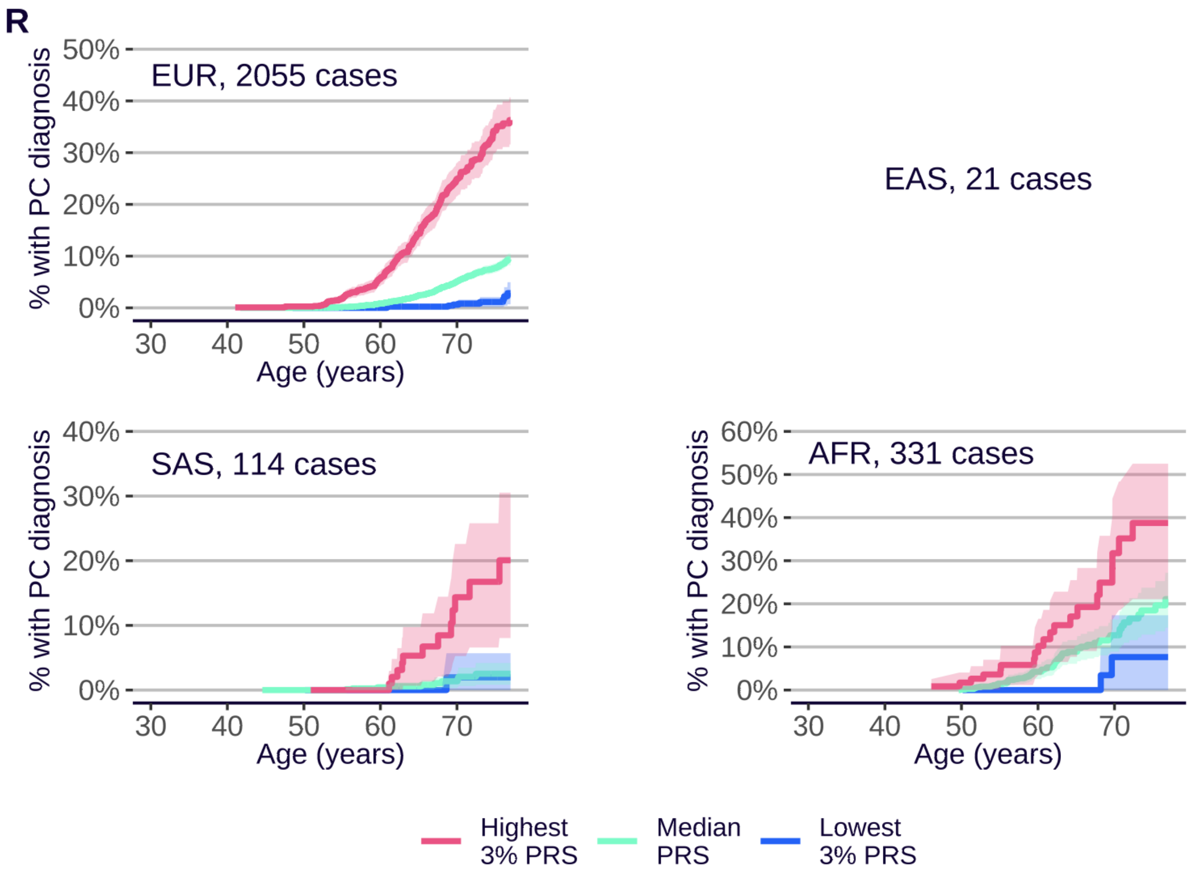
R
EUR, 2055 cases
EAS, 21 cases
SAS, 114 cases
AFR, 331 cases
% with PC diagnosis
Age (years)
0%
10%
20%
30%
40%
50%
60%
30
40
50
60
70
Highest
3% PRS
Median
PRS
Lowest
3% PRS

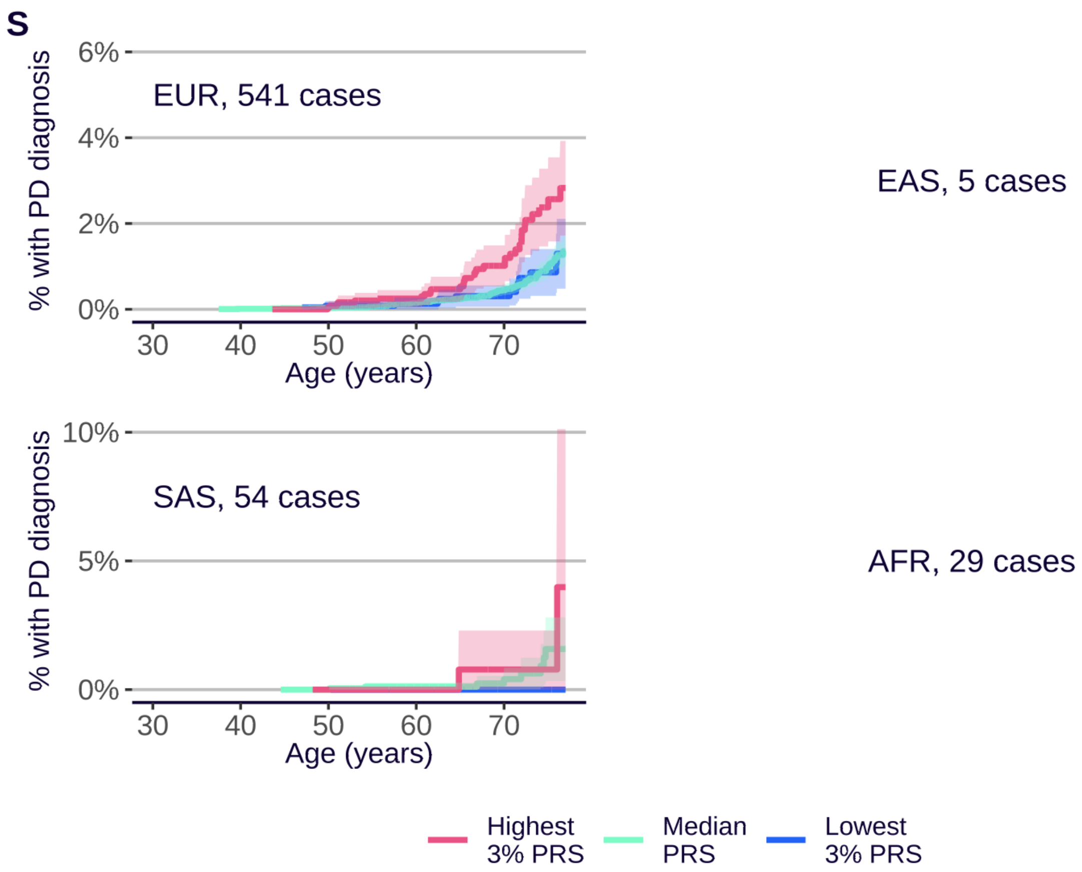
S
% with PD diagnosis
EUR, 541 cases
EAS, 5 cases
SAS, 54 cases
AFR, 29 cases
6%
4%
2%
0%
10%
5%
30
40
50
60
70
Age (years)
Highest
3% PRS
Median
PRS
Lowest
3% PRS

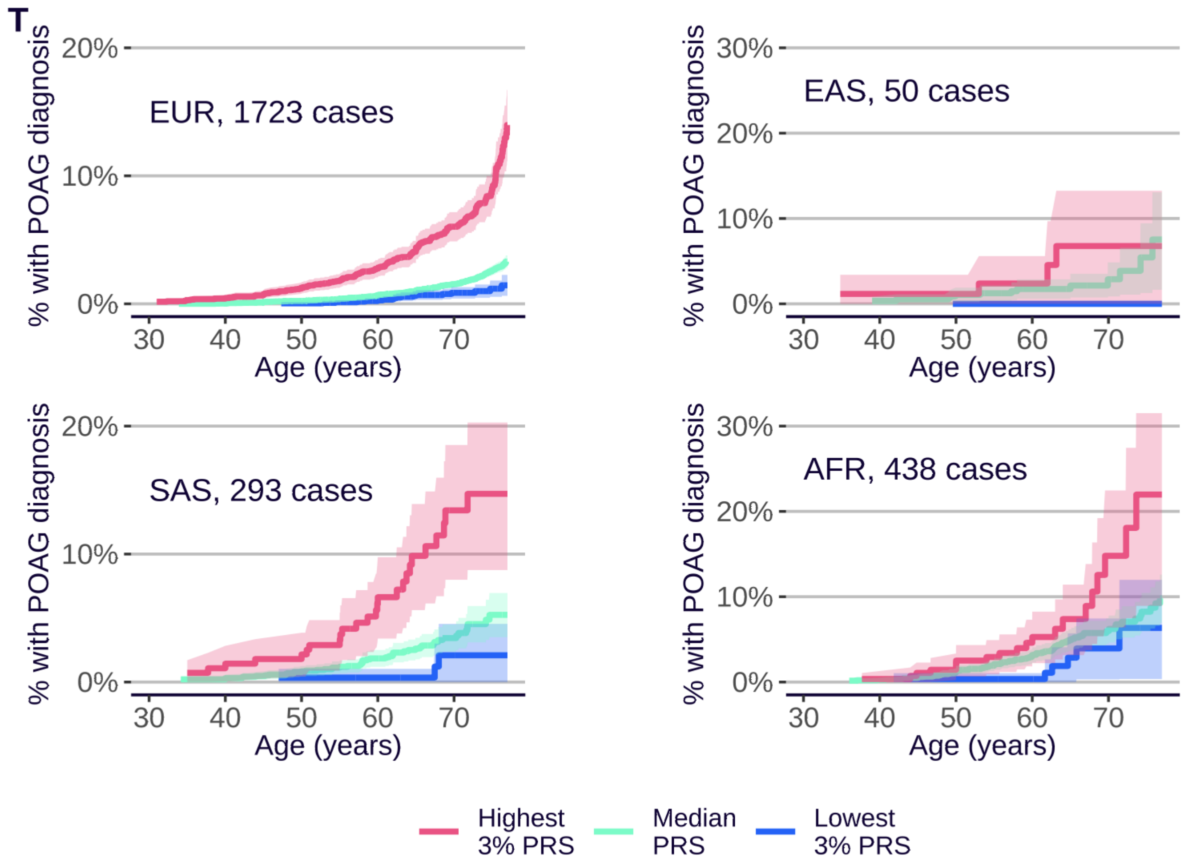
T
% with POAG diagnosis
EUR, 1723 cases
EAS, 50 cases
SAS, 293 cases
AFR, 438 cases
30%
20%
10%
0%
30
40
50
60
70
Age (years)
Highest
3% PRS
Median
PRS
Lowest
3% PRS

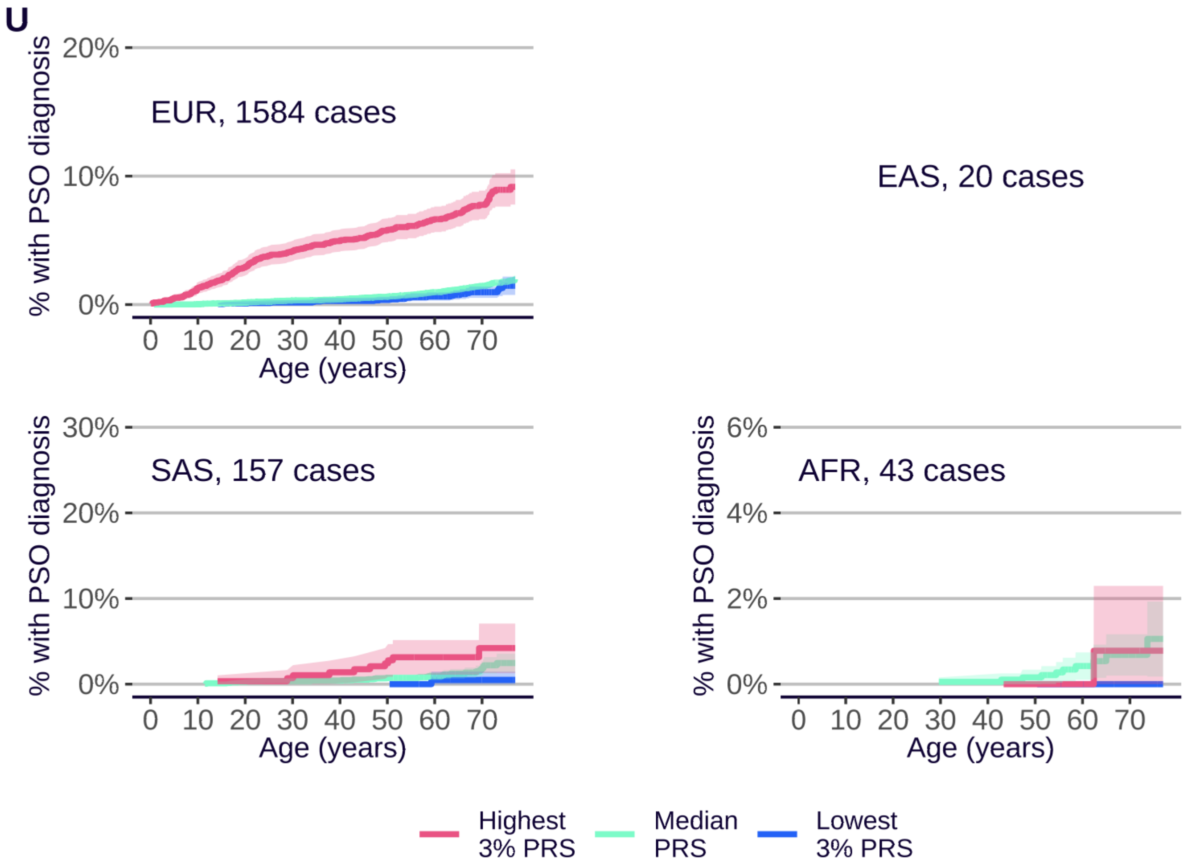
U
% with PSO diagnosis
20%
10%
0%
EUR, 1584 cases
0 10 20 30 40 50 60 70
Age (years)
EAS, 20 cases
30%
20%
10%
0%
SAS, 157 cases
6%
4%
2%
0%
AFR, 43 cases
Highest 3% PRS
Median PRS
Lowest 3% PRS

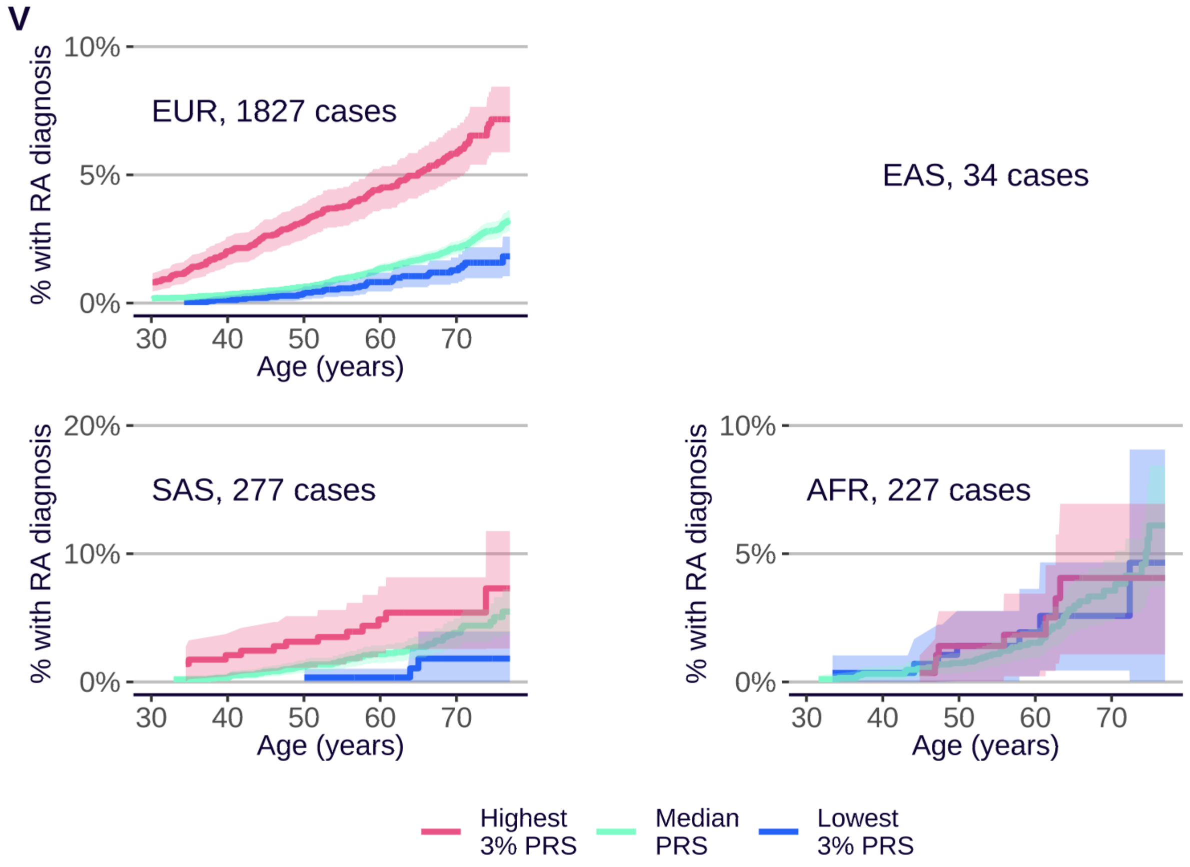
V
% with RA diagnosis
10%
5%
0%
EUR, 1827 cases
30 40 50 60 70
Age (years)
EAS, 34 cases
20%
10%
0%
SAS, 277 cases
10%
5%
0%
AFR, 227 cases
Highest 3% PRS
Median PRS
Lowest 3% PRS

W

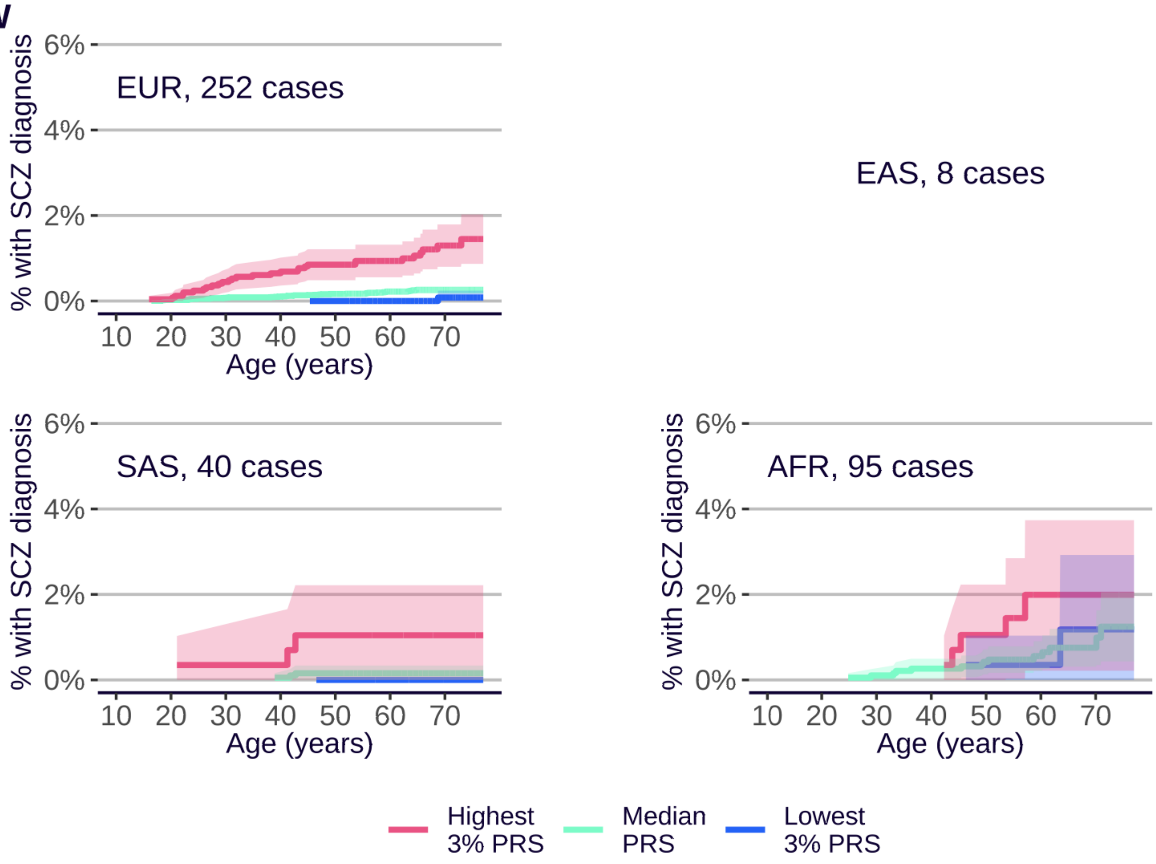

EUR, 252 cases
EAS, 8 cases
SAS, 40 cases
AFR, 95 cases
% with SCZ diagnosis
6%
4%
2%
0%
10 20 30 40 50 60 70
Age (years)
Highest 3% PRS
Median PRS
Lowest 3% PRS


X

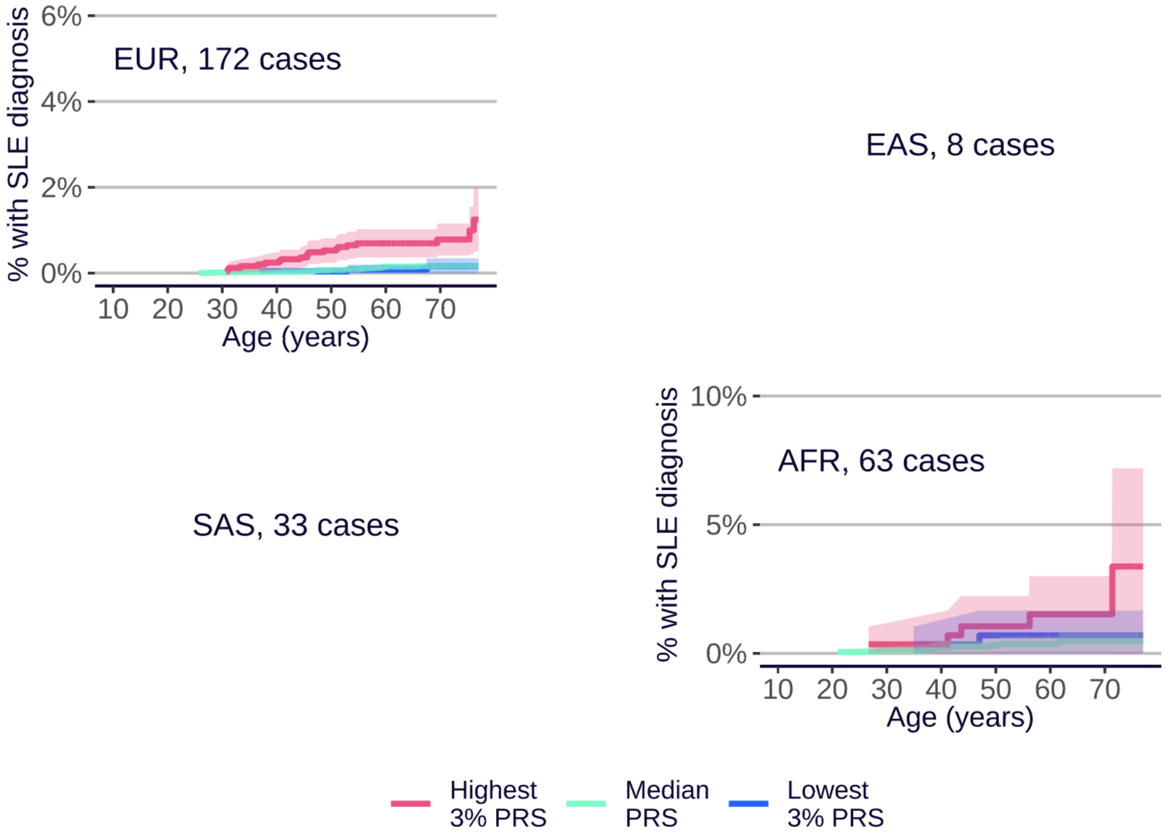

EUR, 172 cases
EAS, 8 cases
SAS, 33 cases
AFR, 63 cases
% with SLE diagnosis
6%
4%
2%
0%
10%
5%
10 20 30 40 50 60 70
Age (years)
Highest 3% PRS
Median PRS
Lowest 3% PRS

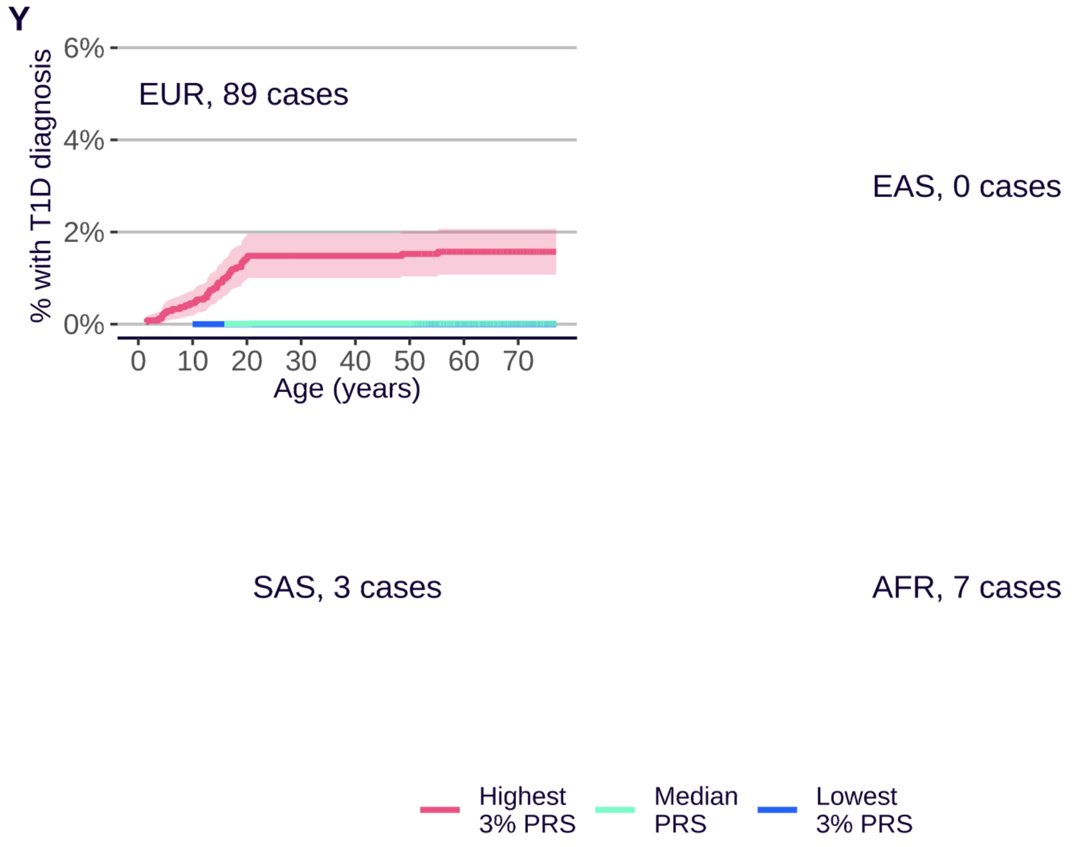
Y
% with T1D diagnosis
6%
4%
2%
0%
EUR, 89 cases
0 10 20 30 40 50 60 70
Age (years)
EAS, 0 cases
SAS, 3 cases
AFR, 7 cases
Highest 3% PRS
Median PRS
Lowest 3% PRS

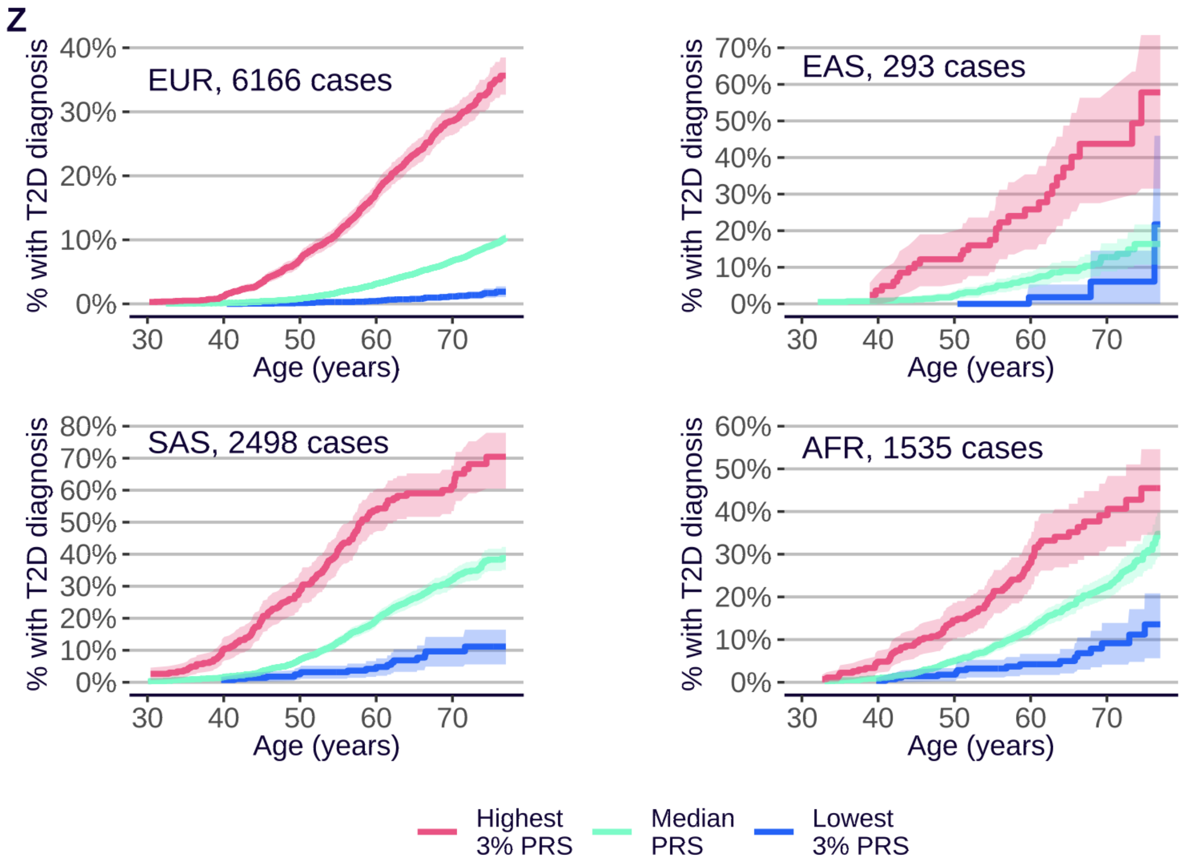
Z
% with T2D diagnosis
EUR, 6166 cases
40%
30%
20%
10%
0%
30 40 50 60 70
Age (years)
EAS, 293 cases
70%
60%
50%
40%
30%
20%
10%
0%
SAS, 2498 cases
80%
70%
60%
50%
40%
30%
20%
10%
0%
AFR, 1535 cases
60%
50%
40%
30%
20%
10%
0%
Highest 3% PRS
Median PRS
Lowest 3% PRS

AA

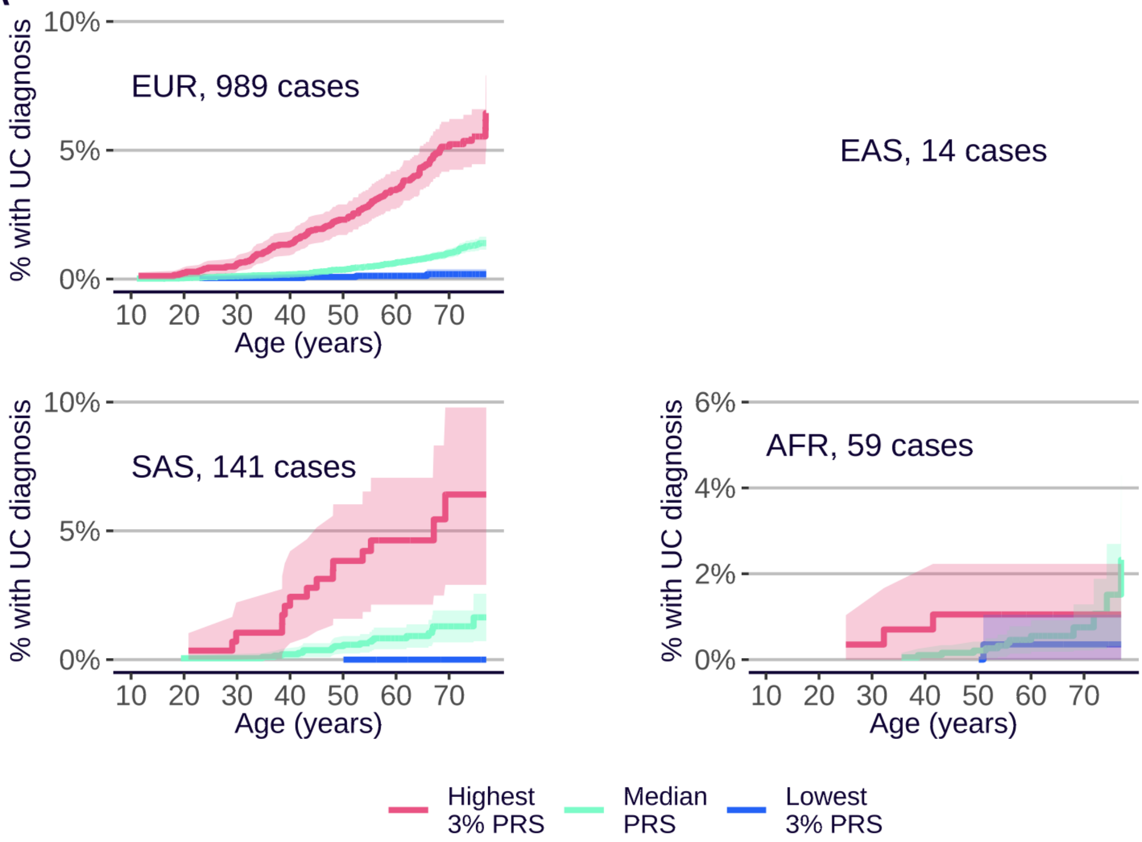
% with UC diagnosis
10%
5%
0%
EUR, 989 cases
10 20 30 40 50 60 70
Age (years)
EAS, 14 cases
SAS, 141 cases
AFR, 59 cases
6%
4%
2%
Highest 3% PRS
Median PRS
Lowest 3% PRS

AB

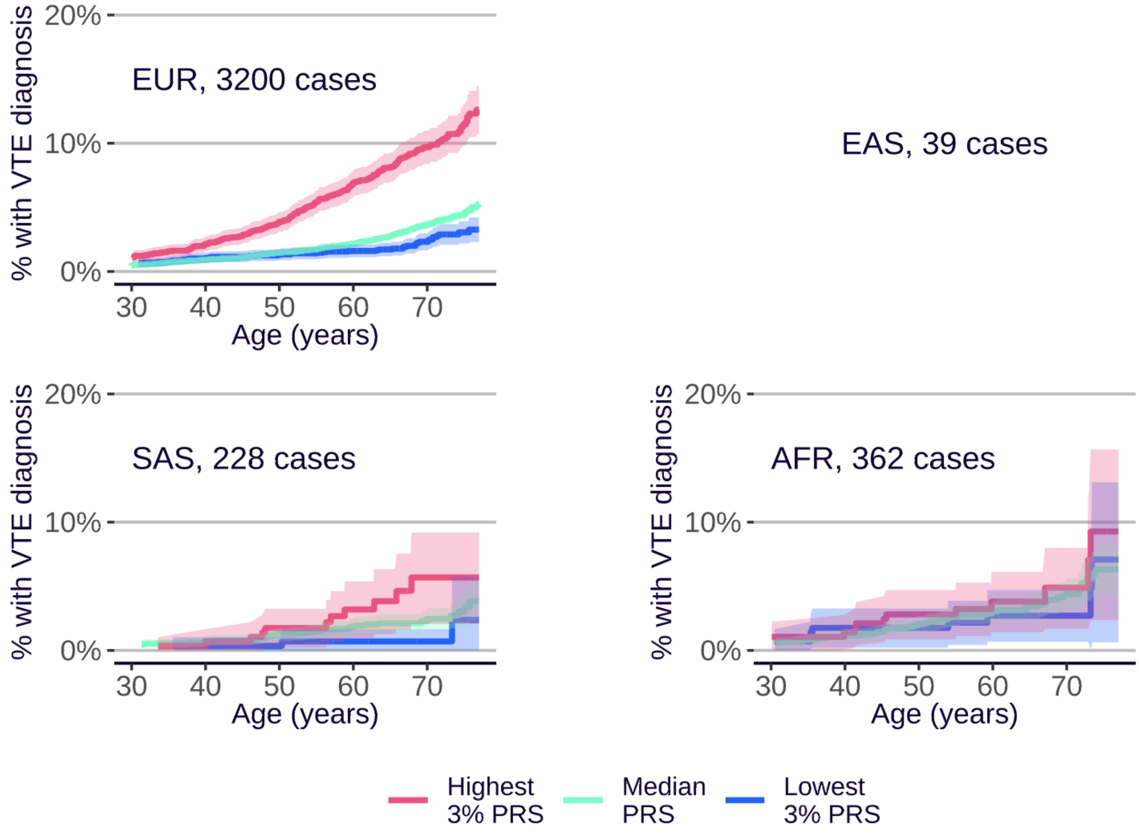
% with VTE diagnosis
20%
10%
0%
EUR, 3200 cases
30 40 50 60 70
Age (years)
EAS, 39 cases
SAS, 228 cases
AFR, 362 cases
Highest 3% PRS
Median PRS
Lowest 3% PRS

**Supplementary Figure 10**. **Cumulative incidence plots by ancestry, Standard PRS Set**. Cumulative incidence plots are shown for each disease and each ancestry group in the UKB Testing Subgroup, provided more than 40 cases are available (the number of cases is printed otherwise), with separate curves for the highest 3% (red), lowest 3% (blue), and median (green) of the PRS distribution. **A**. Alzheimer's disease (AD). **B**. Atrial fibrillation (AF). **C**. Age-related macular degeneration (AMD). **D**. Asthma (AST). **E**. Breast cancer (BC), **F**. Bipolar disorder (BD), **G**. Coronary artery disease (CAD). **H**. Crohn's disease (CD). **I**. Coeliac disease (CED). **J**. Bowel cancer (CRC). **K**. Cardiovascular disease (CVD), **L**. Epithelial ovarian cancer (EOC). **M**. Hypertension (HT). **N**. Ischaemic stroke (ISS). **O**. Melanoma (MEL). **P**. Multiple sclerosis (MS). **Q**. Osteoporosis (OP). **R**. Prostate cancer (PC). **S**. Parkinson's disease (PD). **T**. Primary open angle glaucoma (POAG). **U**. Psoriasis (PSO). **V**. Rheumatoid arthritis (RA). **W**. Schizophrenia (SCZ). **X**. Systemic lupus erythematosus (SLE). **Y**. Type 1 diabetes (T1D). **Z**. Type 2 diabetes (T2D). **AA**. Ulcerative colitis (UC). **AB**. Venous thromboembolic disease (VTE). EUR = European ancestry. EAS = East Asian ancestry. SAS = South Asian ancestry. AFR = Sub-Saharan African ancestry. Shaded areas indicate 95% CI.

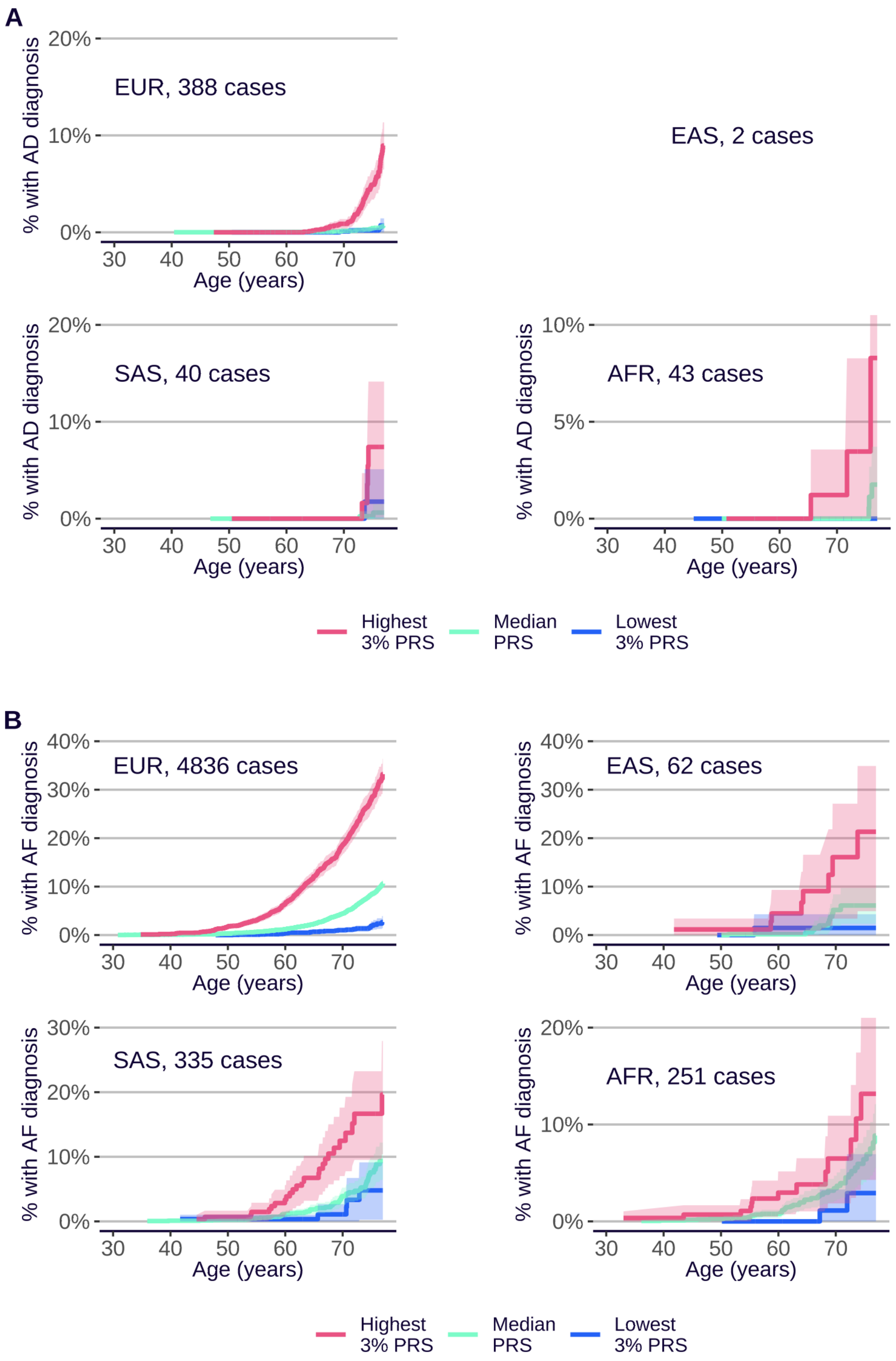
A
% with AD diagnosis
EUR, 388 cases
EAS, 2 cases
SAS, 40 cases
AFR, 43 cases
20%
10%
5%
0%
30 40 50 60 70
Age (years)
Highest 3% PRS
Median PRS
Lowest 3% PRS
B
% with AF diagnosis
EUR, 4836 cases
EAS, 62 cases
SAS, 335 cases
AFR, 251 cases
40%
30%
20%
10%
0%
30 40 50 60 70
Age (years)
Highest 3% PRS
Median PRS
Lowest 3% PRS

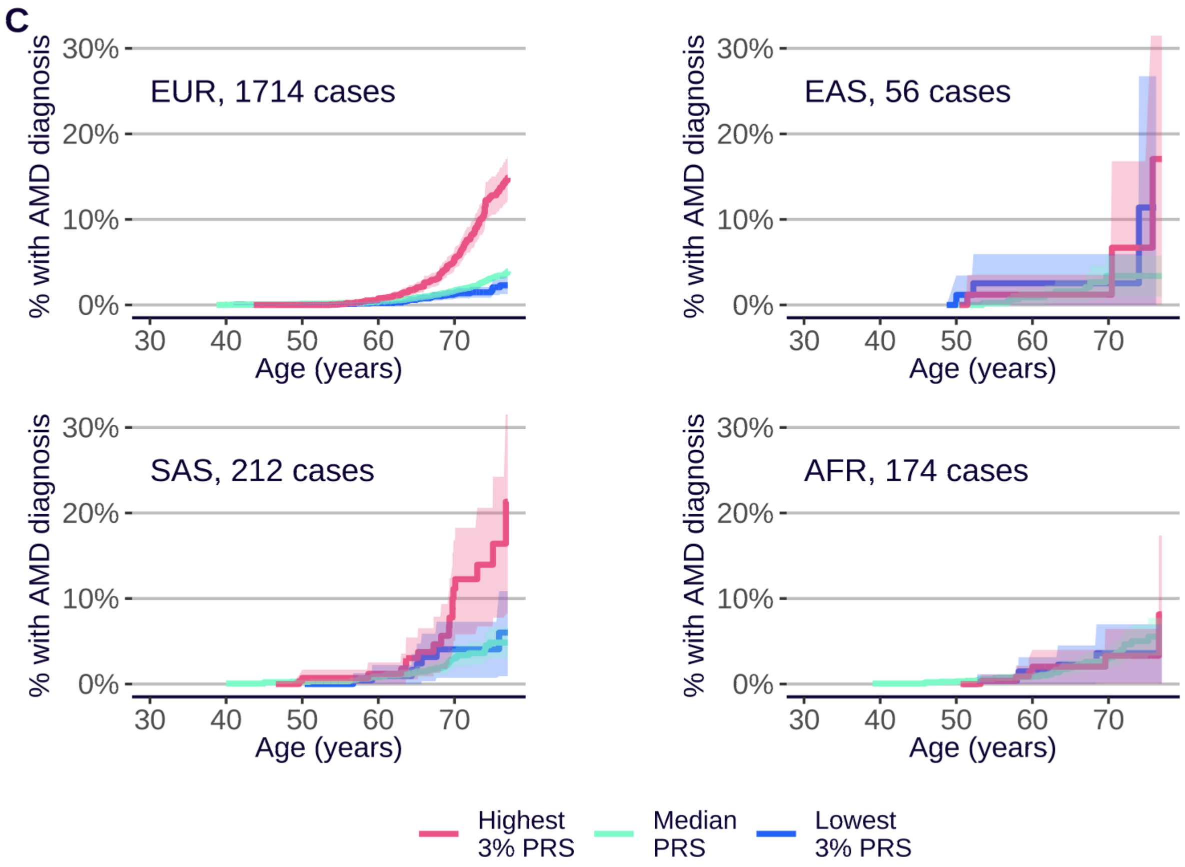
C
EUR, 1714 cases
EAS, 56 cases
SAS, 212 cases
AFR, 174 cases
% with AMD diagnosis
0%
10%
20%
30%
Age (years)
30
40
50
60
70
Highest 3% PRS
Median PRS
Lowest 3% PRS

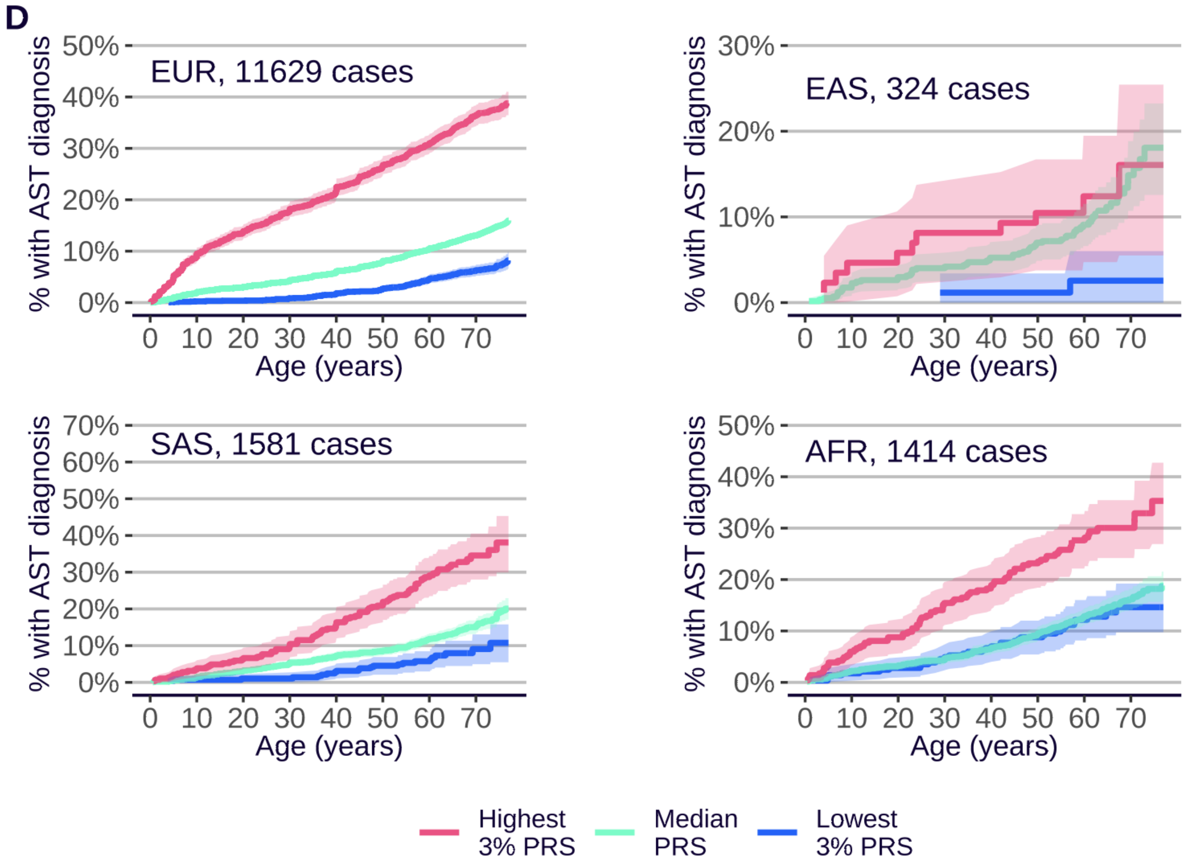
D
EUR, 11629 cases
EAS, 324 cases
SAS, 1581 cases
AFR, 1414 cases
% with AST diagnosis
0%
10%
20%
30%
40%
50%
60%
70%
Age (years)
0
10
20
30
40
50
60
70
Highest 3% PRS
Median PRS
Lowest 3% PRS

E

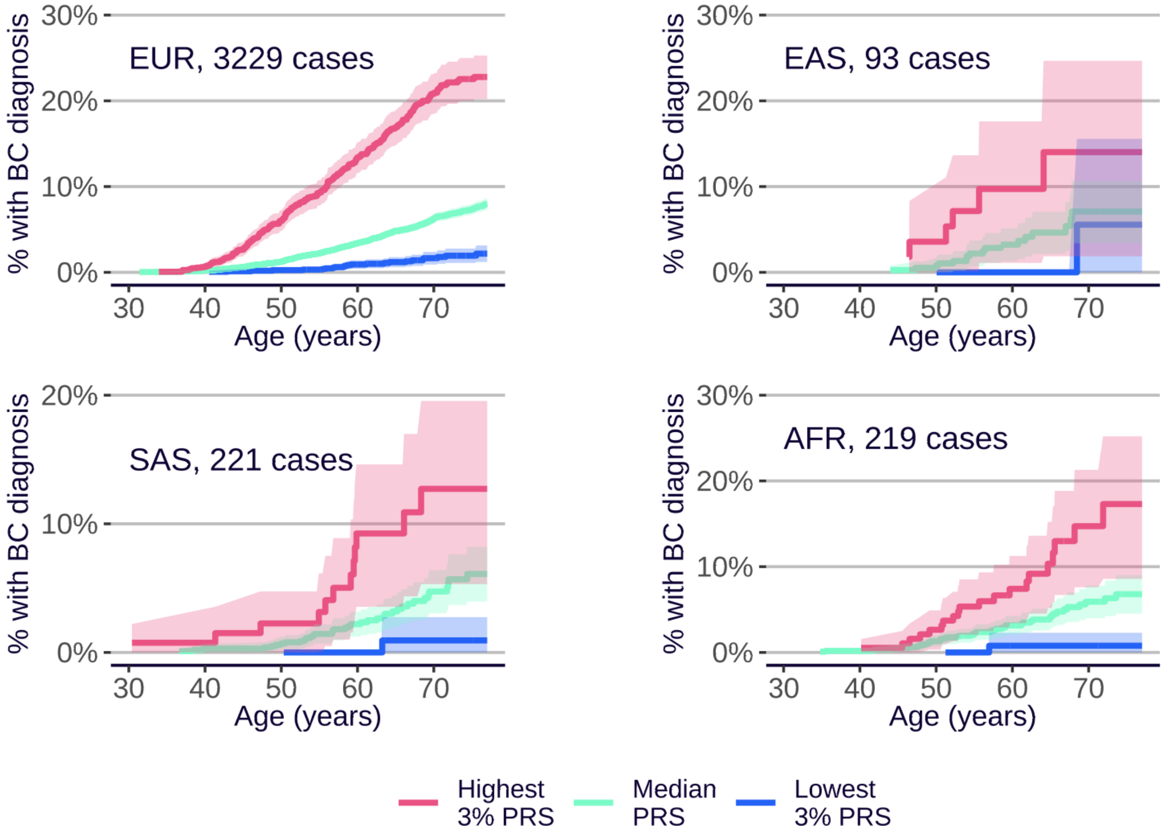
EUR, 3229 cases
EAS, 93 cases
SAS, 221 cases
AFR, 219 cases
% with BC diagnosis
Age (years)
30%
20%
10%
0%
30
40
50
60
70
Highest 3% PRS
Median PRS
Lowest 3% PRS

F

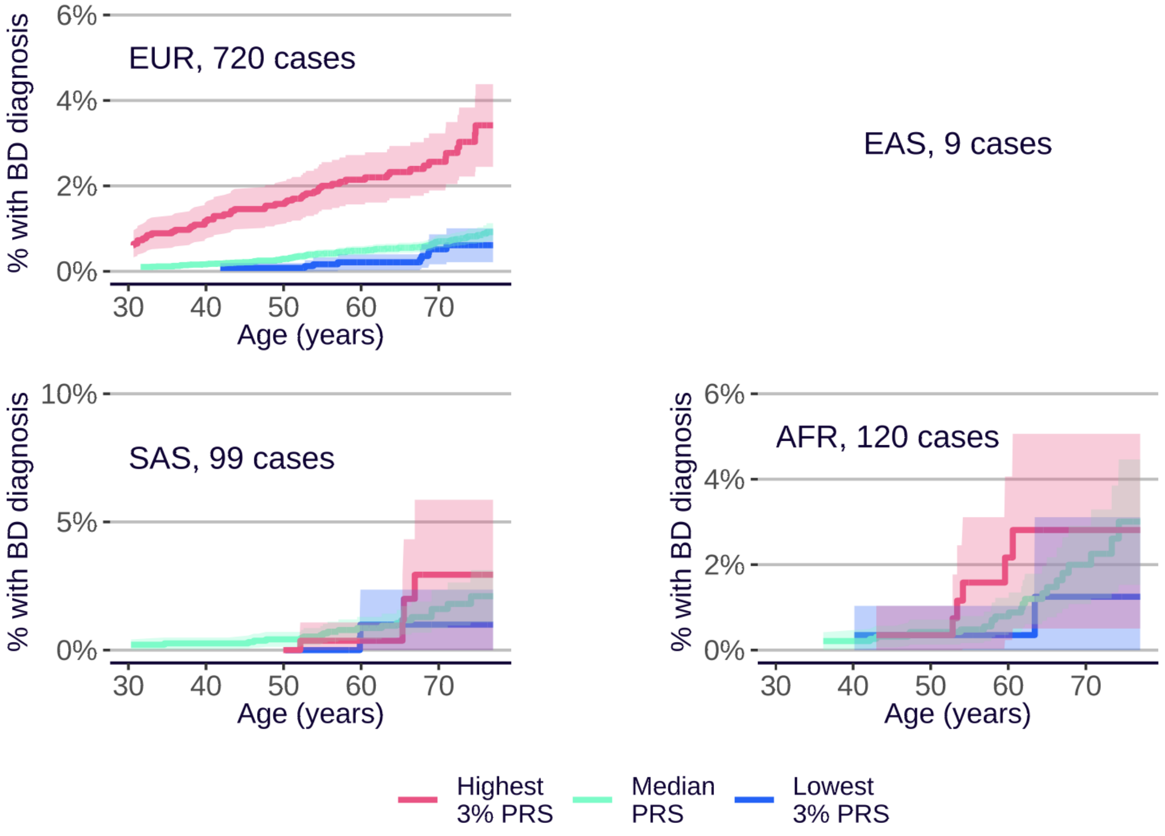
EUR, 720 cases
EAS, 9 cases
SAS, 99 cases
AFR, 120 cases
% with BD diagnosis
Age (years)
10%
6%
5%
4%
2%
0%
30
40
50
60
70
Highest 3% PRS
Median PRS
Lowest 3% PRS

**G**

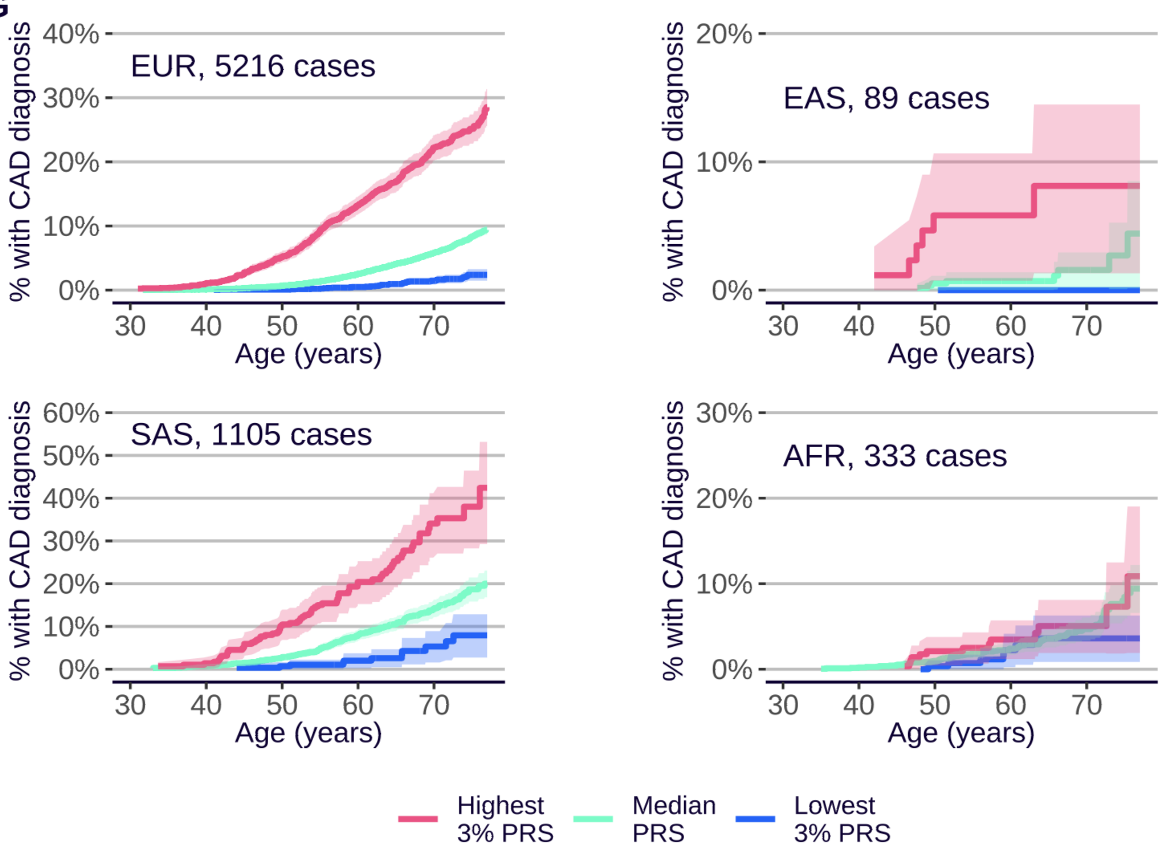

EUR, 5216 cases
EAS, 89 cases
SAS, 1105 cases
AFR, 333 cases
% with CAD diagnosis
Age (years)
40%
30%
20%
10%
0%
60%
50%
30
40
50
60
70
Highest 3% PRS
Median PRS
Lowest 3% PRS


**H**

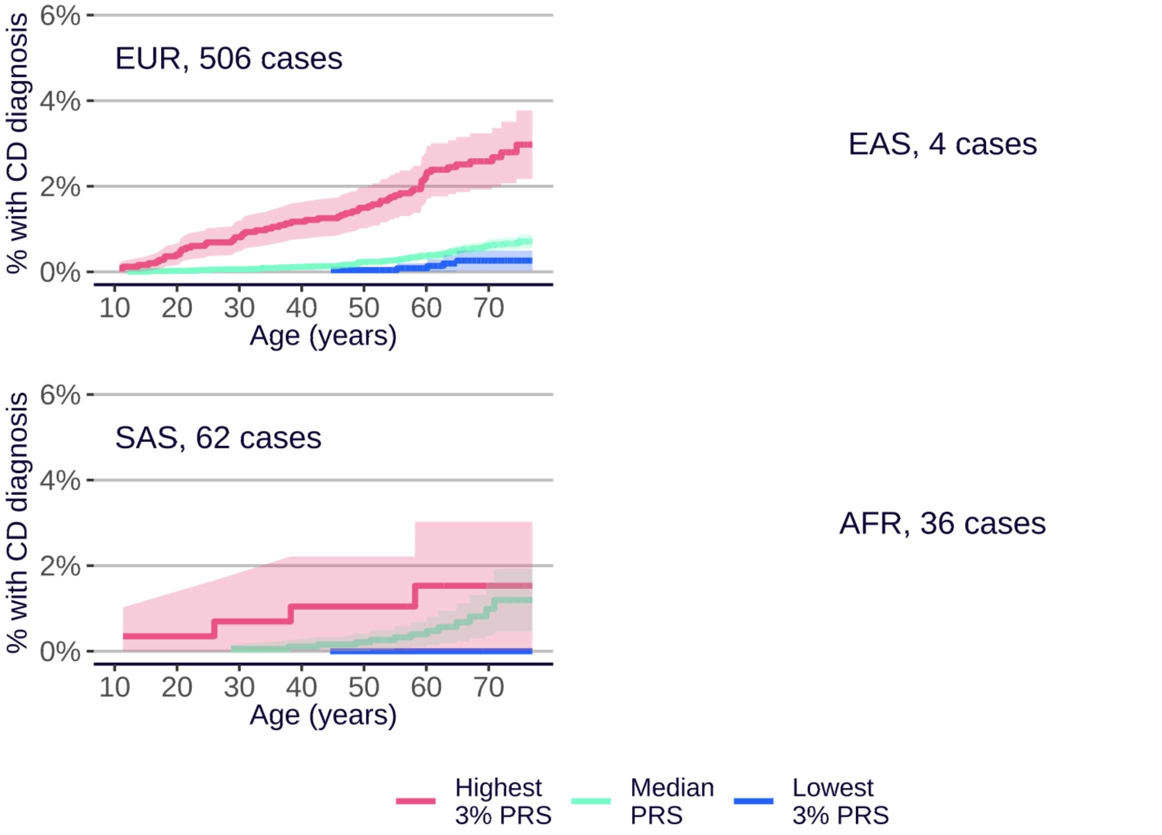

EUR, 506 cases
EAS, 4 cases
SAS, 62 cases
AFR, 36 cases
% with CD diagnosis
Age (years)
6%
4%
2%
0%
10
20
30
40
50
60
70
Highest 3% PRS
Median PRS
Lowest 3% PRS

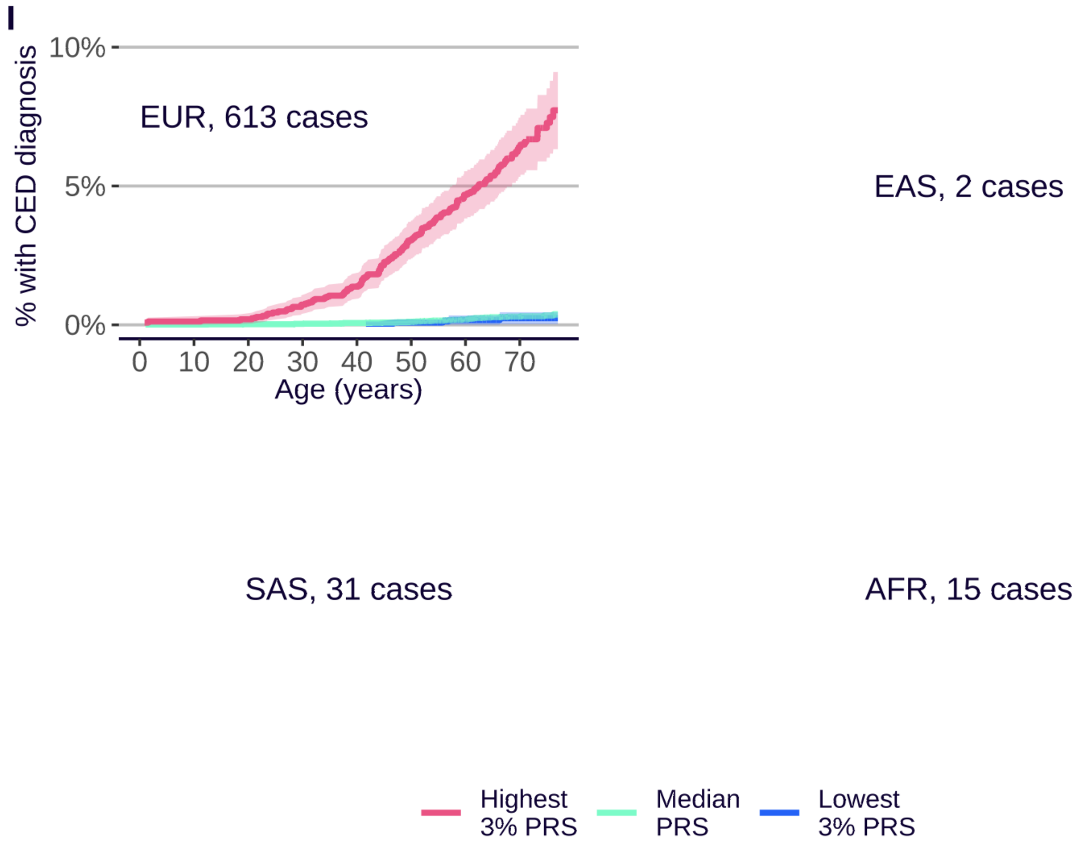
I
% with CED diagnosis
10%
5%
0%
EUR, 613 cases
0 10 20 30 40 50 60 70
Age (years)
EAS, 2 cases
SAS, 31 cases
AFR, 15 cases
Highest 3% PRS
Median PRS
Lowest 3% PRS

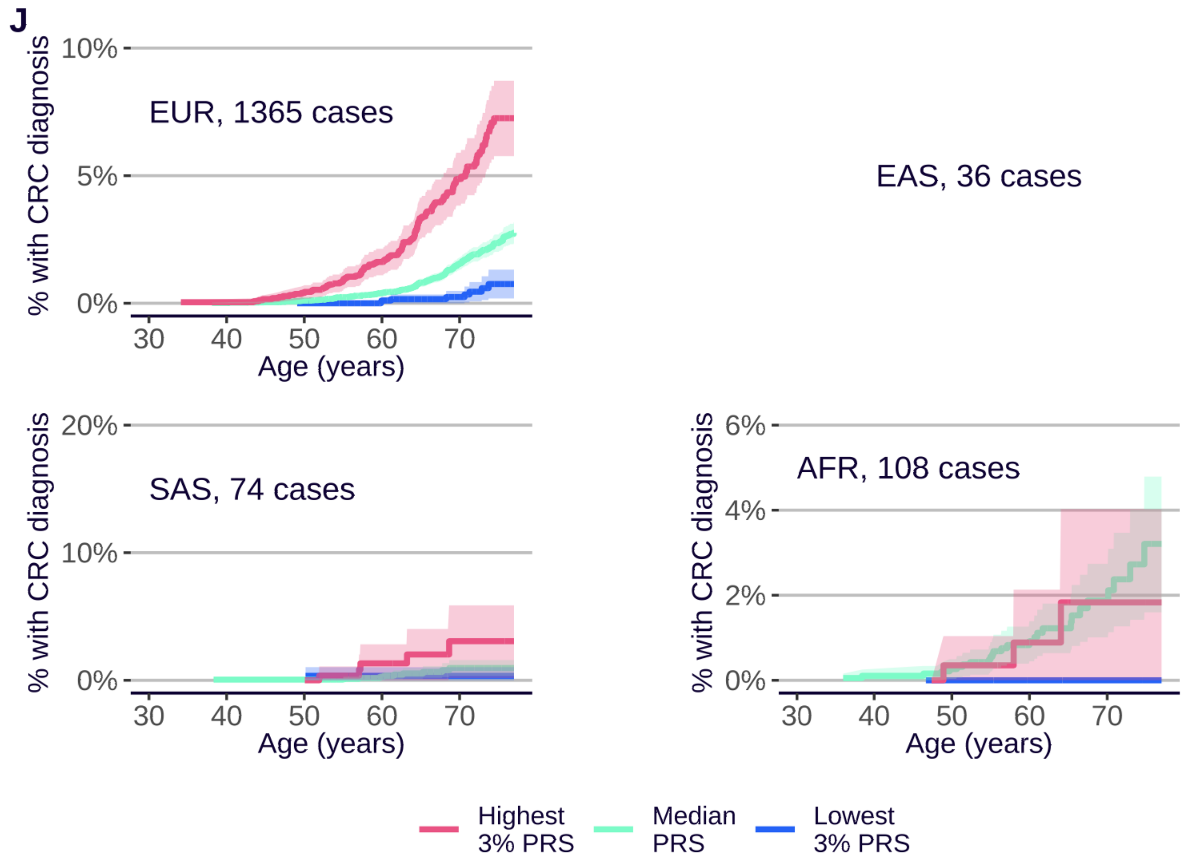
J
% with CRC diagnosis
10%
5%
0%
EUR, 1365 cases
30 40 50 60 70
Age (years)
EAS, 36 cases
20%
10%
0%
SAS, 74 cases
6%
4%
2%
0%
AFR, 108 cases
Highest 3% PRS
Median PRS
Lowest 3% PRS

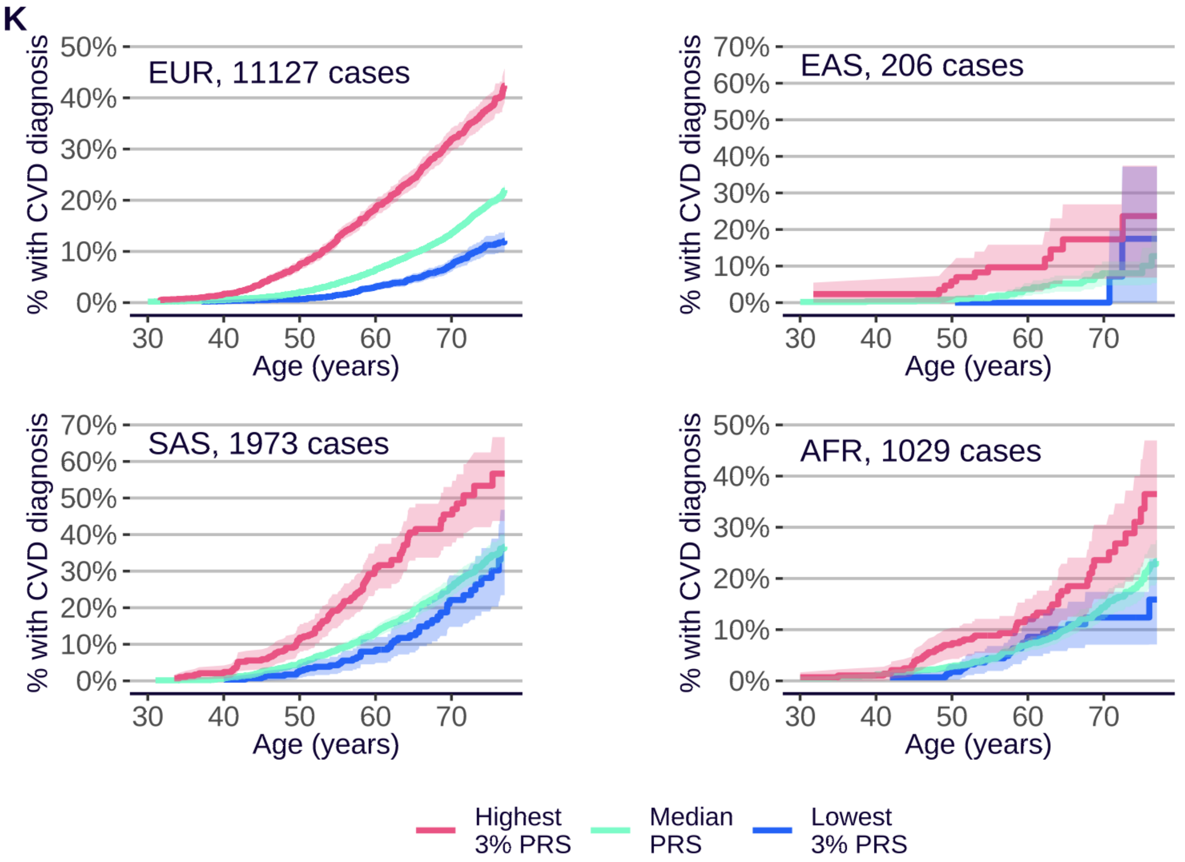
K
EUR, 11127 cases
EAS, 206 cases
SAS, 1973 cases
AFR, 1029 cases
% with CVD diagnosis
Age (years)
Highest 3% PRS
Median PRS
Lowest 3% PRS

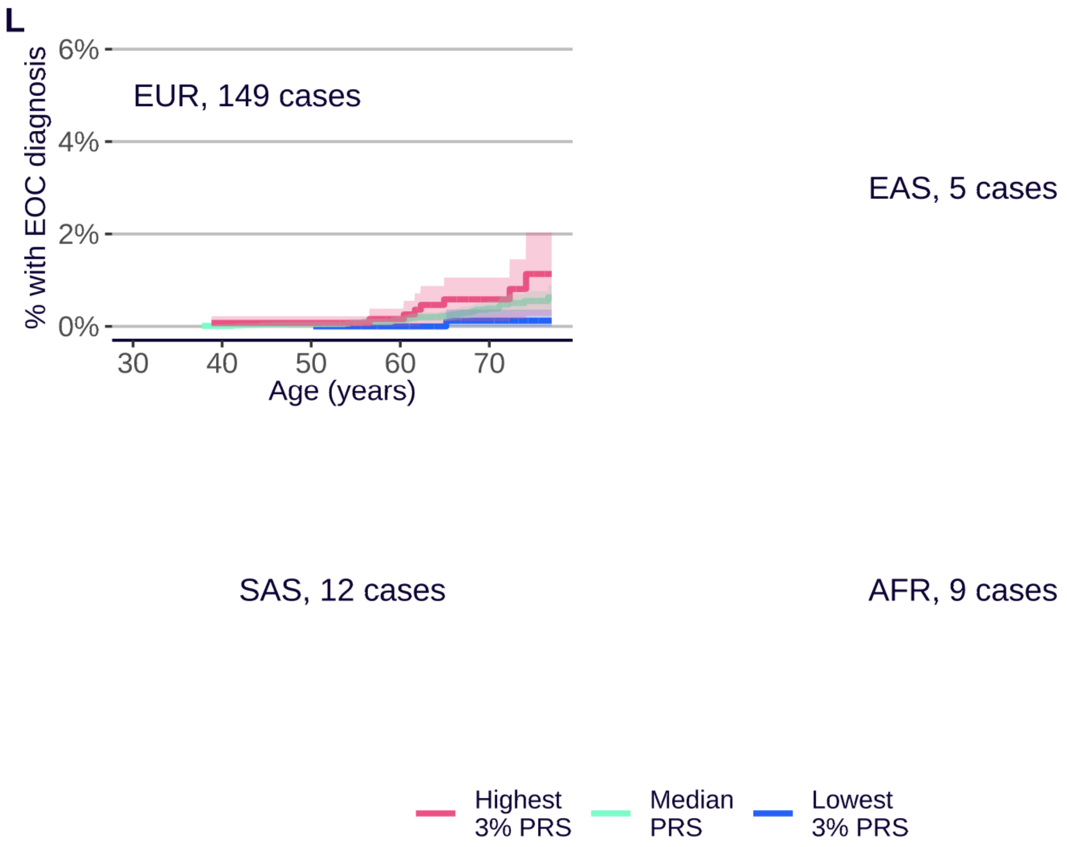
L
EUR, 149 cases
EAS, 5 cases
SAS, 12 cases
AFR, 9 cases
% with EOC diagnosis
Age (years)
Highest 3% PRS
Median PRS
Lowest 3% PRS

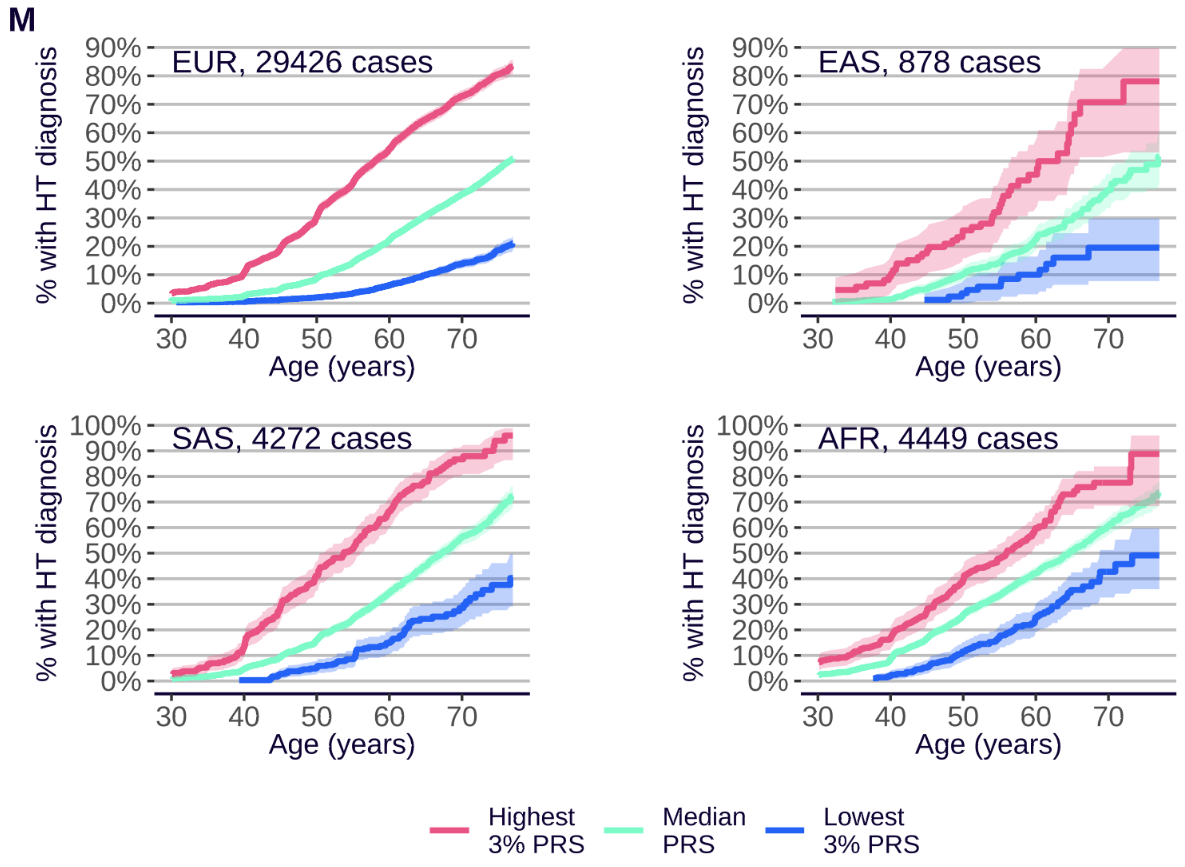
M
EUR, 29426 cases
EAS, 878 cases
SAS, 4272 cases
AFR, 4449 cases
% with HT diagnosis
Age (years)
0%
10%
20%
30%
40%
50%
60%
70%
80%
90%
100%
30
40
50
60
70
Highest 3% PRS
Median PRS
Lowest 3% PRS

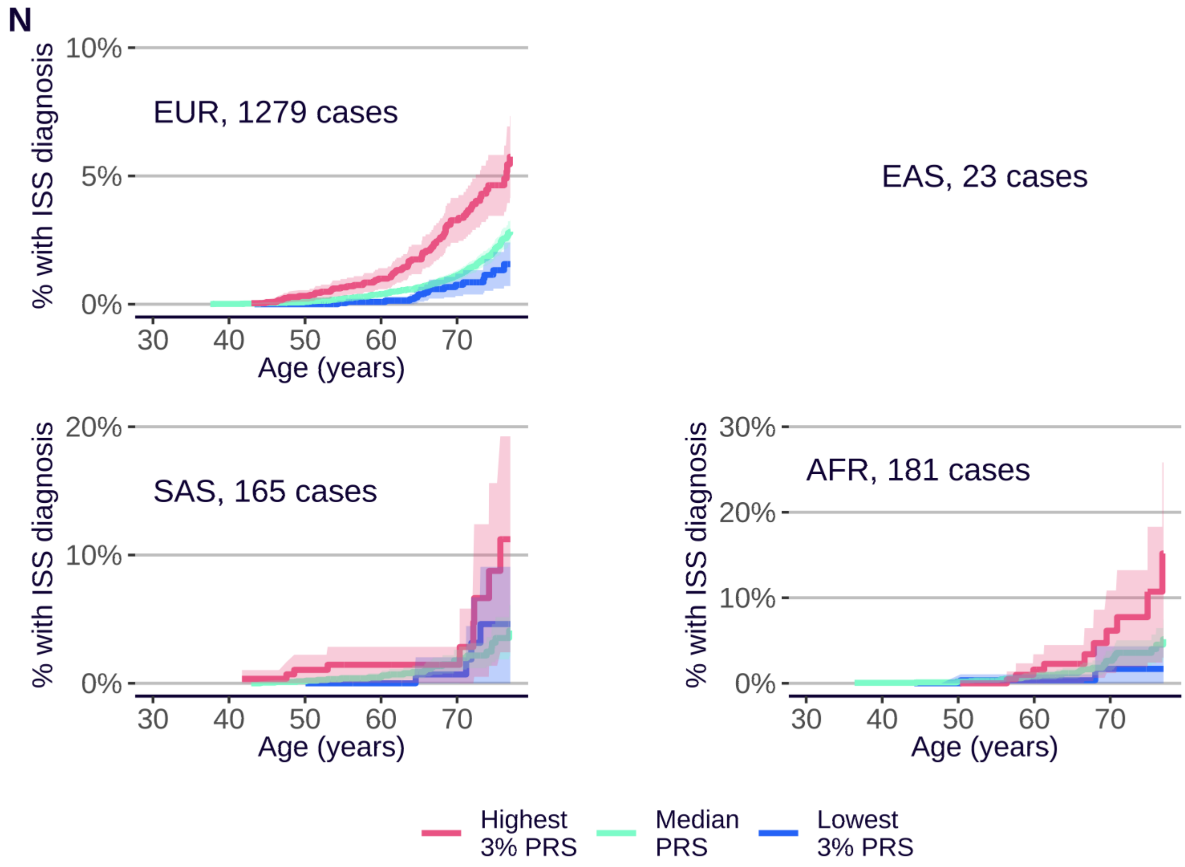
N
EUR, 1279 cases
EAS, 23 cases
SAS, 165 cases
AFR, 181 cases
% with ISS diagnosis
Age (years)
0%
5%
10%
20%
30%
30
40
50
60
70
Highest 3% PRS
Median PRS
Lowest 3% PRS

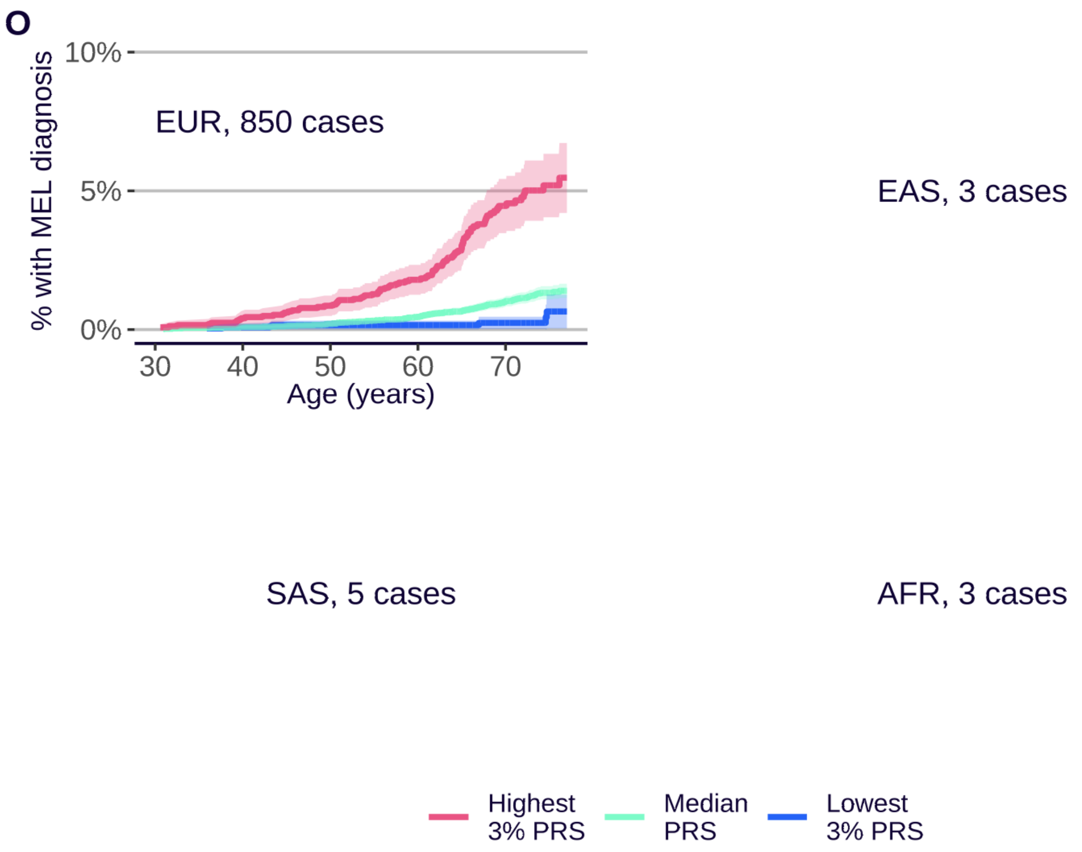
O
% with MEL diagnosis
10%
5%
0%
EUR, 850 cases
30
40
50
60
70
Age (years)
EAS, 3 cases
SAS, 5 cases
AFR, 3 cases
Highest 3% PRS
Median PRS
Lowest 3% PRS

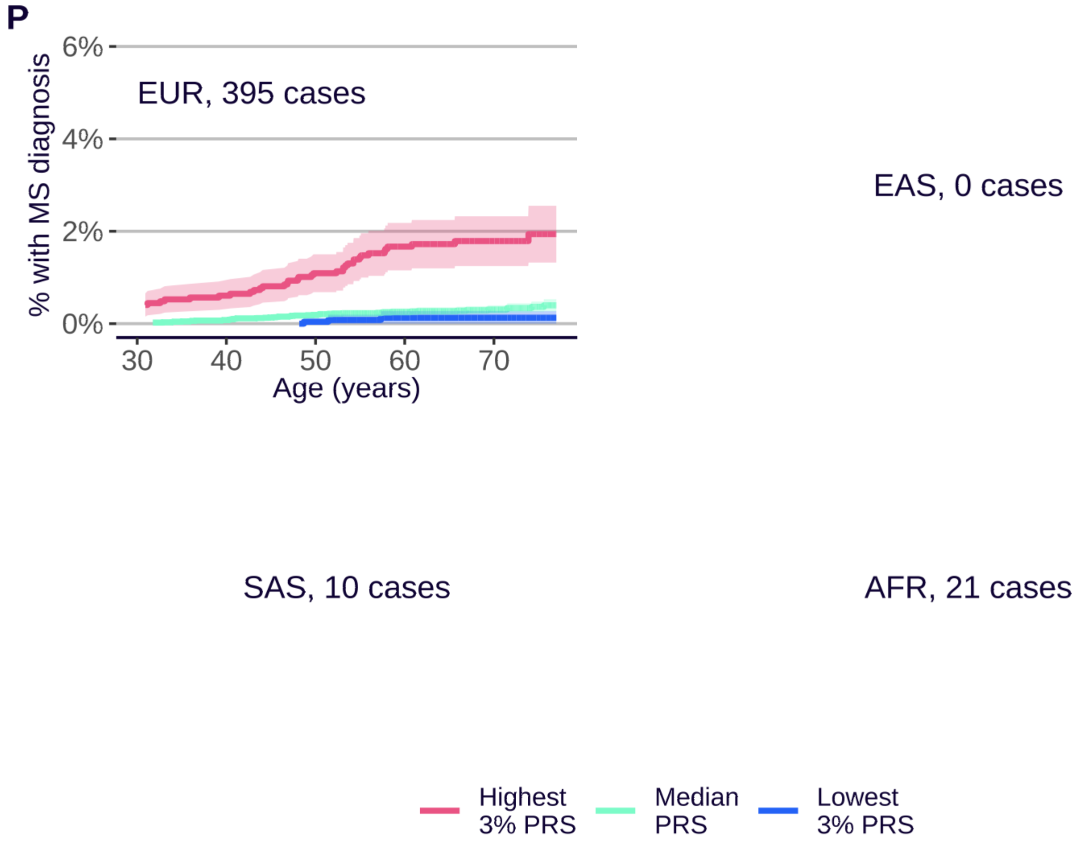
P
% with MS diagnosis
6%
4%
2%
0%
EUR, 395 cases
30
40
50
60
70
Age (years)
EAS, 0 cases
SAS, 10 cases
AFR, 21 cases
Highest 3% PRS
Median PRS
Lowest 3% PRS

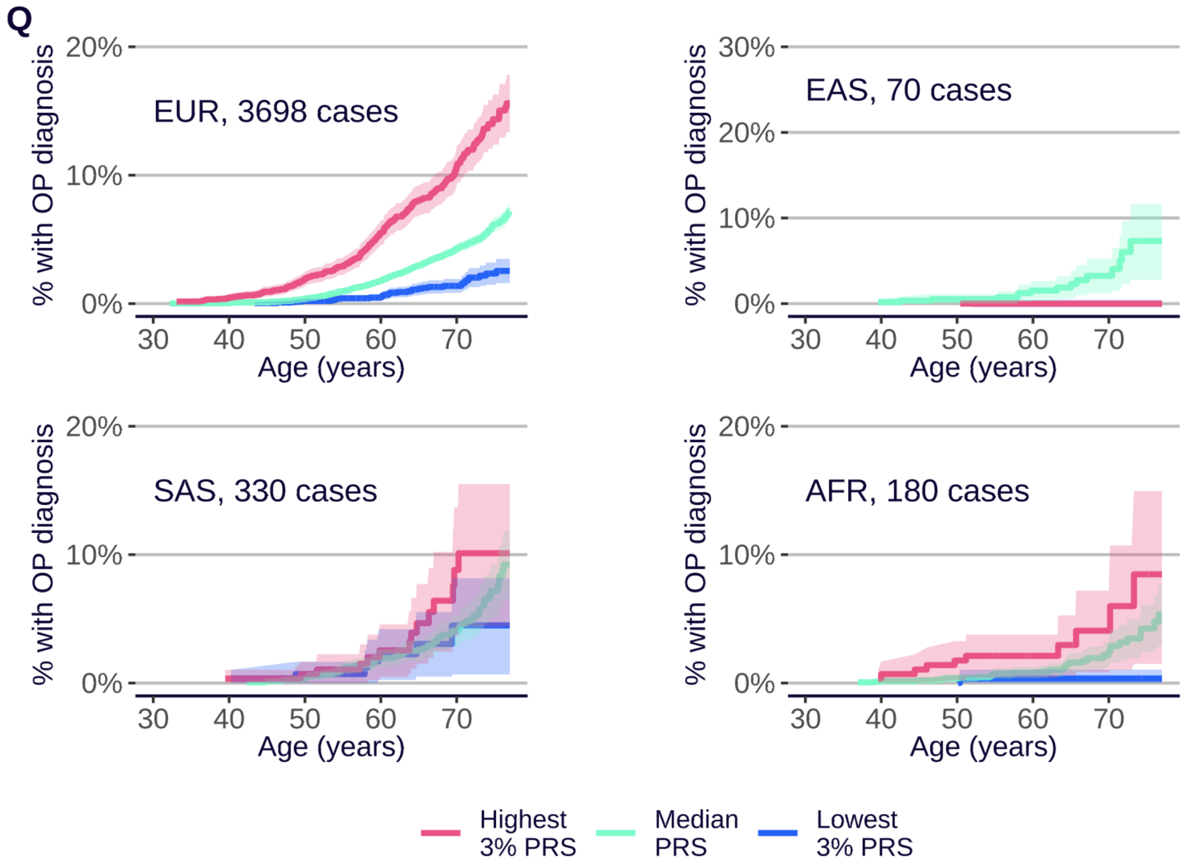
Q
% with OP diagnosis
EUR, 3698 cases
EAS, 70 cases
SAS, 330 cases
AFR, 180 cases
0%
10%
20%
30%
30
40
50
60
70
Age (years)
Highest 3% PRS
Median PRS
Lowest 3% PRS

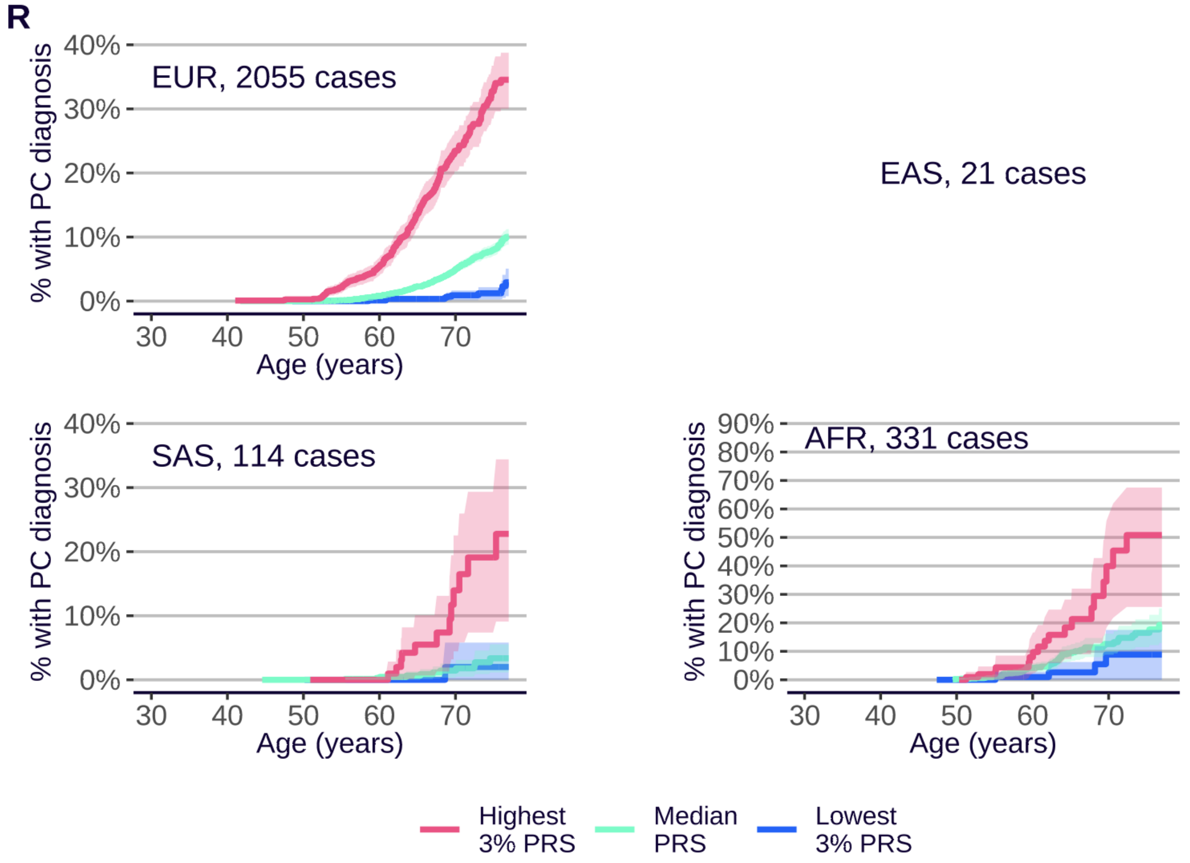
R
% with PC diagnosis
EUR, 2055 cases
EAS, 21 cases
SAS, 114 cases
AFR, 331 cases
0%
10%
20%
30%
40%
50%
60%
70%
80%
90%
30
40
50
60
70
Age (years)
Highest 3% PRS
Median PRS
Lowest 3% PRS

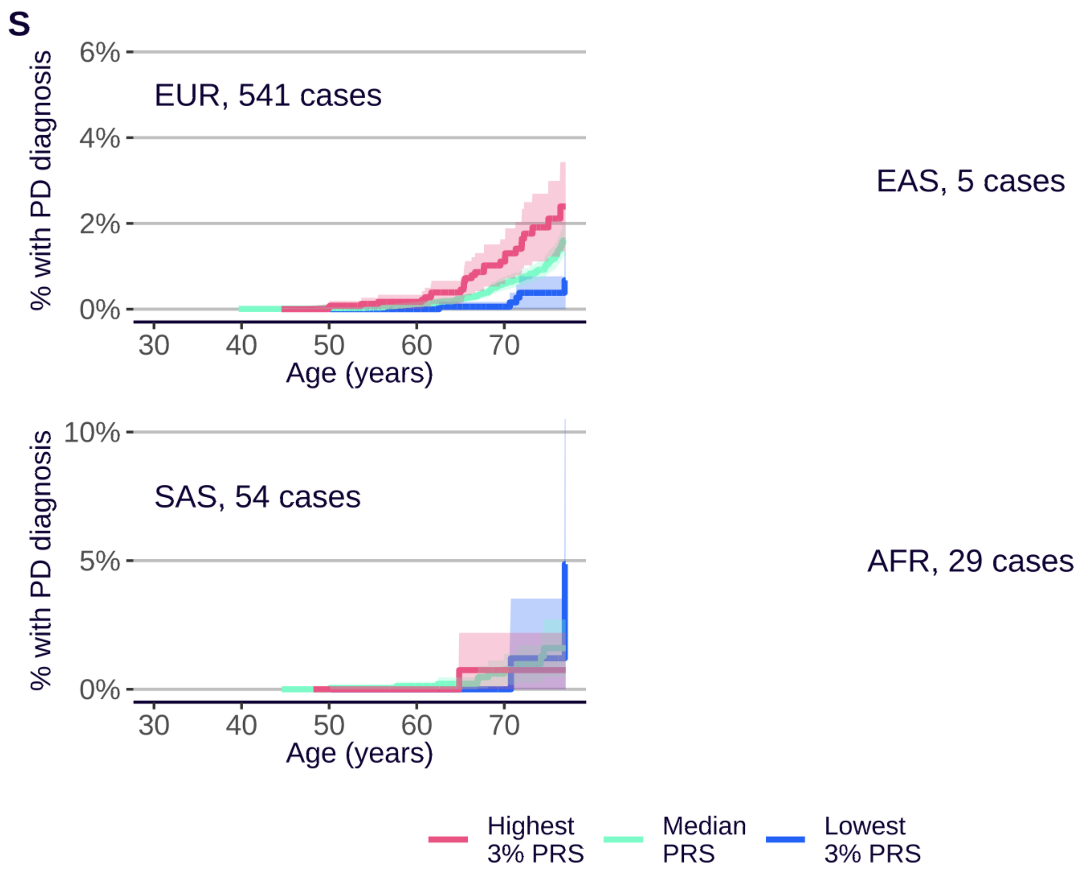
S
% with PD diagnosis
EUR, 541 cases
6%
4%
2%
0%
30
40
50
60
70
Age (years)
EAS, 5 cases
SAS, 54 cases
10%
5%
0%
AFR, 29 cases
Highest 3% PRS
Median PRS
Lowest 3% PRS

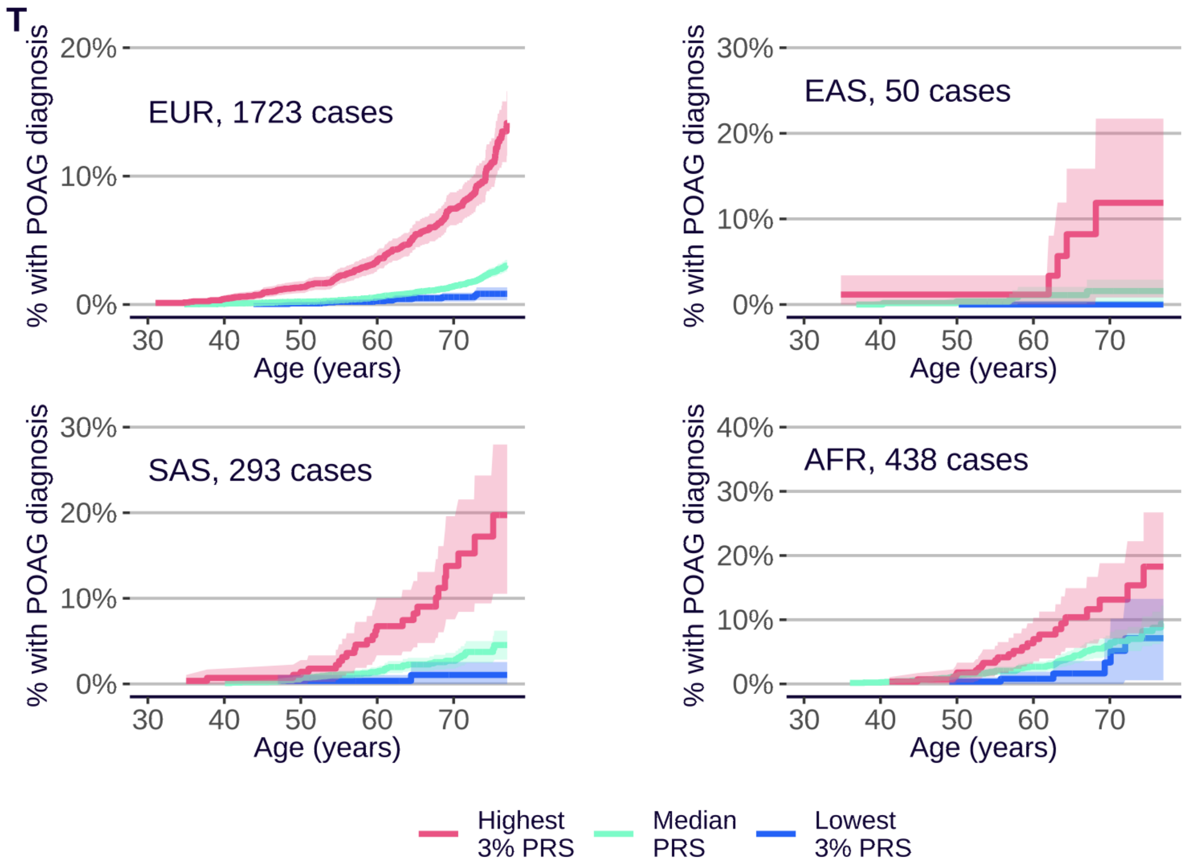
T
% with POAG diagnosis
EUR, 1723 cases
20%
10%
0%
Age (years)
EAS, 50 cases
30%
SAS, 293 cases
AFR, 438 cases
40%
Highest 3% PRS
Median PRS
Lowest 3% PRS

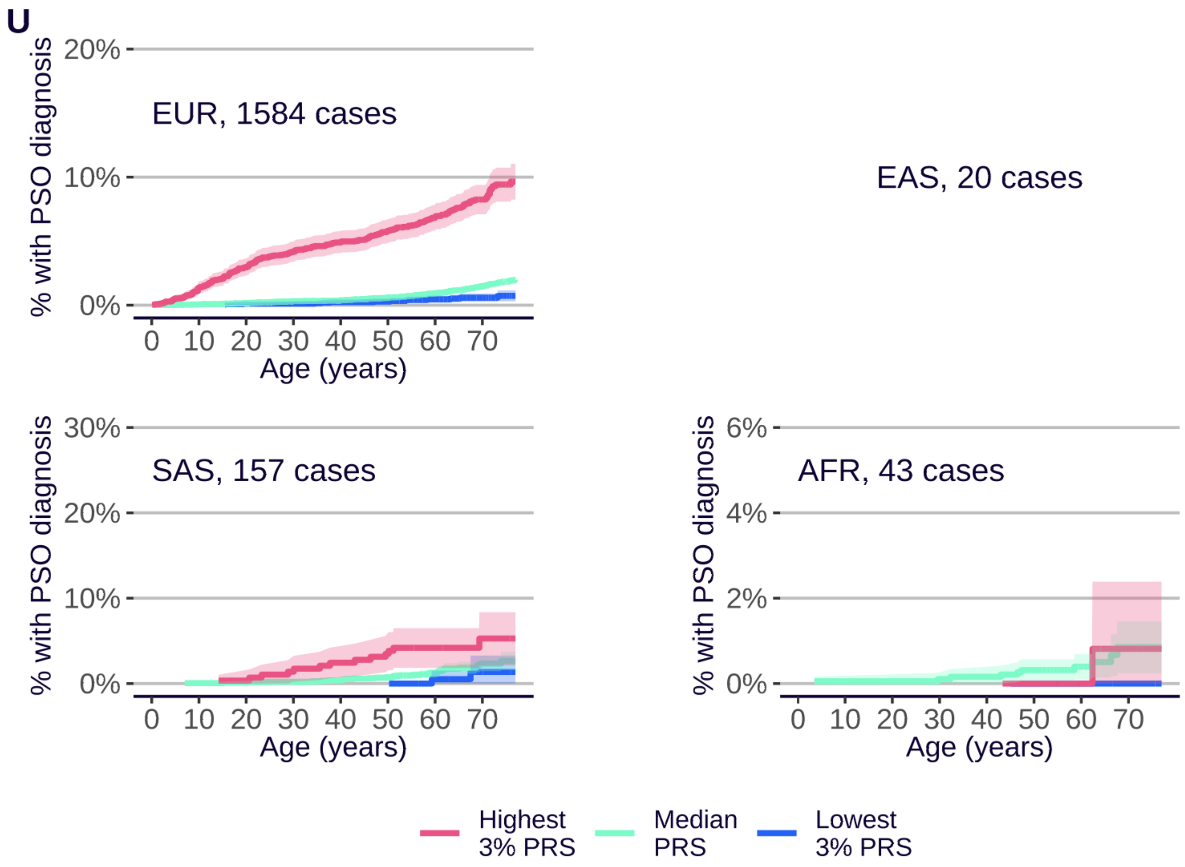
U
EUR, 1584 cases
EAS, 20 cases
SAS, 157 cases
AFR, 43 cases
% with PSO diagnosis
Age (years)
Highest 3% PRS
Median PRS
Lowest 3% PRS

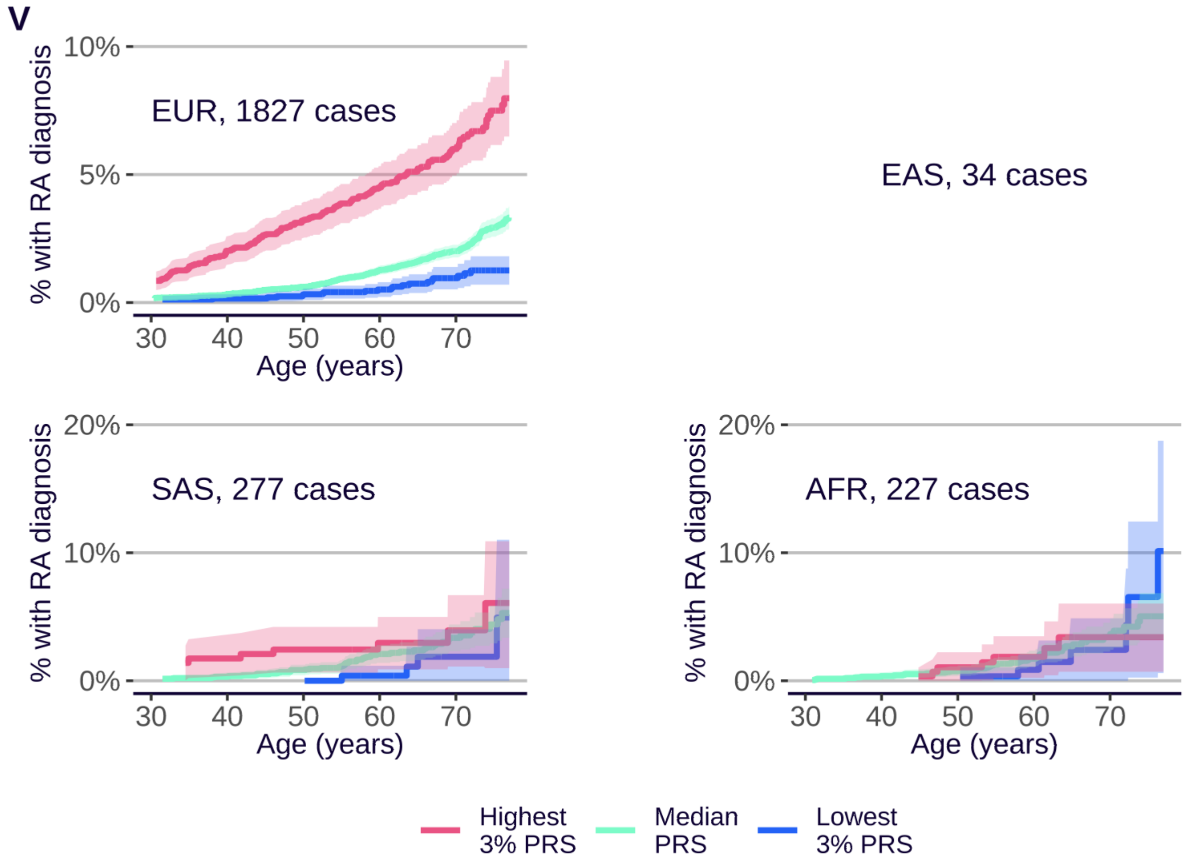
V
EUR, 1827 cases
EAS, 34 cases
SAS, 277 cases
AFR, 227 cases
% with RA diagnosis
Age (years)
Highest 3% PRS
Median PRS
Lowest 3% PRS

**W**

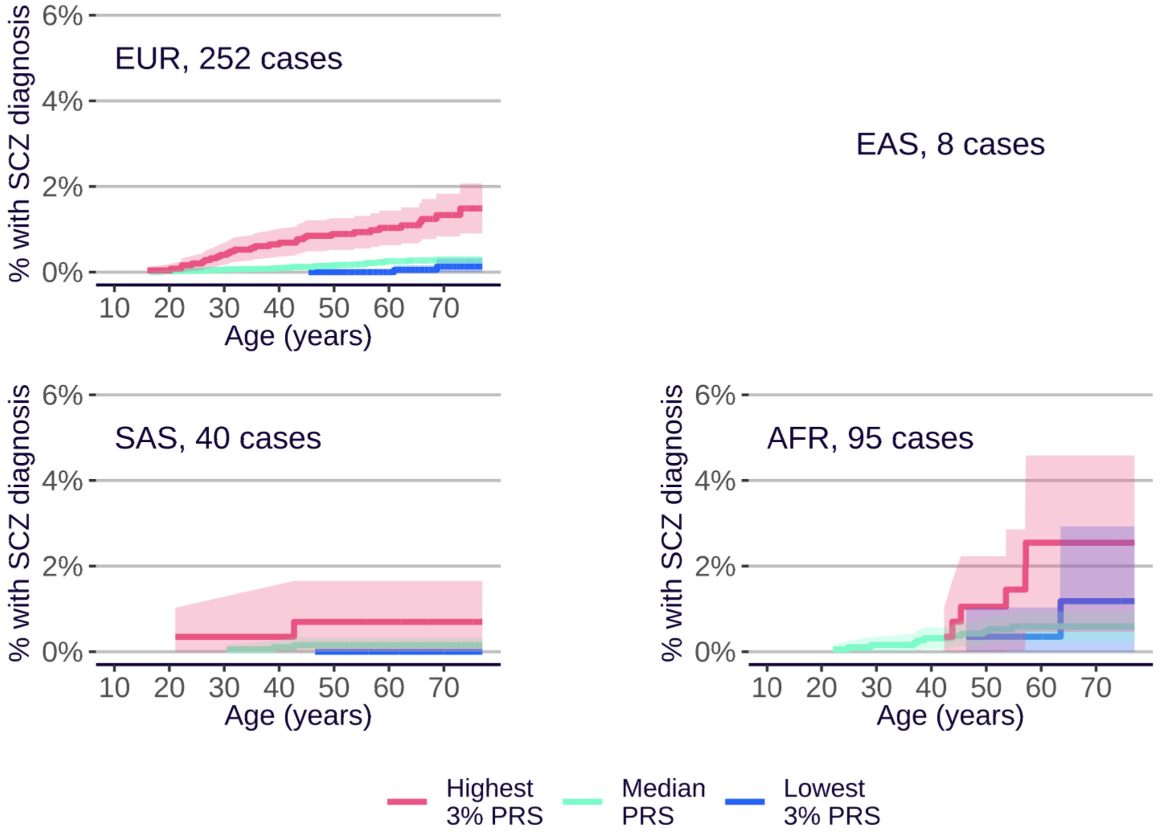

EUR, 252 cases
EAS, 8 cases
SAS, 40 cases
AFR, 95 cases
% with SCZ diagnosis
6%
4%
2%
0%
10 20 30 40 50 60 70
Age (years)
Highest 3% PRS
Median PRS
Lowest 3% PRS


**X**

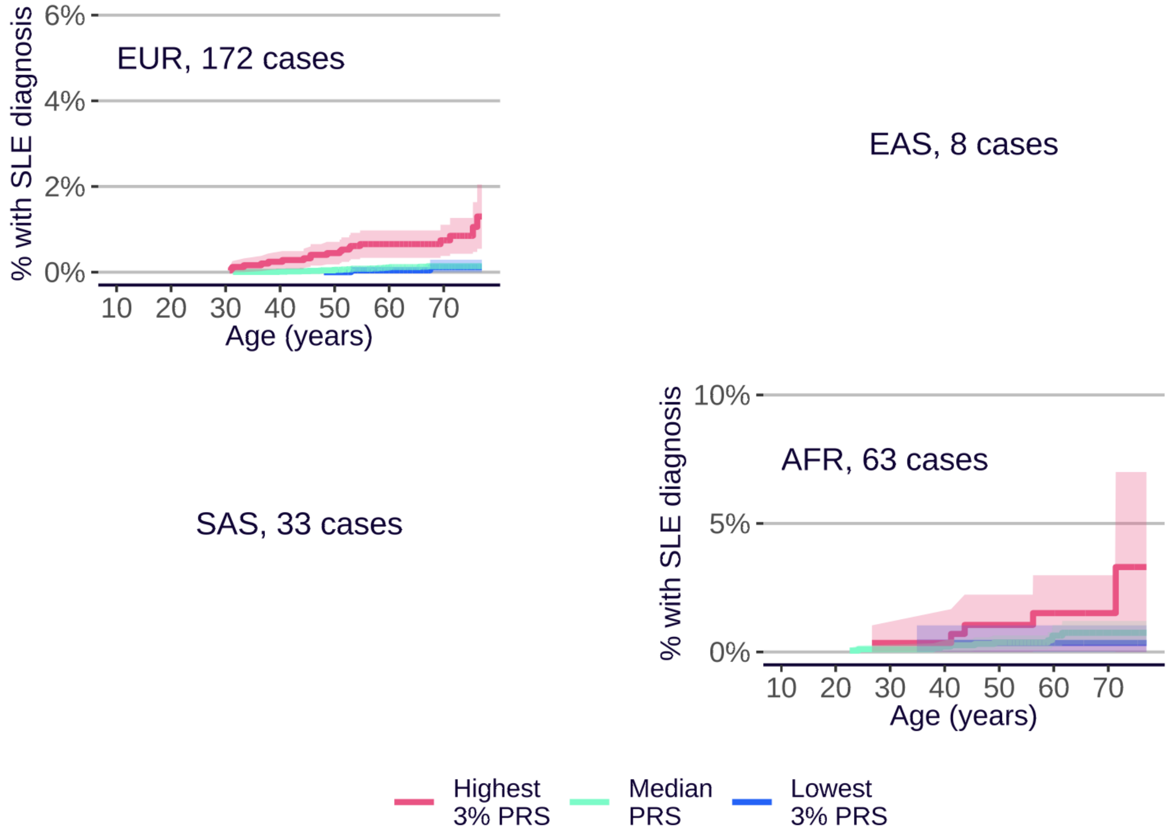

EUR, 172 cases
EAS, 8 cases
SAS, 33 cases
AFR, 63 cases
% with SLE diagnosis
6%
4%
2%
0%
10%
5%
10 20 30 40 50 60 70
Age (years)
Highest 3% PRS
Median PRS
Lowest 3% PRS

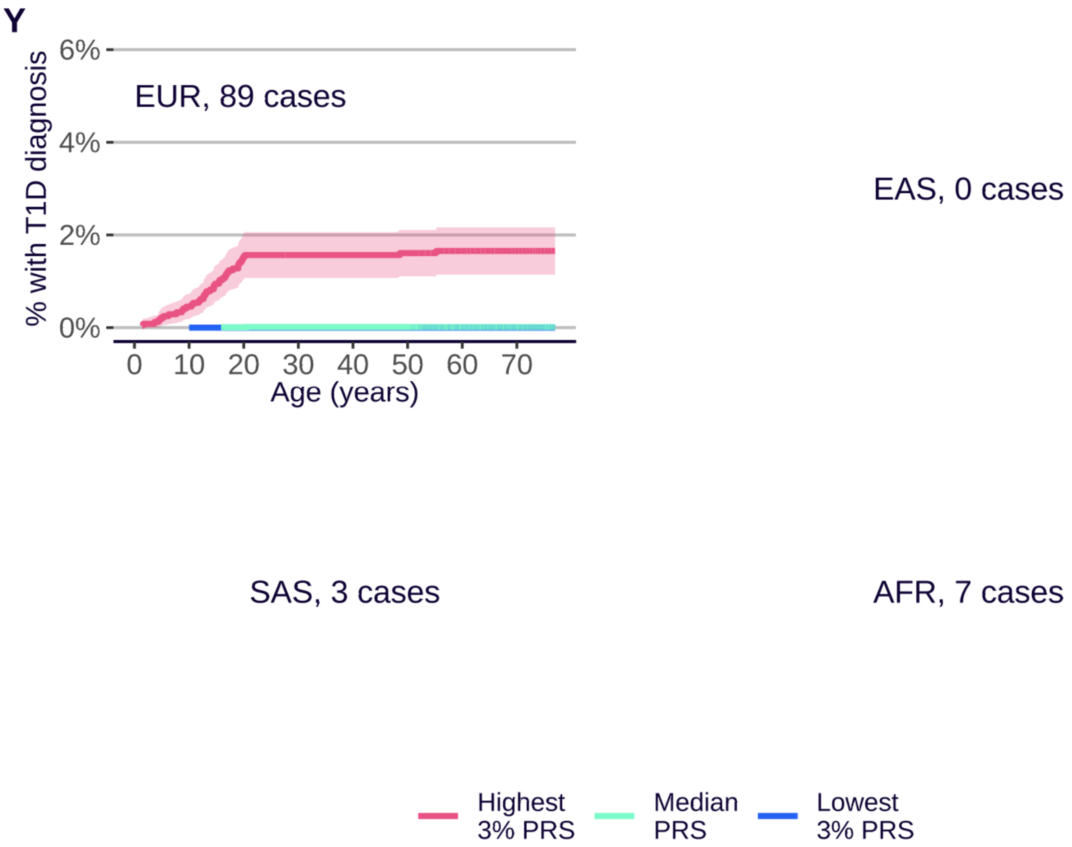
Y
% with T1D diagnosis
6%
4%
2%
0%
EUR, 89 cases
0
10
20
30
40
50
60
70
Age (years)
EAS, 0 cases
SAS, 3 cases
AFR, 7 cases
Highest
3% PRS
Median
PRS
Lowest
3% PRS

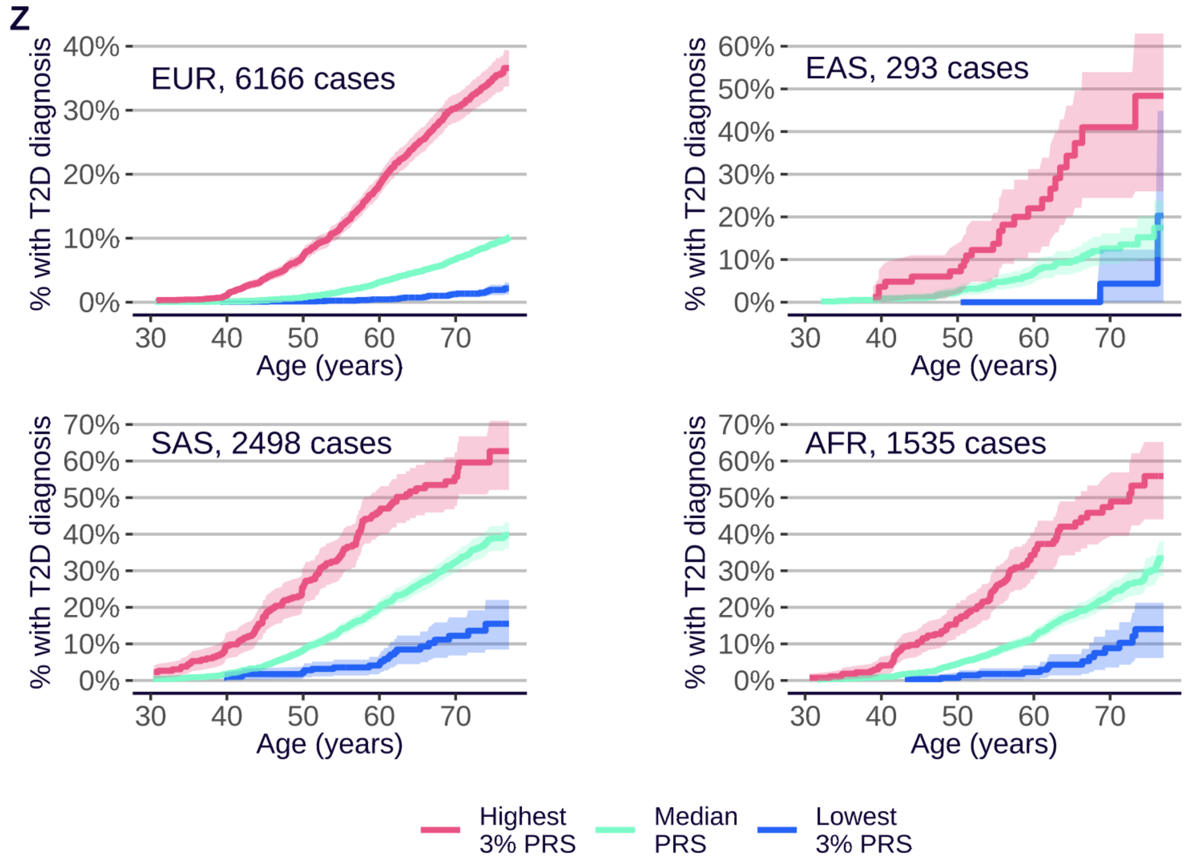
Z
% with T2D diagnosis
EUR, 6166 cases
40%
30%
20%
10%
0%
EAS, 293 cases
60%
50%
40%
30%
20%
10%
0%
SAS, 2498 cases
70%
60%
50%
40%
30%
20%
10%
0%
AFR, 1535 cases
30
40
50
60
70
Age (years)
Highest
3% PRS
Median
PRS
Lowest
3% PRS

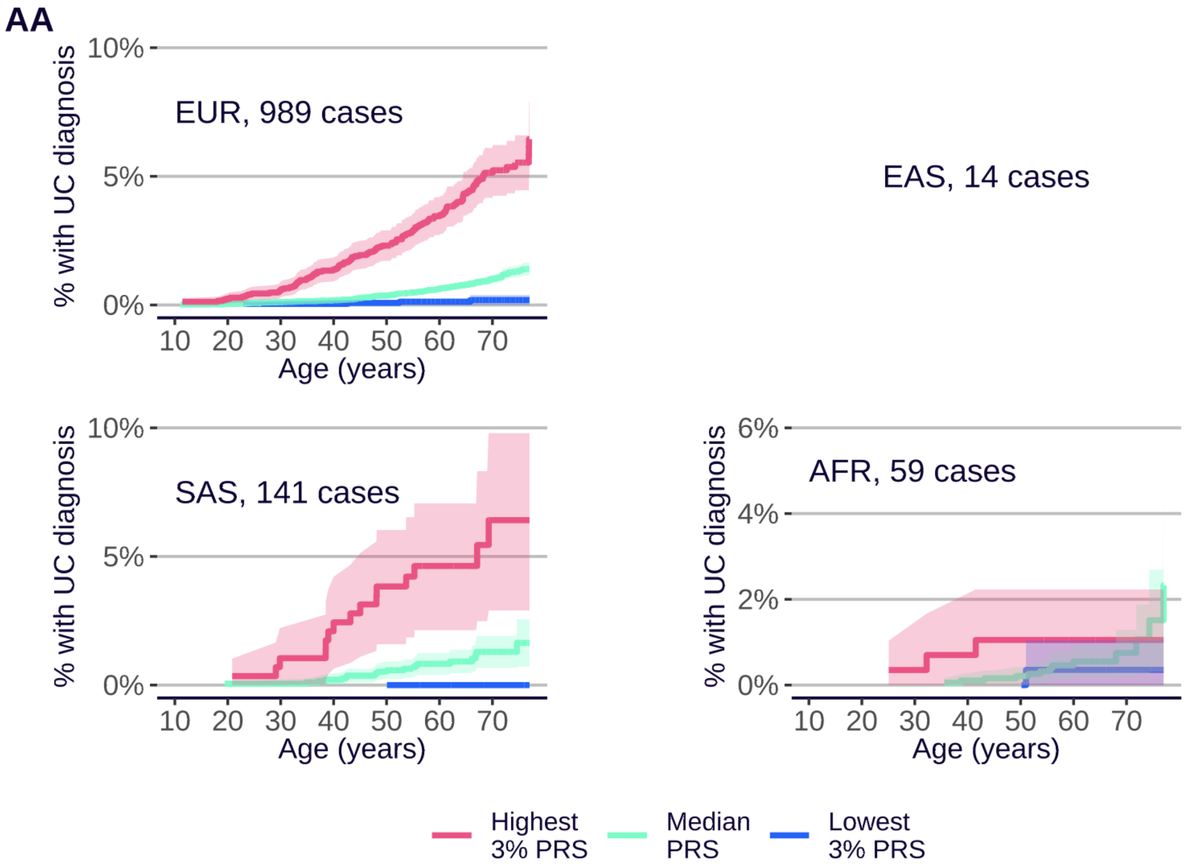
AA
EUR, 989 cases
EAS, 14 cases
SAS, 141 cases
AFR, 59 cases
% with UC diagnosis
Age (years)
Highest 3% PRS
Median PRS
Lowest 3% PRS

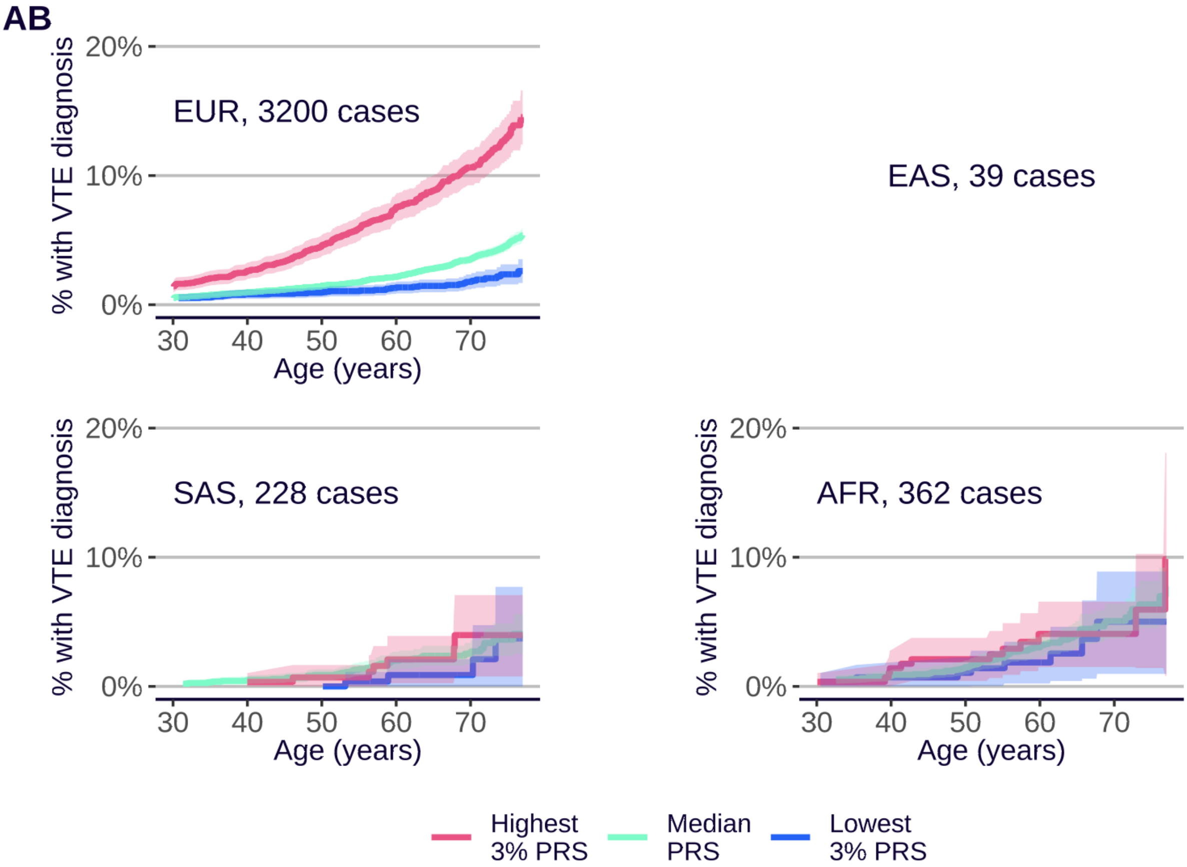
AB
EUR, 3200 cases
EAS, 39 cases
SAS, 228 cases
AFR, 362 cases
% with VTE diagnosis
Age (years)
Highest 3% PRS
Median PRS
Lowest 3% PRS

**Supplementary Figure 11**. **Cumulative incidence plots by ancestry, Enhanced PRS Set**. Cumulative incidence plots are shown for each disease and each ancestry group in the UKB Testing Subgroup, provided more than 40 cases are available (the number of cases is printed otherwise), with separate curves for the highest 3% (red), lowest 3% (blue), and median (green) of the PRS distribution. **A**. Alzheimer's disease (AD). **B**. Atrial fibrillation (AF). **C**. Age-related macular degeneration (AMD). **D**. Asthma (AST). **E**. Breast cancer (BC), **F**. Bipolar disorder (BD), **G**. Coronary artery disease (CAD). **H**. Crohn's disease (CD). **I**. Coeliac disease (CED). **J**. Bowel cancer (CRC). **K**. Cardiovascular disease (CVD), **L**. Epithelial ovarian cancer (EOC). **M**. Hypertension (HT). **N**. Ischaemic stroke (ISS). **O**. Melanoma (MEL). **P**. Multiple sclerosis (MS). **Q**. Osteoporosis (OP). **R**. Prostate cancer (PC). **S**. Parkinson's disease (PD). **T**. Primary open angle glaucoma (POAG). **U**. Psoriasis (PSO). **V**. Rheumatoid arthritis (RA). **W**. Schizophrenia (SCZ). **X**. Systemic lupus erythematosus (SLE). **Y**. Type 1 diabetes (T1D). **Z**. Type 2 diabetes (T2D). **AA**. Ulcerative colitis (UC). **AB**. Venous thromboembolic disease (VTE). EUR = European ancestry. EAS = East Asian ancestry. SAS = South Asian ancestry. AFR = Sub-Saharan African ancestry. Shaded areas indicate 95% CI.

A

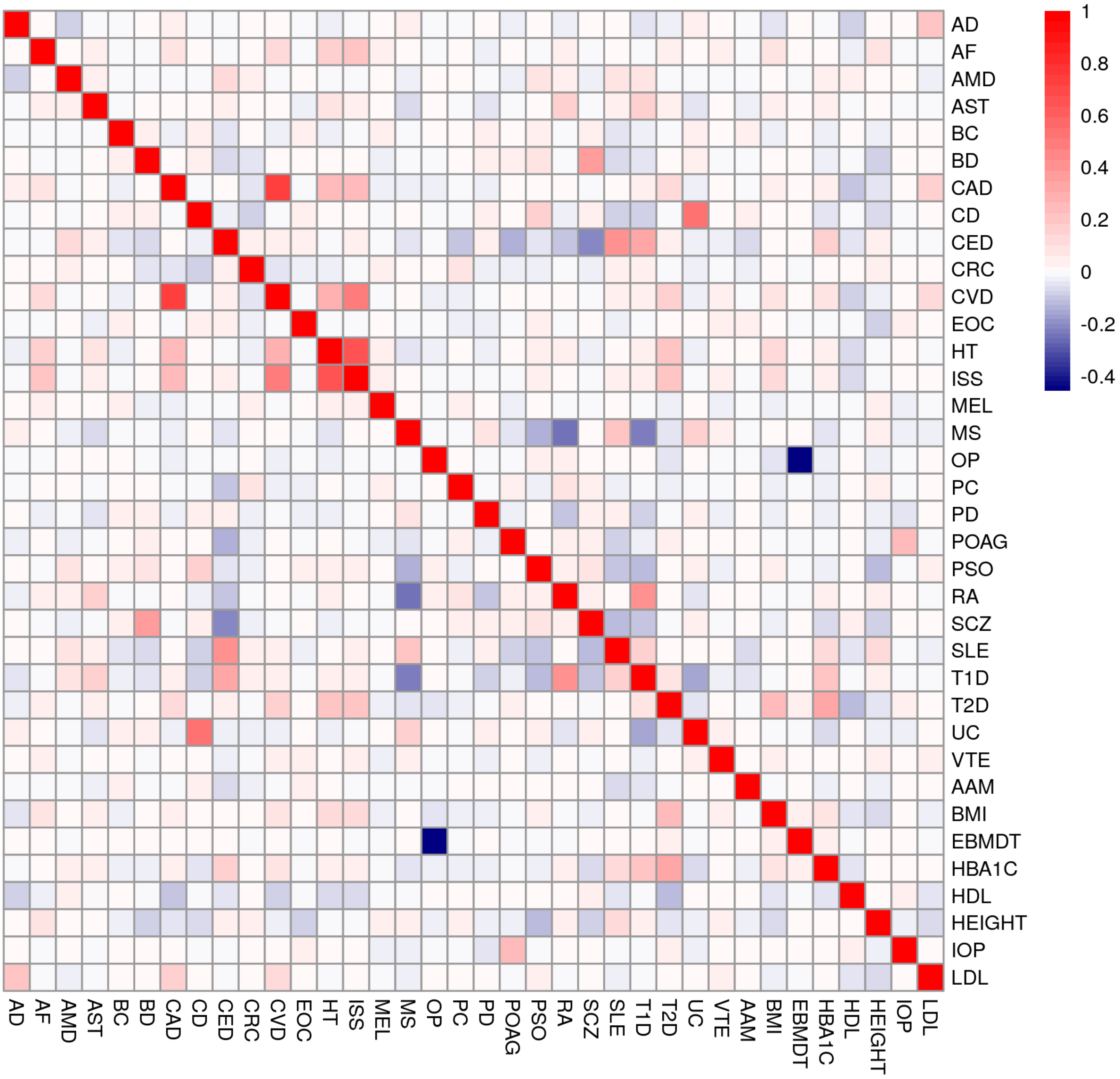
AD
AF
AMD
AST
BC
BD
CAD
CD
CED
CRC
CVD
EOC
HT
ISS
MEL
MS
OP
PC
PD
POAG
PSO
RA
SCZ
SLE
T1D
T2D
UC
VTE
AAM
BMI
EBMDT
HBA1C
HDL
HEIGHT
IOP
LDL
1
0.8
0.6
0.4
0.2
0
-0.2
-0.4

**B**

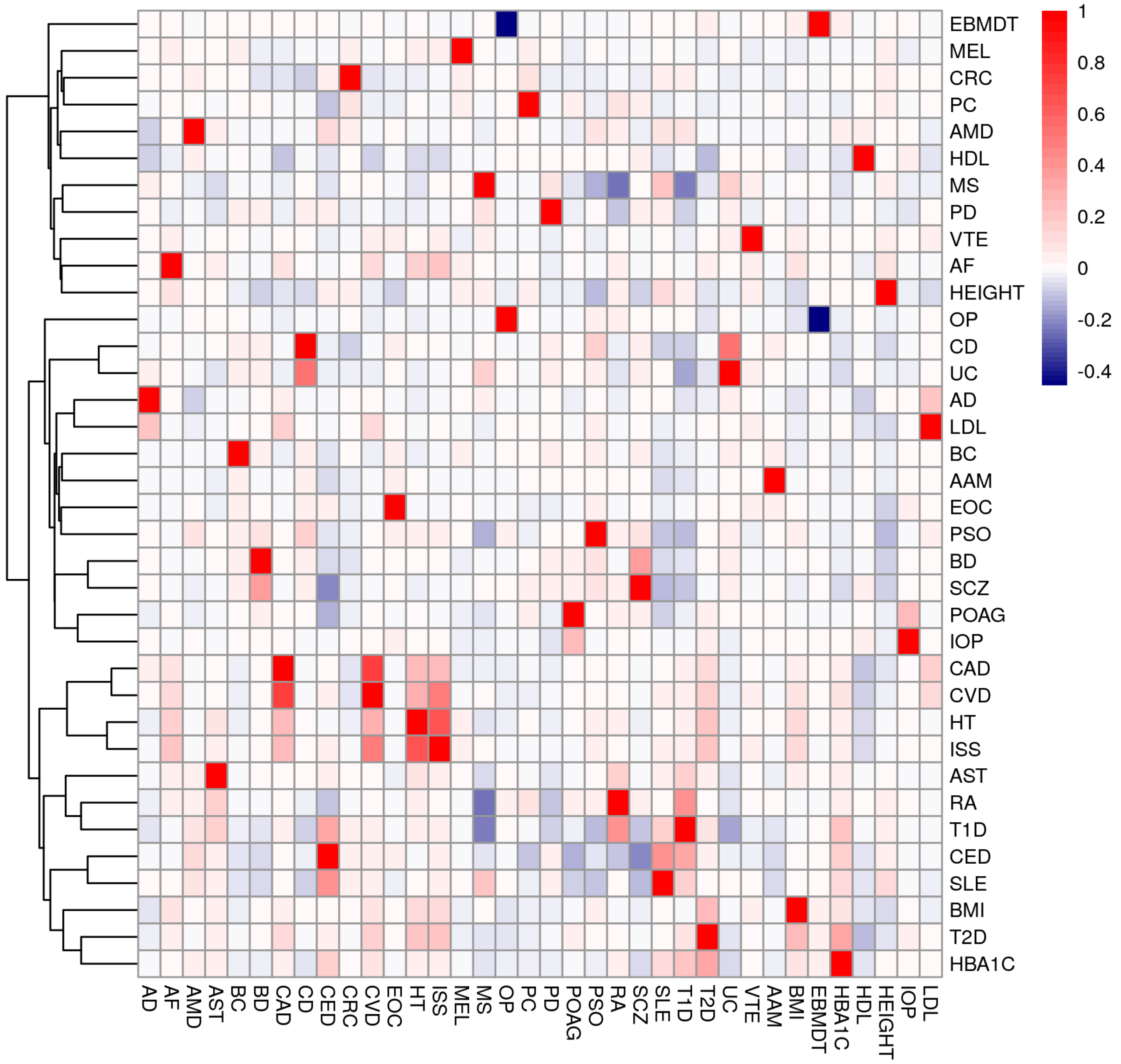
EBMDT
MEL
CRC
PC
AMD
HDL
MS
PD
VTE
AF
HEIGHT
OP
CD
UC
AD
LDL
BC
AAM
EOC
PSO
BD
SCZ
POAG
IOP
CAD
CVD
HT
ISS
AST
RA
T1D
CED
SLE
BMI
T2D
HBA1C
AD AF AMD AST BC BD CAD CD CED CRC CVD EOC HT ISS MEL MS OP PC PD POAG PSO RA SCZ SLE T1D T2D UC VTE AAM BMI EBMDT HBA1C HDL HEIGHT IOP LDL
1
0.8
0.6
0.4
0.2
0
-0.2
-0.4

C

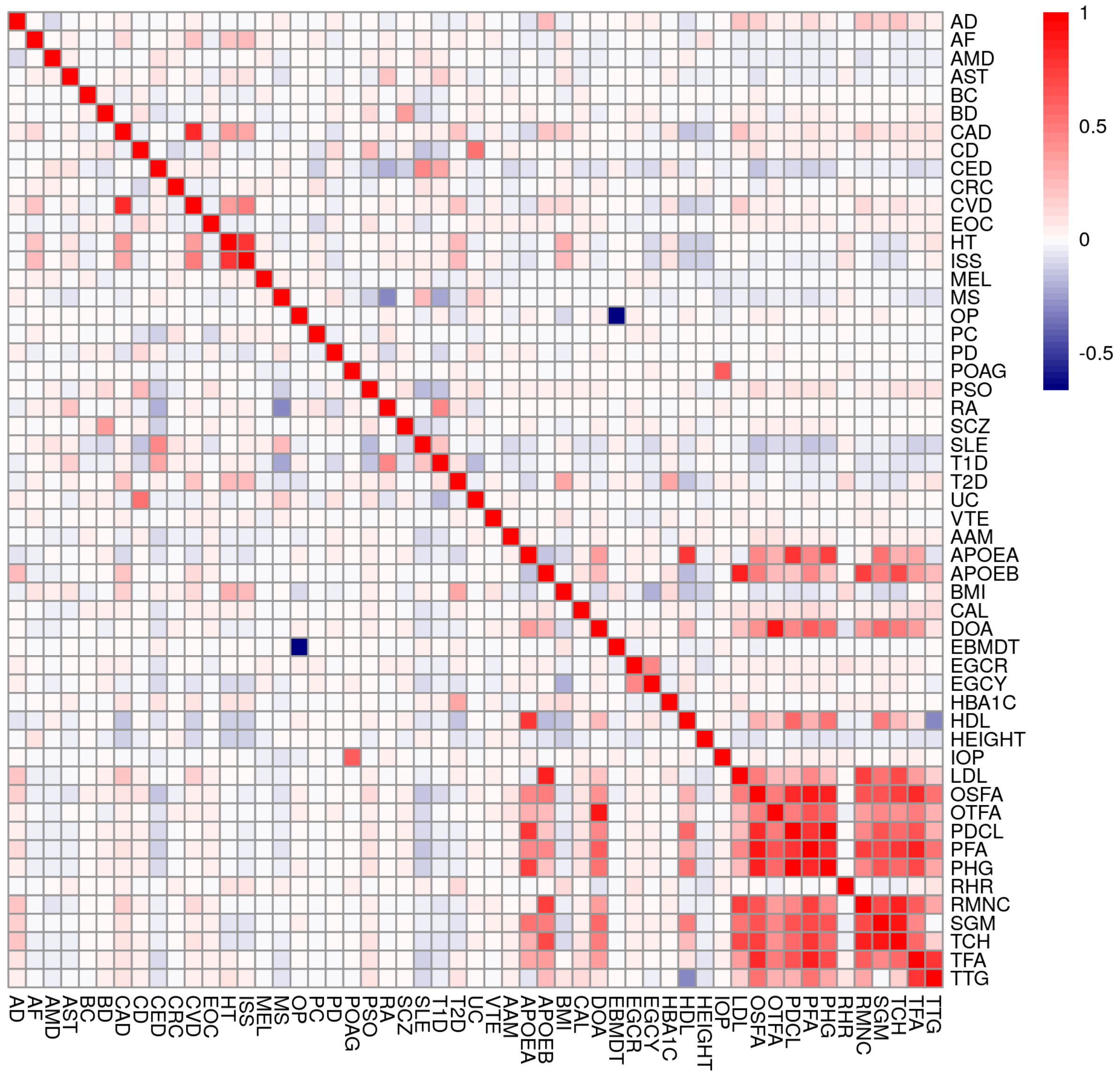

AD
AF
AMD
AST
BC
BD
CAD
CD
CED
CRC
CVD
EOC
HT
ISS
MEL
MS
OP
PC
PD
POAG
PSO
RA
SCZ
SLE
T1D
T2D
UC
VTE
AAM
APOEA
APOEB
BMI
CAL
DOA
EBMDT
EGCR
EGCY
HBA1C
HDL
HEIGHT
IOP
LDL
OSFA
OTFA
PDCL
PFA
PHG
RHR
RMNC
SGM
TCH
TFA
TTG
1
0.5
0
-0.5

**D**

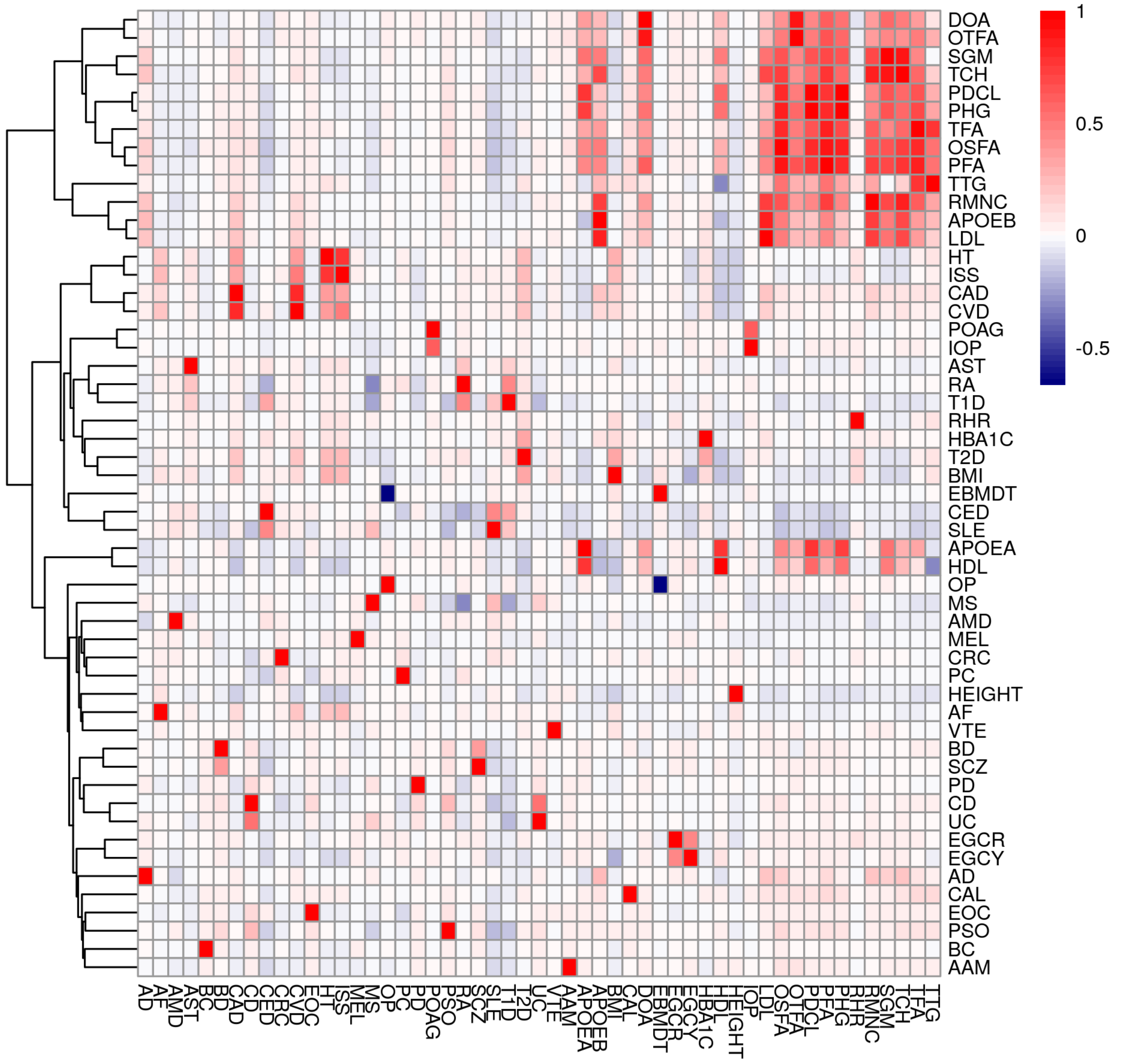


**Supplementary Figure 12. Heatmaps of correlations among PRS scores**. All correlations are calculated from European ancestry individuals in the UKB Testing Subgroup. **A** Correlations among the Standard Set, 28 disease codes ordered alphabetically, then 8 quantitative trait codes ordered alphabetically (recall the Standard Set has fewer quantitative traits than the Enhanced Set). **B** Correlations among the Standard Set, ordered according to a hierarchical clustering dendrogram (complete linkage on Euclidean distance, see hclust() function in R). **C** Correlations among the Enhanced Set, 28 disease codes ordered alphabetically, then 25 quantitative trait codes ordered alphabetically. **D** Correlations among the Enhanced Set, ordered according to a hierarchical clustering dendrogram (complete linkage on Euclidean distance, see hclust() function in R). See Supplementary Table 5 for trait code mappings.

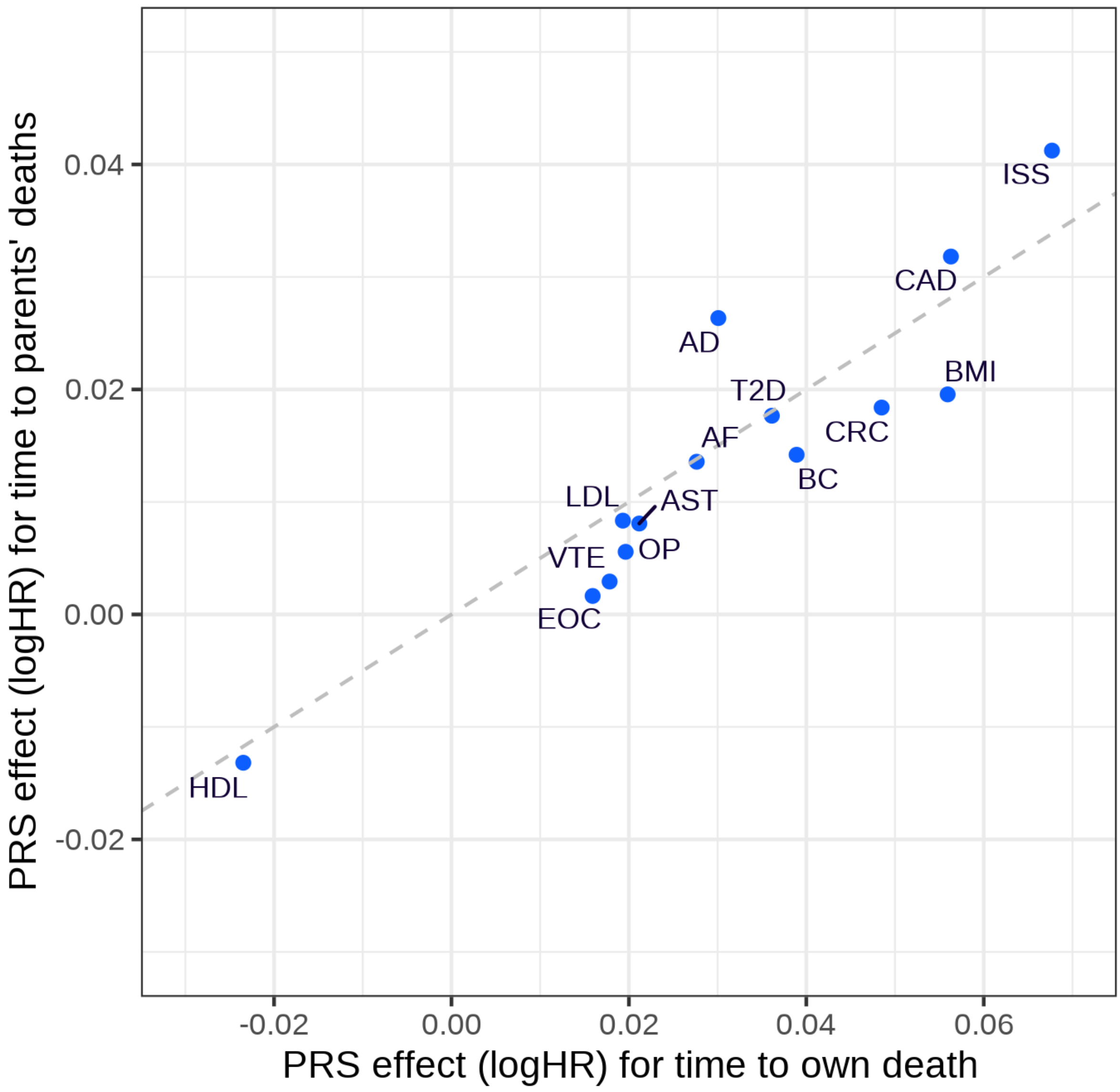


**Supplementary Figure 13. Multivariate PRS model for all-cause mortality**. Traits are shown if selected by stepwise regression of time-to-death from first assessment, if selected both for participants' own death and for their parents' death (maternal and paternal data entered as separate observations). Dashed line shows the expected parent:offspring effect size ratio of 1:2. See Supplementary Table 5 for trait code mappings.

A

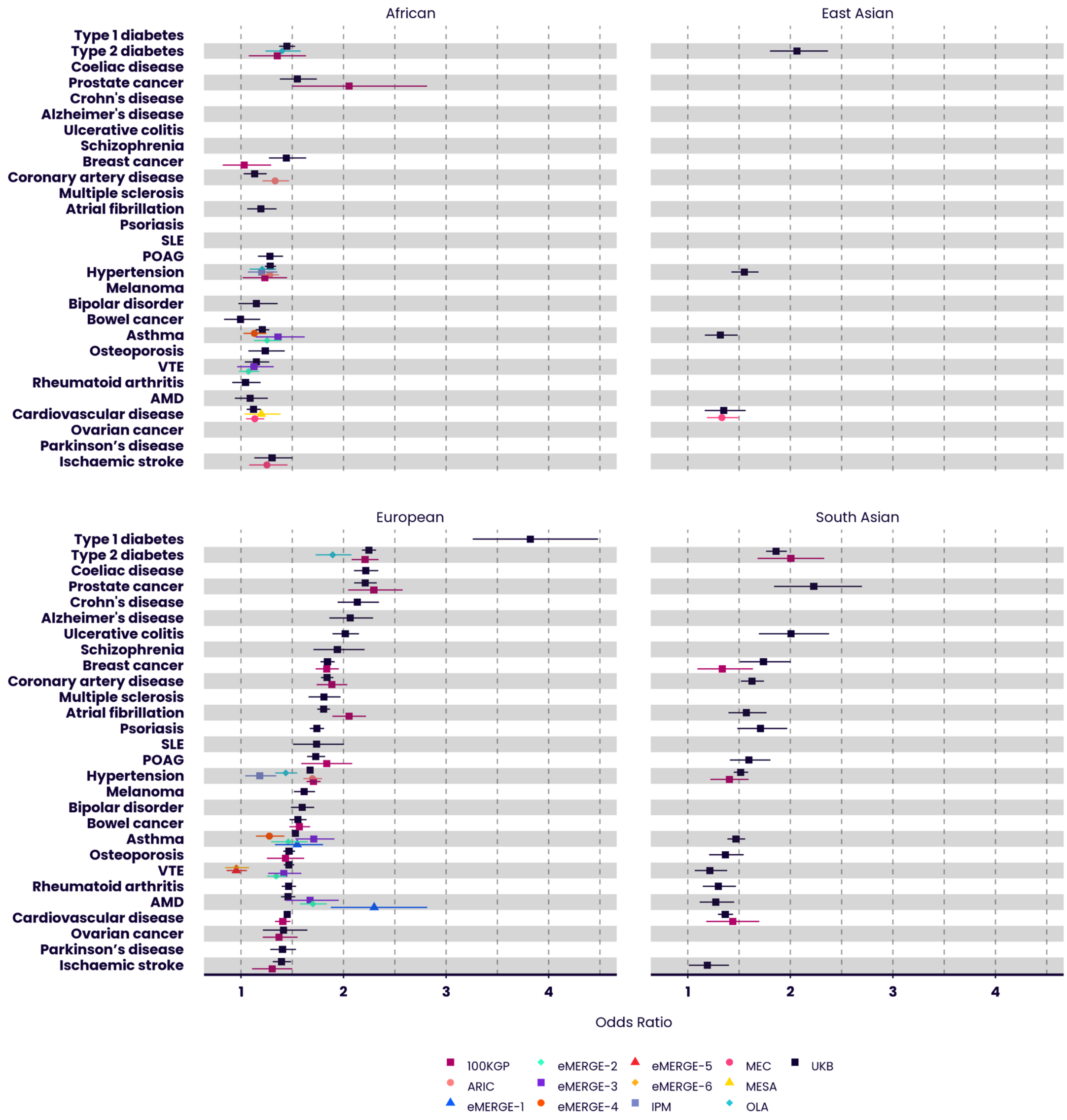
African
East Asian
European
South Asian
Type 1 diabetes
Type 2 diabetes
Coeliac disease
Prostate cancer
Crohn's disease
Alzheimer's disease
Ulcerative colitis
Schizophrenia
Breast cancer
Coronary artery disease
Multiple sclerosis
Atrial fibrillation
Psoriasis
SLE
POAG
Hypertension
Melanoma
Bipolar disorder
Bowel cancer
Asthma
Osteoporosis
VTE
Rheumatoid arthritis
AMD
Cardiovascular disease
Ovarian cancer
Parkinson's disease
Ischaemic stroke
1
2
3
4
Odds Ratio
100KGP
ARIC
eMERGE-1
eMERGE-2
eMERGE-3
eMERGE-4
eMERGE-5
eMERGE-6
IPM
MEC
MESA
OLA
UKB

B

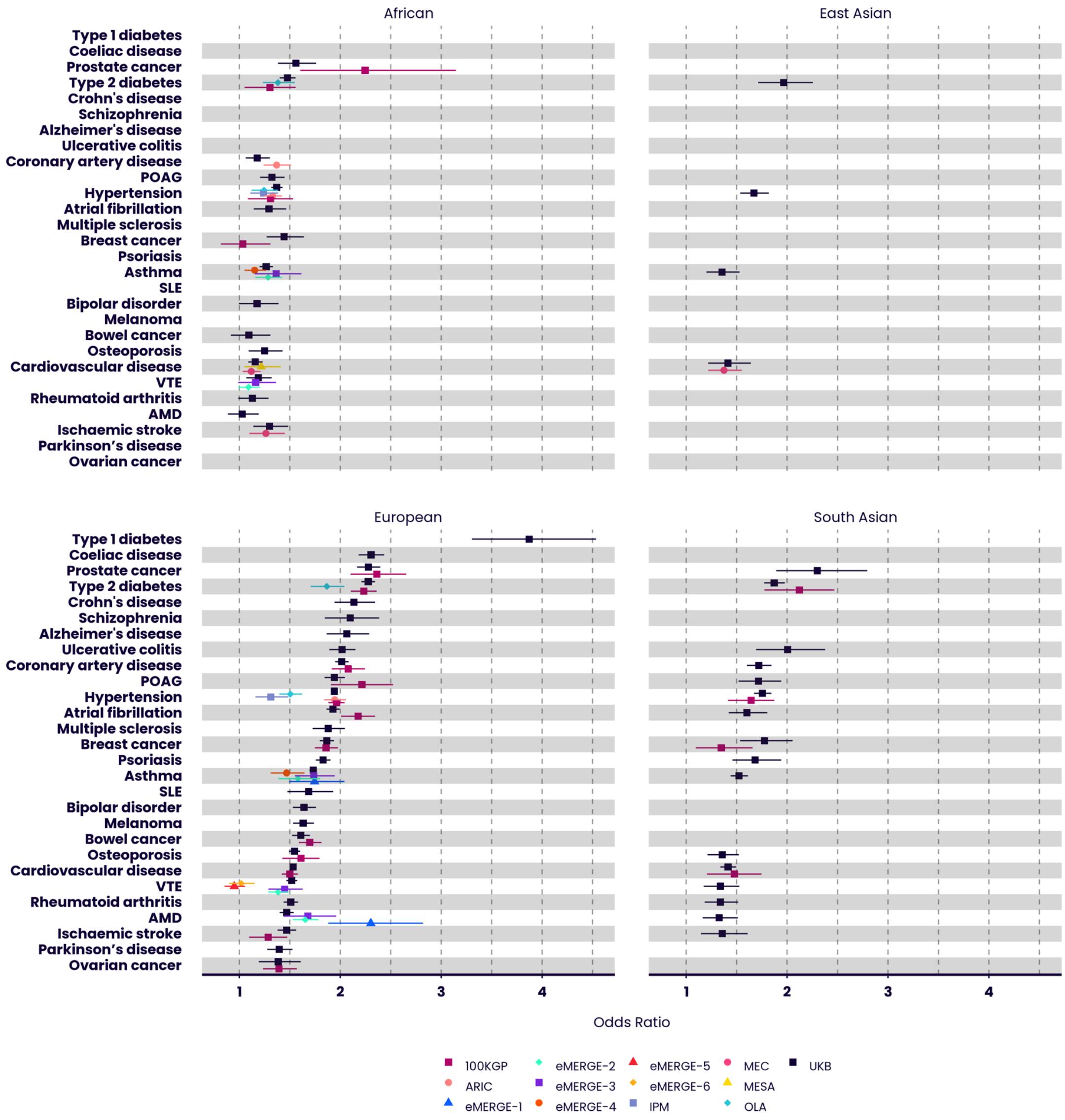
African
East Asian
European
South Asian
Type 1 diabetes
Coeliac disease
Prostate cancer
Type 2 diabetes
Crohn's disease
Schizophrenia
Alzheimer's disease
Ulcerative colitis
Coronary artery disease
POAG
Hypertension
Atrial fibrillation
Multiple sclerosis
Breast cancer
Psoriasis
Asthma
SLE
Bipolar disorder
Melanoma
Bowel cancer
Osteoporosis
Cardiovascular disease
VTE
Rheumatoid arthritis
AMD
Ischaemic stroke
Parkinson's disease
Ovarian cancer
1
2
3
4
Odds Ratio
100KGP
ARIC
eMERGE-1
eMERGE-2
eMERGE-3
eMERGE-4
eMERGE-5
eMERGE-6
IPM
MEC
MESA
OLA
UKB

C

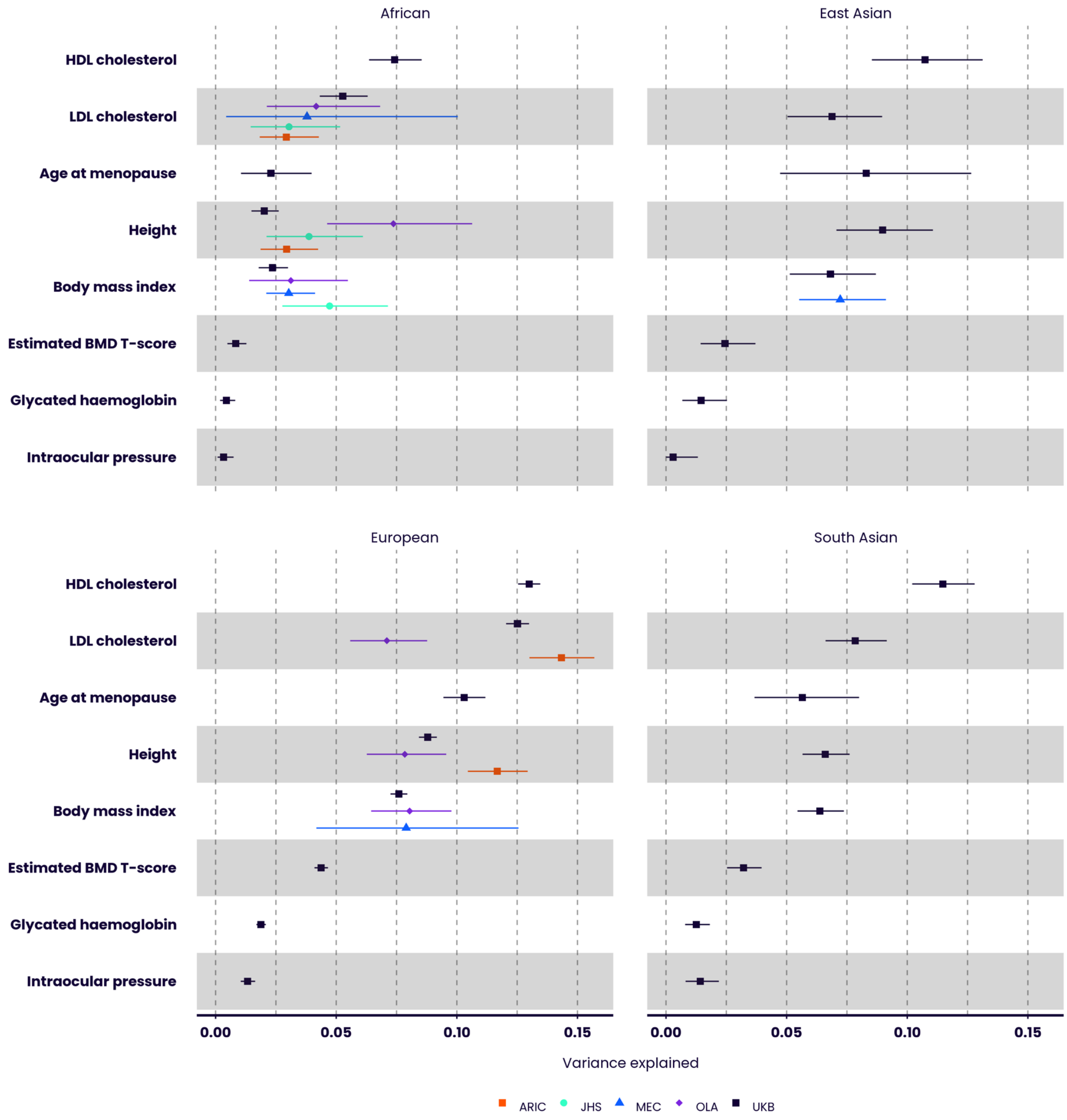
African
East Asian
European
South Asian
HDL cholesterol
LDL cholesterol
Age at menopause
Height
Body mass index
Estimated BMD T-score
Glycated haemoglobin
Intraocular pressure
0.00
0.05
0.10
0.15
Variance explained
ARIC
JHS
MEC
OLA
UKB

D

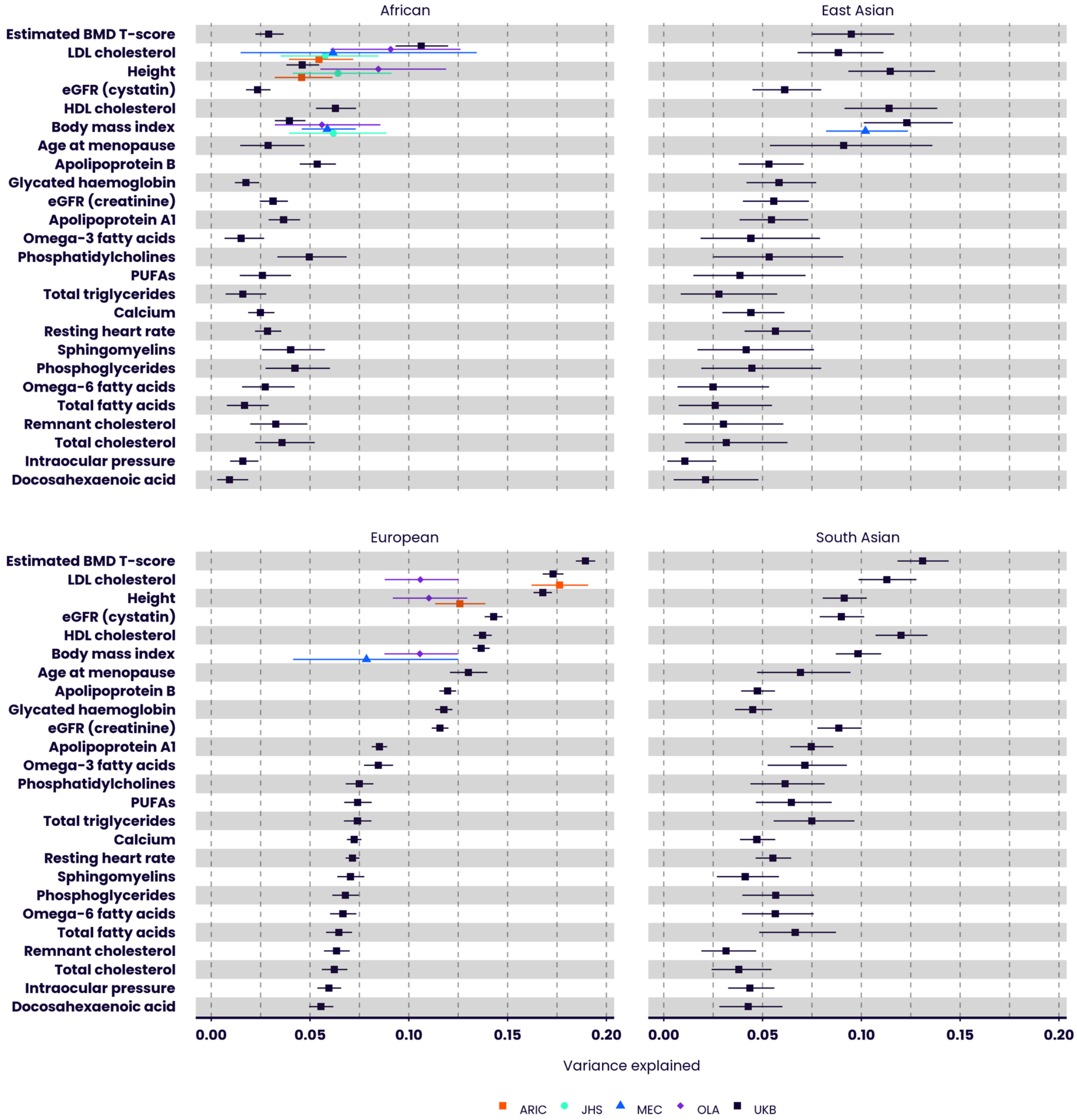


**Supplementary Figure 14. Performance of the UK Biobank PRS Release algorithms in other cohorts**. Performance (odds ratio per SD of PRS) of the disease trait PRSs, stratified by genetically inferred ancestry, in different cohorts, for the Standard PRSs (**A**) and the Enhanced PRSs (**B**). Results for non-European ancestries are shown if at least 100 cases are available for testing. Performance (proportion of variance in trait explained by PRS, $r^2$) of the quantitative trait PRSs, stratified by genetically inferred ancestry, for the Standard PRSs (**C**) and the Enhanced PRSs (**D**). $r^2$ was used as the performance metric because not all cohorts had information on participants' age. Bars indicate 95% confidence intervals. ARIC = Atherosclerosis Risk in Communities. DRIVE = Discovery, Biology, and Risk of Inherited Variants in Breast Cancer. eMERGE = Electronic Medical Records and Genomics Network. IPM = Institute for Personalized Medicine BioMe Biobank Project. JHS = Jackson Heart Study. MEC = Multi-Ethnic Cohort. MESA = Multi-Ethnic Study of Atherosclerosis. OLA = Omics in Latinos component of the Hispanic Community Health Study / Study of Latinos Project. ROOT = GWAS of Breast Cancer in the African Diaspora (ROOT) study. UKB = UK Biobank. Refer to Figure 1 and 2 for disease and quantitative trait abbreviations respectively.

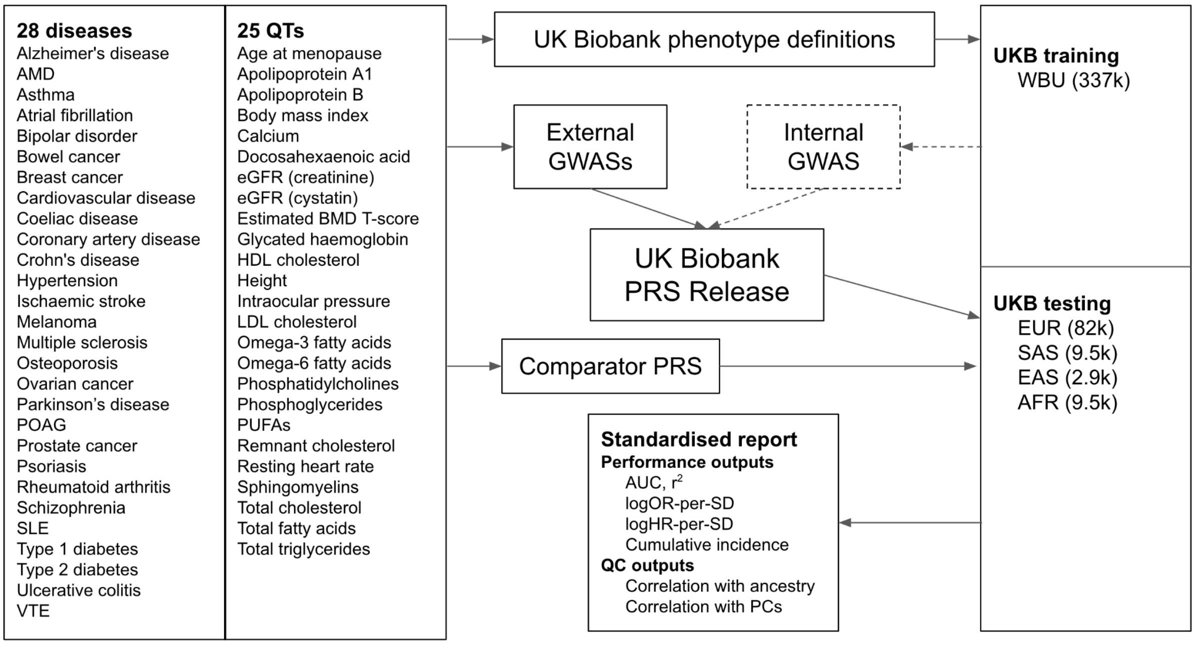


**Supplementary Figure 15**. **Schematic workflow for the generation and standardised evaluation of the UK Biobank PRS Release**. UK Biobank phenotype definitions were developed for 28 diseases (binary case/control status) and 25 quantitative traits, and then applied to internal GWAS generation and standardised PRS evaluation. GWAS datasets were identified and meta-analysed to generate input data for the UK Biobank PRS Release. Where sufficient external GWAS data were available for a trait, a 'Standard' polygenic score was generated for every UK Biobank participant. For all traits, an internal GWAS was generated using the previously described White British Unrelated (WBU) subset of UK Biobank [1], and meta-analysed with other GWAS data to generate an 'Enhanced' polygenic score for every participant in the UK Biobank Testing Subgroup. An independent multi-ancestry UK Biobank Testing Subgroup was used for reporting standardized performance and QC outputs. EUR = European ancestry. SAS = South Asian ancestry. EAS = East Asian ancestry. AFR = African (Sub-Saharan) ancestry. Refer to Figure 1 and 2 for disease and quantitative trait abbreviations respectively.
